# Highly scalable technology-assisted differential diagnostics of ASD

**DOI:** 10.1101/2025.10.17.25338146

**Authors:** Irene Sophia Plank, Jana C. Koehler, Jonathan Eckelmann, Afton M. Bierlich, Richard Musil, Nikolaos Koutsouleris, Christine M. Falter-Wagner

## Abstract

Diagnosing autism spectrum disorder (ASD) in adulthood is time-consuming and markedly complicated by the requirement to distinguish between ASD and differential diagnoses also associated with social interaction difficulties, such as Borderline Personality Disorder (BPD) – a distinction for which currently no valid screening or diagnostic tool exists. While technology-assisted diagnostics (TAD) has emerged, existing algorithms have focused on classifying between ASD and no diagnosis, not fully addressing clinical reality. Therefore, we assessed the feasibility of TAD for differential diagnostics by classifying between ASD and BPD. We extracted features from live reciprocal conversations, allowing us to capture the core area of defining symptoms for both conditions: social interactions. We collected a rich, multimodal dataset of dyads using hyperrecording to capture different communication channels in a time-locked manner (speech, facial expressions, motion). Then, we trained support vector machines to classify between dyad types (ASD-involved, BPD-involved and comparison dyad). Stacking several models containing conceptually related features, our algorithm achieves a near 82% of balanced accuracy, solely based on 20 minutes of conversation. These results show the immense potential of TAD for differential diagnostics: data collection only requires microphones and webcams while feature-extraction is automated, making this approach highly objective, scalable and user-friendly.

## Introduction

Autism diagnostics in adulthood is among the top ten most important topics for autism research^1^. Outpatient clinics for Autism Spectrum Disorder (ASD) have very long waiting times affected by rising referral numbers and by the complex challenge of differential diagnostics ^2^. ASD is a neurodevelopmental disorder which is diagnosed based on alterations in social communication and interaction as well as restricted or repetitive patterns of behaviours leading to significant impairments ^3^. Currently no screening tool or diagnostic tool exists that is recommendable for differential diagnostics ^4^. Clinical reality does not usually entail differentiating between presence of ASD and absence of any diagnosis, on the basis of which current screening and diagnostic tools have been developed, but instead it entails a distinction between different conditions associated with social interaction difficulties. One distinction, especially challenging and thus highly relevant, is the distinction between ASD and Borderline Personality Disorder (BPD), particularly in women ^2,5^. BPD is characterised by “instability of interpersonal relationships, self-image and affects” as well as marked impulsivity ^6^; thus, BPD is a diagnosis which significantly affects social interactions and behaviour, but in a different manner from ASD ^7,8^.

In order to meet the increasing demand for assessment, technology-assisted diagnostics (TAD) could support diagnosticians, make diagnostics more reliable and objective, and free up valuable resources ^9^. Thus, the current study tested the feasibility not only of classifying ASD versus non-clinical controls but also of differential diagnostics between two of the most prominent clinical diagnostic groups with social interaction difficulties, ASD and BPD. To provide a scalable and economic approach which is non-invasive and user-friendly, the developed algorithm is based on data collected with webcams and microphones.

Whether for children or adults, diagnostic assessment for ASD is time- and resource-consuming and requires a trained expert ^3^. In the last years, machine learning has been used to develop algorithms to assist the diagnostic process ^3^. These algorithms though have focused mainly on the classification between autistic individuals and non-clinical controls ^10^. While there are some studies looking into TAD for differential diagnostics, none have investigated personality disorders and ASD. For instance, Demetriou and colleagues ^11^ performed a classification on the basis of questionnaire data and could distinguish between ASD versus social anxiety disorder with an accuracy of 72.3% and with psychosis patients with an accuracy of 69.3%. Although the diagnosis of these conditions is mainly based on behaviour, a behavioural quantification was not included in the study by Demetrious and colleagues ^11^. Furthermore, BPD was not considered in the study by Demetriou and colleagues, even though it is both a particularly relevant and at the same time complex diagnostic differentiation to perform ^5^.

Many approaches to TAD of ASD are based on neuroimaging or genetic data ^10^. While these approaches have been promising, they offer little interpretability and are associated with significant costs ^12^. Furthermore, ASD diagnoses are based on behavioural symptoms and there are no diagnostic biomarkers of ASD yet ^3^; thus, algorithms assisting diagnostics should be based on these behavioural symptoms until reliable biomarkers with increased interpretability and a clear connection to the diagnostic criteria are found. Finally, having an fMRI scan or blood samples taken might not be an option for many individuals with ASD. Hence, we argue for capturing clinically relevant behaviour using scalable, user-friendly and economic technology. Other studies have relied on behavioural data from ASD for machine learning classification, but they did so by relying on input data from questionnaires or semi-structured interviews, such as scores of the Autism Diagnostic Observation Schedule (ADOS) ^3,13^. Here, the downside is the circularity: the algorithm classifies the people based on the same information as the diagnostician.

Social symptoms are at the core of both ASD and BPD and offer a data-rich context to extract features for TAD. Previous work has demonstrated the feasibility of using kinematics ^14–16^, facial expressions ^17^ and speech ^18–20^; however, most studies collected these data from individuals rather than using social interaction where symptoms are most pronounced. In contrast, we have demonstrated in previous studies that quantifying behaviour from live dyadic interactions has been a promising approach for diagnostic classification ^21–24^. Specifically, we computed synchrony, the coordination of behaviour, both intrapersonally, i.e., between communication channels, and interpersonally between interaction partners for different modalities to capture the flow of dyadic interaction ^25^. Reduced interpersonal synchrony on multiple channels ^23,26,27^ as well as reduced intrapersonal synchrony ^28–30^ have been specifically associated with ASD, possibly leading to less favourable impressions of autistic adults and ASD-involved interactions ^31,32^. Interestingly, there is some evidence suggesting BPD might be associated with an *increase* in interpersonal synchrony of motion ^33^. In addition, comparing ASD to a mixed clinical comparison sample drawn from a referral population for autism diagnostics due to social interaction difficulties, interpersonal synchrony turned out to be the sole differentiating marker where questionnaire data performed at chance level ^23^. Thus, interpersonal coordination during reciprocal exchange specifically lends itself as potential objective automatized differential diagnostics of ASD vs. BPD.

In our previous studies comparing ASD to non-clinical controls, support vector machines (SVM) were able to classify between autistic and non-autistic adults based on interpersonal synchrony of speech ^24^ and facial expressions ^22^ as well as intrapersonal synchrony of movement ^21^. The dyadic nature of these interactions is also reflected in the classification labels: we distinguish between interactions which include a person with ASD (ASD-involved dyads) and those that do not. This approach reflects the inherently reciprocal nature of social interactions.

Thus, the goal of the current study was to focus on clinically relevant behaviour from which features were extracted in an automated manner to differentiate between ASD-involved and BPD-involved social interactions using support vector machines. This project is a continuation of Plank *et al*. ^24^ and Koehler *et al*. ^22^, additionally including participants with BPD. Furthermore, another novelty of the current approach is that we go beyond measuring single modalities. To reflect the complex, concerted coordination happening between interaction partners during the dynamics of live interactions, we included cross-modal features, allowing us to capture the non-verbal reaction of one interaction partner towards speech of the other interaction partner. For instance, one might smile in reaction to someone’s joke or nod to acknowledge their gestures. We used webcam recordings of body movements and facial expressions as well as speech recordings during two ten-minute dyadic conversations between two strangers, one where they discussed their hobbies and one where they collaboratively planned a meal with foods and drinks they both dislike ^26,34^ (see Figure 1). This combination of modalities offers complementary information, allowing us to capture the conversations on multiple behavioural levels ^12^. The data were easy to collect noninvasively, the feature extraction pipeline is automated and the approach avoids circularity. Thus, we argue that this approach has enormous potential for TAD of ASD.

**Figure 1.**
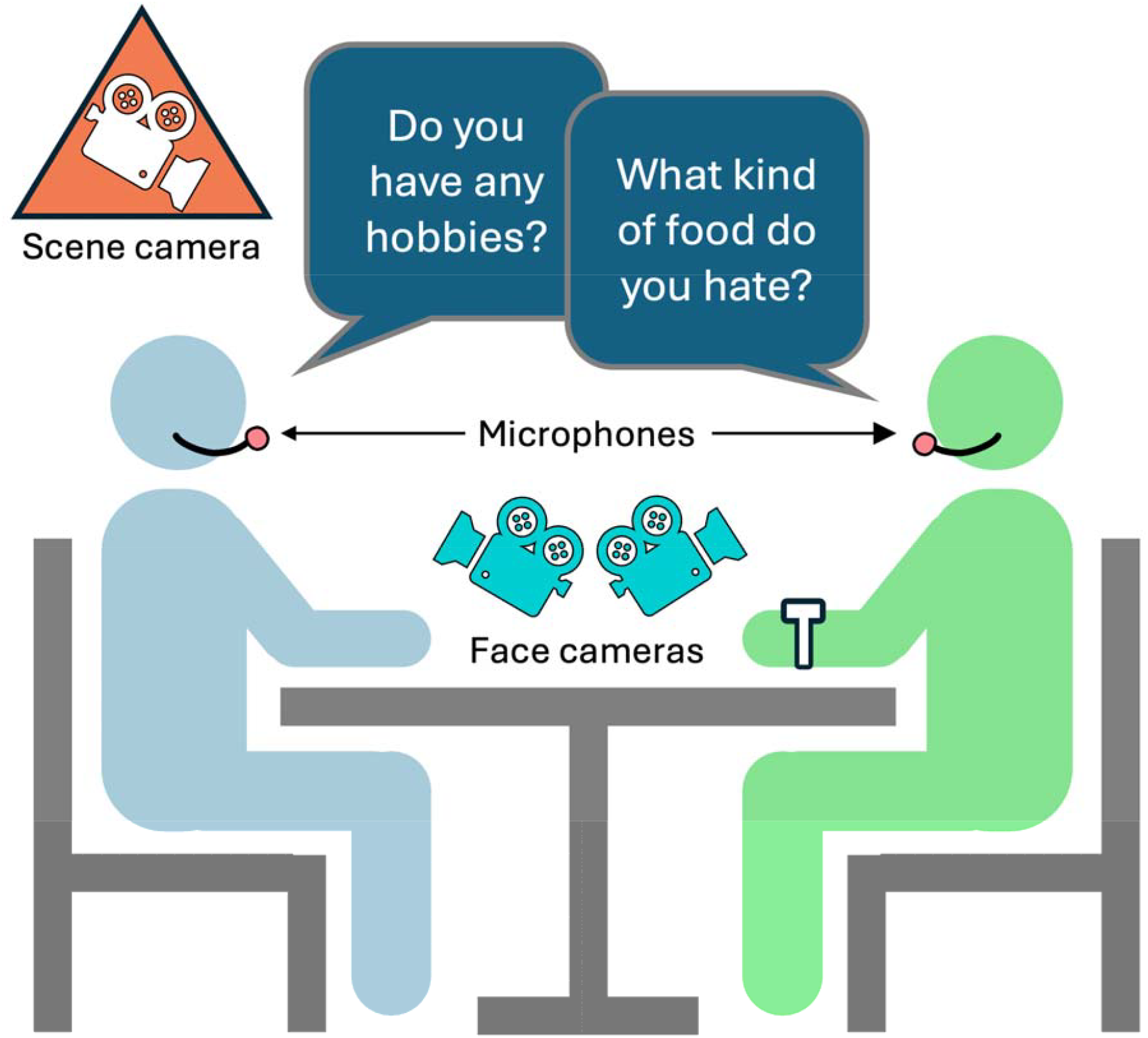
Participants engaged in dyadic conversations with strangers which were audio and video recorded. They were given a prompt for each of two ten-minute conversations: their hobbies or to plan a meal with foods and drinks they both dislike. During the conversations, they were seated at a table approximately 190cm apart and asked not to move their chairs. In front of them, we placed two webcams on tripods with which we video recorded their facial expressions (pictured in turquoise). We also placed another webcam at a 240cm distance from the table to capture the whole scene from a side angle (pictured in orange). Furthermore, we captured their speech using shotgun microphones (pictured in coral). Participants were also wearing wristbands to capture heart rate and electrodermal activity on their left hand (pictured in white on the right participant), which will be reported elsewhere.

## Results

We used support vector machines to classify between three types of interaction contexts: ASD-involved, BPD-involved and comparison dyads (COMP). Eight base models were created, each focusing on one type of feature. Five of these base models contain synchrony features: the *FACEsync* model contains features of interpersonal synchrony of facial expressions, the *HEADsync* and the *BODYsyn*c model contain features of interpersonal synchrony of head and body movements, respectively, and the *CROSSsync* captures interpersonal synchrony of crossed head and body movements, i.e., the head movements of one person influencing the body movements of the other. The *INTRAsync* model contains features of intrapersonal synchrony between head and body movements of the same interaction partner. Furthermore, the *MovEx* model contains features assessing total movement and expressiveness of the face. All features extracted from the audio data are included in the Speech model, including turn-taking features, interpersonal synchrony and individual speech features. Lastly, the *CROSSturn* model includes features that capture movement and facial expressions while someone is speaking or listening to their interaction partner speak. All base models were combined in a stacking model to leverage information from all influential features for the classification. We used permutation testing and compared the base models’ performance to the stacking model. All *p-*values are corrected using the false discovery rate (FDR), except for posthoc group comparisons which are corrected using the Tukey method ^35,36^.

### Distinguishing ASD-involved from BPD-involved interactions

Four of our eight base models were able to distinguish between ASD-involved and BPD-involved interactions based on permutation testing, with the exception of the *BODYsyn*c, *CROSSsync, HEADsync* and *INTRAsync* models (*BODYsyn*c: *p*_*FDR*_ = 1; *CROSSsync*: *p*_*FDR*_ = 0.136; *HEADsync*: *p*_*FDR*_ = 0.936; *INTRAsync*: *p*_*FDR*_ = 1; all other models *p*_*FDR*_ < 0.05). Of the base models, the *CROSSturn* model performed best, with a balanced accuracy of 76.7%, weighing sensitivity and specificity equally, which were 67.6% and 85.7%, respectively. The other base models performing above chance were the Speech model with a balanced accuracy of 70.4% (64.7% sensitivity; 76.2% specificity), followed by the *MovEx* model with 68.7% (58.8% sensitivity; 78.6% specificity) and the *FACEsync* model with 68% (76.5% sensitivity; 59.5% specificity). The stacking model outperformed all base models, although the difference between the stacking model and the *CROSSturn* model was not significant (see Table 1). The stacking model achieved a balanced accuracy of 81.7% (70.6% sensitivity; 92.9% specificity; see Figure 2), meaning that only two interaction partners from BPD-involved interactions were misclassified as ASD-involved, and ten interaction partners were misclassified as BPD-involved despite being ASD-involved. The other 64 interaction partners were classified correctly by the stacking model (see Figure 3). For performance parameters and FDR-corrected *p*-values for all models, please consult the supplementary materials S5.

**Table 1.**
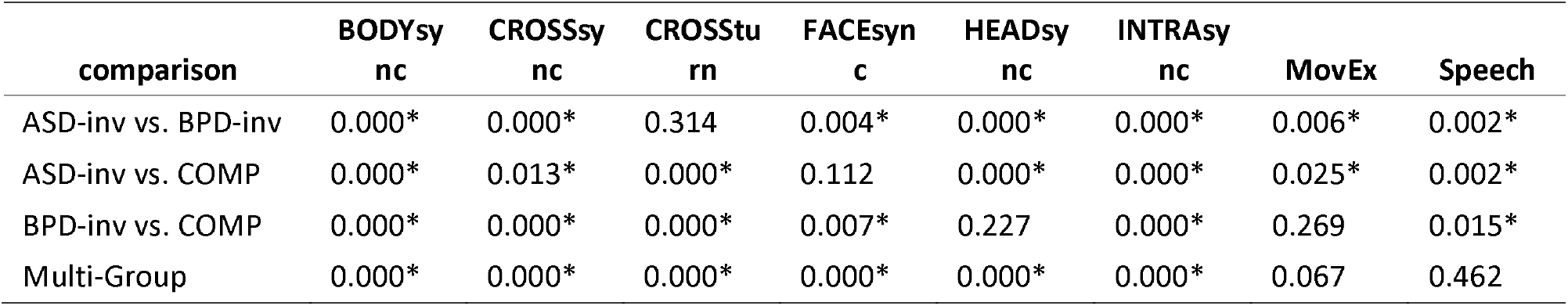
Comparisons of the base model performances with the stacking model performances, denoted by *p*_*FDR*_. Significant differences are highlighted with asterisks. COMP = comparison interaction context.

**Figure 2.**
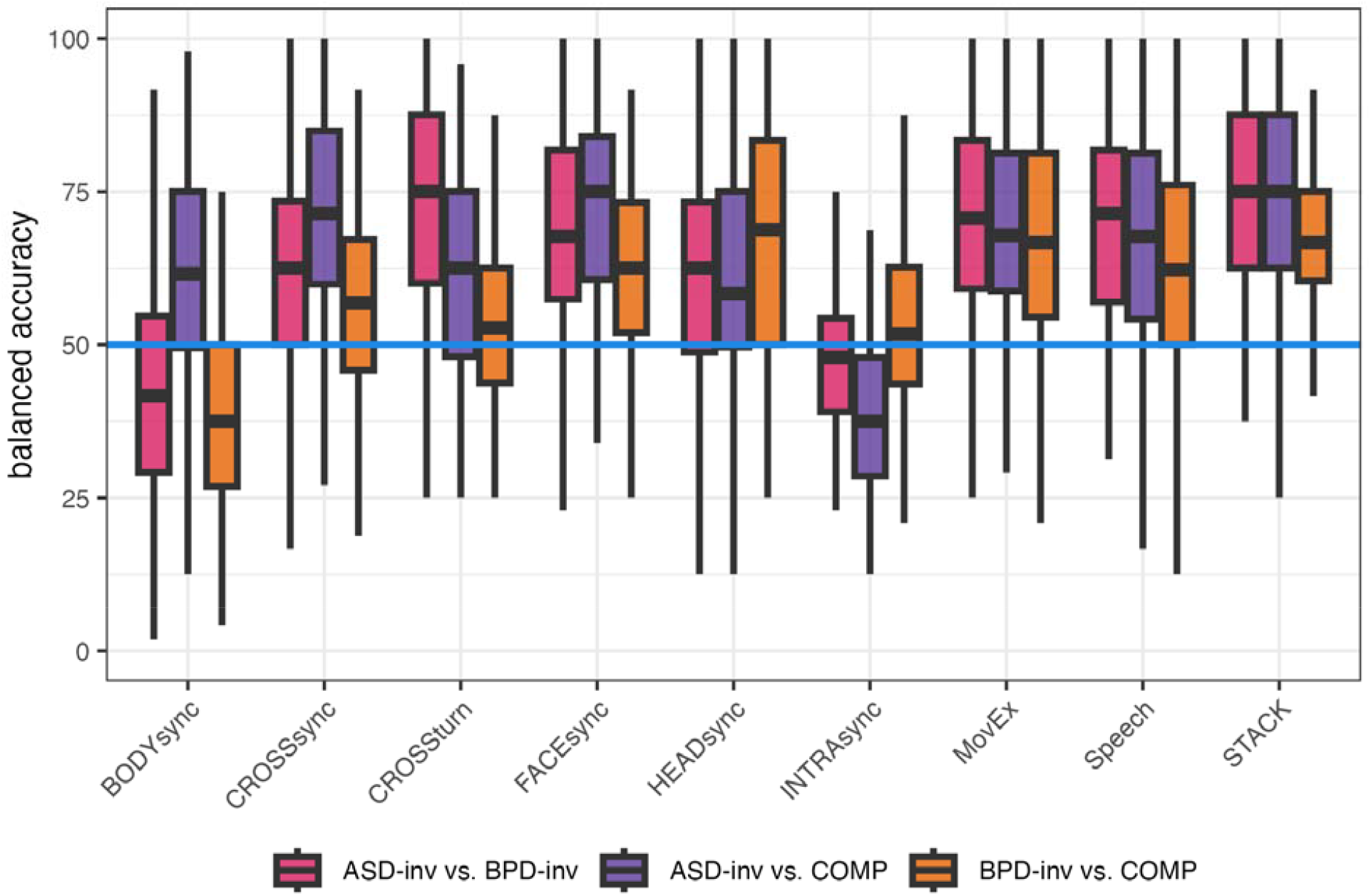
Performance of the base models and the stacking models for the One-vs.-One comparisons. Interaction contexts are classified as BPD-involved, ASD-involved or COMP. We plot the best performance value for each of the ten permutations of the ten folds of the outer loop of the cross-validation structure. Performance is measured in balanced accuracy which weighs sensitivity and specificity equally. The median performance is plotted as a stronger line within the boxes. Hinges capture the interquartile range while whiskers reach 1.5 of the interquartile range from the respective hinges.

**Figure 3.**
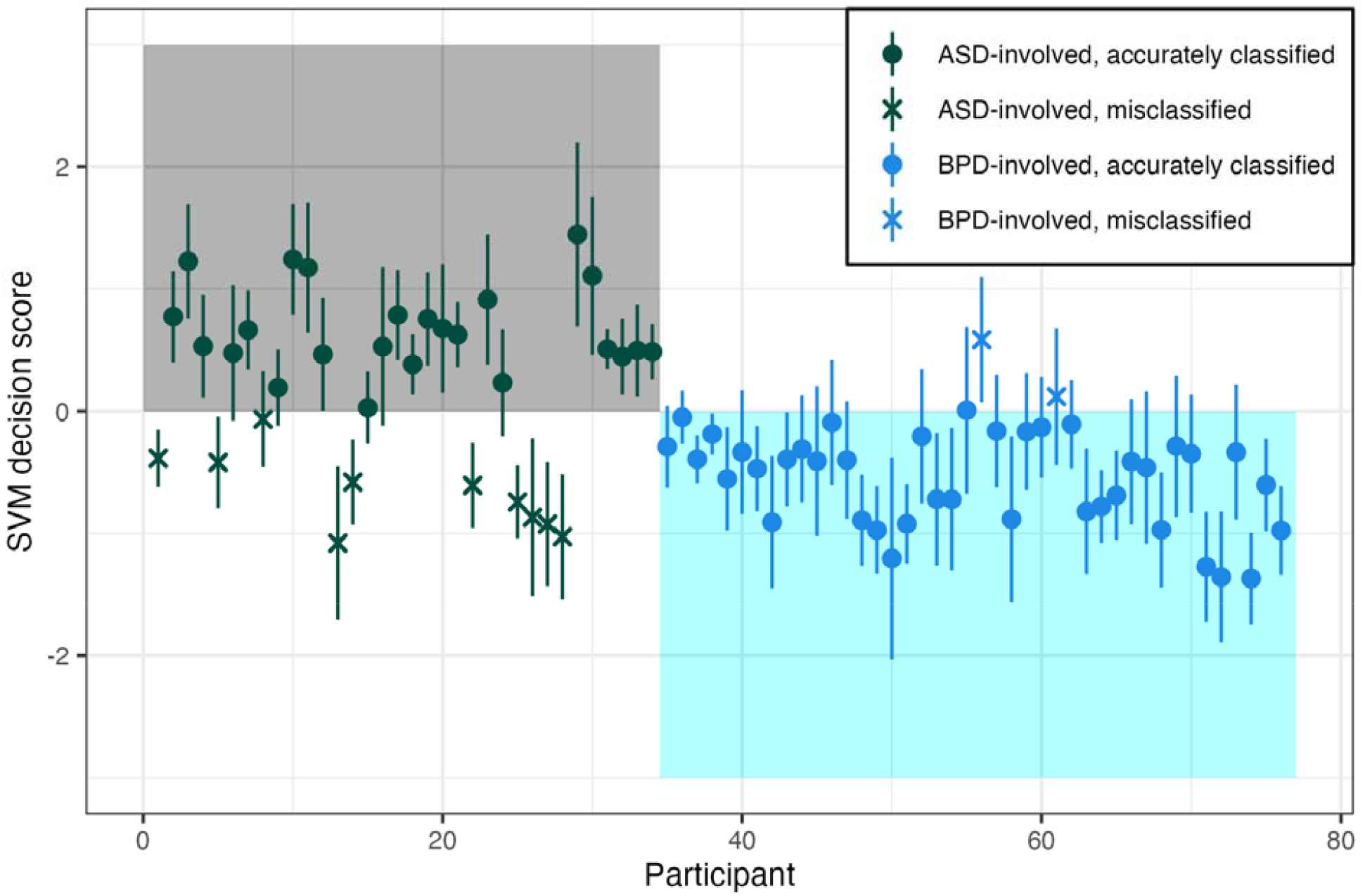
Decision scores from the stacking model classifying between interaction partners from BPD-and ASD-involved interactions. Colour denotes the real labels, i.e., whether a person was taking part in BPD-involved or ASD-involved interactions. The shape shows whether this person was accurately classified with the shading showing the area of accurate classifications for both interaction contexts. Almost all the interaction partners from BPD-involved interactions were accurately classified, with only two exceptions. Most interaction partners from ASD-involved interactions were accurately classified as well, with ten interaction partners being mistakenly classified as having been part of a BPD-involved interaction.

### Distinguishing ASD-involved from comparison interactions

All base models except the *HEADsync* and the *INTRAsync* models performed significantly better than chance according to permutation tests (*HEADsync*: *p*_*FDR*_ = 1; *INTRAsync*: *p*_*FDR*_ = 1). For this comparison, the *FACEsync* model achieved the highest balanced accuracy, specifically 74.9% (79.4% sensitivity; 70.5% specificity), closely followed by the *CROSSsync* model with 71.3% balanced accuracy (67.6% sensitivity; 75% specificity). The stacking model had a significantly higher balanced accuracy of 80.6% (79.4% sensitivity; 81.8% specificity). Here, seven ASD-involved interaction partners were misclassified as COMP, while eight COMP interaction partners were mistaken as ASD-involved. The rest of the interaction partners, 63 in total, were classified accurately. The stacking did not perform significantly better than the best base model (*FACEsync*: *p*_*FDR*_ = 0.112) but outperformed all other base models (see Table 1).

### Exploring influential features

To explore the features as expressed by the interaction partners, we plotted the three most influential features for the distinction between ASD-involved and BPD-involved as well as for the distinction between ASD-involved and COMP interactions. We focused on the *CROSSturn, FACEsync, MovEx* and Speech models and determined the most influential features based on their feature weights. For the Speech model, we focused on individual features. Visual exploration of these features revealed COMP interaction partners in the ASD- or BPD-involved dyads adapted their behaviour to their clinical counterpart in some of the features (see Figure 4). For instance, group comparisons revealed that COMP interaction partners did not differ from their clinical interaction partners in the variance of their speech intensity in the hobbies conversation (ASD-involved: *p*_*HSD*_ = 0.711; BPD-involved: *p*_*HSD*_ = 0.999), but COMP interaction partners’ speech variance differed between COMP interaction partners in BPD-involved and COMP interaction partners in ASD-involved interactions (*p*_*HSD*_ < 0.05). Furthermore, COMP interaction partners in COMP dyads behaved comparably to COMP interaction partners in both clinical interactions (ASD-involved: *p*_*HSD*_ = 0.2; BPD-involved: *p*_*HSD*_ = 0.203). A similar pattern can be observed for the standard deviation of synchronisation of AU26, which measures jaw dropping, in the hobbies conversation. COMP interaction partners in clinical interactions did not differ from their interaction partners (ASD-involved: *p*_*HSD*_ = 0.999; BPD-involved: *p*_*HSD*_ = 0.936), but COMP interaction partners in BPD-involved interactions differed from COMP interaction partners in ASD-involved interactions (*p*_*HSD*_ < 0.05). COMP interaction partners also showed comparable total head motion in the hobbies conversation to their interaction partners (ASD-involved: *p*_*HSD*_ = 1; BPD-involved: *p*_*HSD*_ = 0.151), but here they also did not differ between clinical interactions (*p*_*HSD*_ = 0.57). However, COMP interaction partners in COMP dyads exhibited more head motion than COMP interaction partners in ASD-involved interactions (*p*_*HSD*_ = 0.04), but not from COMP interaction partners in BPD-involved interactions (*p*_*HSD*_ = 0.726). For visualisations of influential features in other comparisons, see S6.

**Figure 4.**
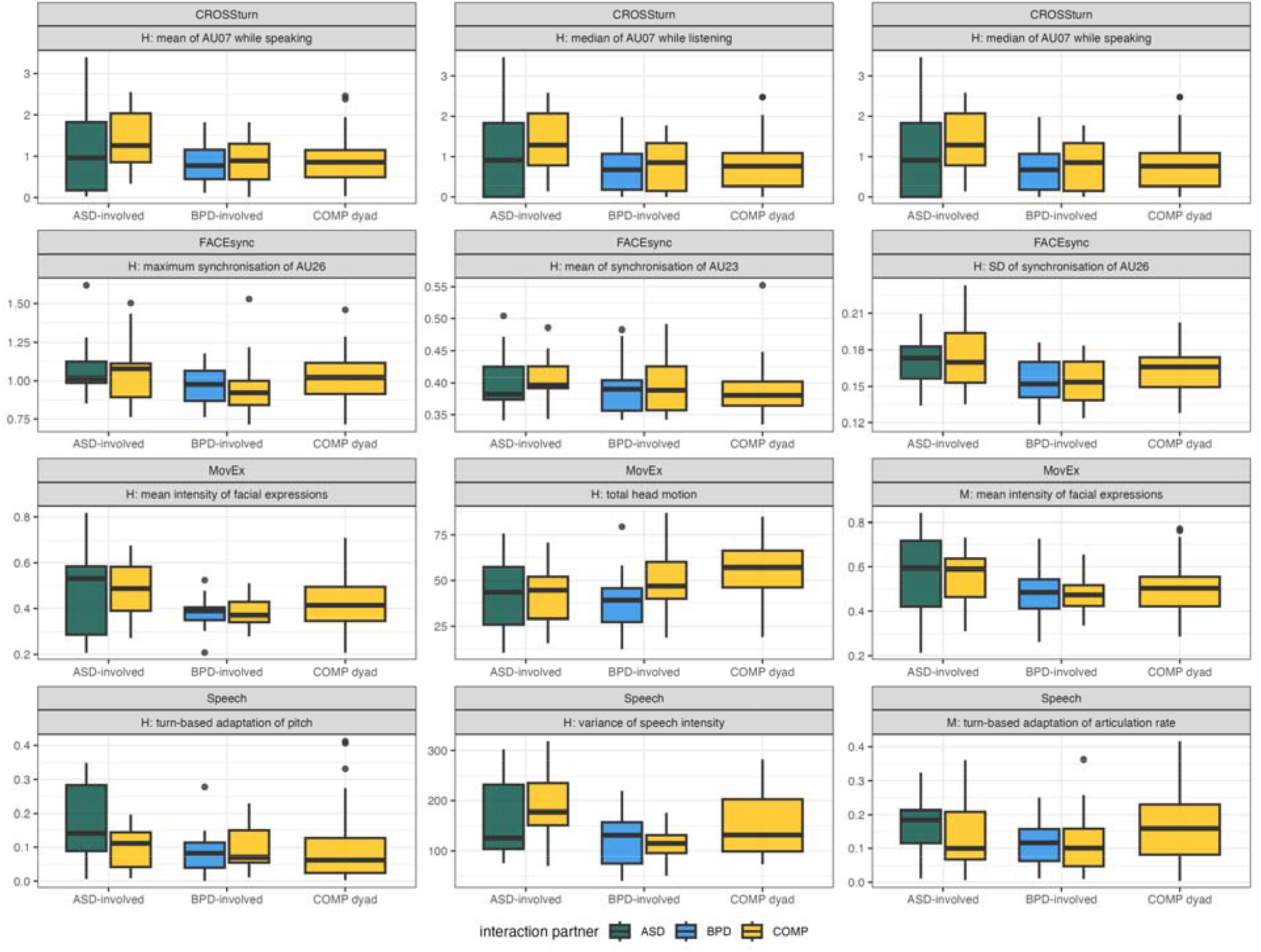
Box plots comparing interaction partners between and within interaction contexts. We focus here on the most influential features from base models with the highest balanced accuracy when classifying between interaction partners from BPD- or ASD-involved interactions. Comparison adults without any psychiatric diagnoses (COMP) seem to adjust their behaviour to their BPD or ASD interaction partner for some features, for instance the variance of their speech intensity. AU = facial action unit (AU07 = lid tightener; AU23 = lip tightener; AU26 = jaw drop); H = hobbies conversation; M = mealplanning conversation; SD = standard deviation.

### Exploring multi-group classification

Arguably, the above results already show that distinguishing between the two clinical labels involves a different pattern of models compared to distinguishing between the ASD-involved and the COMP interactions, which also reflects clinical decision making (a clinical decision between clinical labels is derived on different information than the decision between the presence and absence of a clinical label). Nevertheless, we also explored the use of a multi-group classification which uses the output of the three one-vs-one classifiers to predict one of the three possible labels: ASD-involved, BPD-involved or COMP interactions. All base models performed significantly above chance except for the *BODYsyn*c model (*p*_*FDR*_ = 1). The best base model was the Speech model which achieved a balanced accuracy of 63.9%, followed by the *MovEx* model which reached 60.8% balanced accuracy. The stacking model performed comparable to those two base models with 63.2% balanced accuracy, while outperforming all other base models (see Table 1). The confusion matrix of the stacking model revealed that many of BPD-involved and COMP interactions were misclassified as ASD-involved, 47.6% and 40.9% respectively, while ASD-involved interactions were detected accurately in 61.8% of the cases, possibly reflecting our focus on ASD associated features.

## Discussion

Theory-driven selected features capturing clinically relevant behaviour showed an enormous potential of TAD for differential diagnostics of ASD. A stacking support vector machine leveraging decision scores from base models including information from video and audio recordings of dyadic conversations distinguished between interaction partners from ASD- and BPD-involved interactions with approximately 82% balanced accuracy. This model only misclassified twelve of 76 interaction partners.

At the same time a stacking model classifying between interaction partners from ASD-involved and COMP dyads reached a balanced accuracy of 80%. These features were specifically chosen to capture social interaction behaviour associated with ASD, including multiple measures of behavioural synchrony, demonstrating the efficacy of theory-driven feature selection to develop algorithms for TAD.

Most importantly, the algorithms tackle the so-far overlooked but highly relevant differential diagnostics for autistic adults and potentially adolescents. In the clinical setting, the diagnostic decision, particularly in adolescence and adulthood, is usually not one of absence/presence of ASD but the clinical decision between ASD and differential diagnoses, such as BPD. One of the most common differential diagnoses for adults indeed are personality disorders ^2^. Thus, the focus of this project was to evaluate the use of digital technology for assisting differential diagnostics, specifically distinguishing between BPD and ASD, a distinction particularly relevant for late and female diagnostics. Differential diagnostics is a resource- and time-intensive process; however, our algorithm was able to differentiate between those two diagnoses with high precision based only on two ten-minute conversations. Thus, our algorithm could be a valuable tool for clinicians assessing ASD versus differential diagnoses in adults, freeing up resources, shortening waiting times and reducing suffering.

An important advantage of our approach is its scalability and efficiency. The presently used tools, webcams and microphones, are cheap and user-friendly. Feature extraction algorithms are mainly automated, requiring little working power. While some projects used semi-social interactions in virtual reality ^37^ or simulated virtual interactions ^38^, most projects developing TAD have focused on neuroimaging which is not only expensive to acquire and analyse, but also excludes individuals not able to lie still in the scanner or to handle the noise ^10^. As sensitivity to noise is common in autistic individuals, autistic people might be less likely to sign up for diagnostic evaluation that includes neuroimaging ^39,40^. Especially people in rural areas or the global south may not have easy access to these methods, to a certain extent limiting these approaches to Western, Educated, Industrialized, Rich and Democratic (WEIRD) cultures ^41^. Our approach could be extended to other cultures without the need of expensive equipment. While some features are certainly going to differ in their expression between cultures and languages, the same process could be applied to collect data, extract features and develop machine learning algorithms. We successfully demonstrated this potential in a proof-of-concept study focusing on classifying autism in a South Korean sample ^42^.

While most base models focusing on conceptually relevant features performed above chance when distinguishing between interaction partners from ASD- and BPD-involved interactions, the balanced accuracy of those models varied, ranging from 40% for the model focusing on synchronisation of body movements to 77% in the model focusing on movement and facial expressiveness while talking or listening. While some of the base models were comparably powerful when distinguishing between ASD- and BPD-involved or between ASD-involved or COMP interactions, there were some differences. The model focusing on synchronisation of body movements only successfully distinguished between ASD-involved and COMP interactions, possibly indicating that adults with ASD and BPD are similar in this regard. Similarly, the model including features capturing synchronisation of head with body movements and vice versa was able to distinguish between both clinical labels and the comparison label, but not between ASD- and BPD-involved interactions. Other models were able to distinguish between all combinations, in particular: the model focusing on speech and turn-taking features, the model focusing on synchronisation of facial expressions as well as the model focusing on facial expressiveness and movement while talking or listening. Thus, these models seem to be especially promising for differential diagnostics and could be assessed with interaction contexts including people with other psychiatric diagnoses than BPD or ASD.

Although the dyadic approach we chose comes with challenges, both for constructing the cross-validation and for the interpretation, we chose it to be able to assess social interactions. Simply classifying between ASD and non-clinical controls would disregard the mutual influence the two interaction partners pose on each other’s behaviour. Instead, only a dyadic approach can fully capture the reciprocal nature of social interactions, the core of ASD symptoms. However, a dyadic approach comes with specific challenges for analysis. For example, since the interaction partners are influencing each other’s behaviour, one must ensure that the model does not merely learn who was in a dyad with whom. Thus, we carefully designed a cross-validation structure where interaction partners belonging together are always in the same fold, both for the outer and the inner loop, to reflect their dependence. We used the same cross-validation structure for all models and included this constraint for our permutation testing. Therefore, we can rule out that the model merely learned dyad membership. We argue that it is worthwhile considering and incorporating a dyad structure, because dyadic conversations provide researchers with insights into the dynamic nature of social interactions that cannot be accessed on an individual basis.

We selected features for our base models in a theory-driven way, focusing on aspects of social interaction behaviour for which reliable differences between autistic and non-autistic people have been documented. Furthermore, the conversation prompts were also chosen with ASD in mind: autistic people often monologue about their hobbies but struggle with conversations requiring higher levels of reciprocity with smooth turn-taking which is important during collaborative planning ^43,44^. This autism-focused feature selection process could explain why the comparison between BPD-involved and COMP interactions did not perform as well as the other models, although the stacking model still reached a respectable balanced accuracy of 65% showing great promise. Selecting interaction contexts to elicit social interaction behaviour implicated in BPD would probably increase the performance, although this was not the goal of the current study in which differential diagnostics of ASD was the focus and BPD served as the clinical comparison group. If future studies were to consider developing further diagnostic algorithms specifically to classify BPD, the following considerations might be helpful. BPD is associated with differences in trust behaviour and hypersensitivity to social rejection ^45^. Furthermore, people with BPD are prone to misinterpreting neutral situations, experiencing feelings of social rejection in typical inclusion scenarios and demonstrating difficulties in repairing cooperative efforts following experiences of disappointment ^8^. These differences could be leveraged by including more competitive conversation prompts or possibly extending the conversations to include more than two people, thus, increasing the possibility of misinterpreting neutrality for social rejection.

As such the approach presented in the current study shows a big potential for developing objective, scalable and efficient tools for TAD in a variety of populations, although tailoring the interactions in a theory-driven way would be warranted, as performed here for the purpose of ASD diagnostics. Concerning ASD, the algorithms developed in the current study only apply to speaking individuals. While a significant portion of autistic people is non-speaking ^46^, diagnostics of non-speaking autistic people arguably is less in need of TAD because being non-speaking is a strong and clear symptom which initiates the diagnostic process already during childhood. A similar case can be made for autistic people with intellectual disabilities – many are already diagnostically assessed before reaching adolescence or even adulthood. Concerning speaking individuals, the algorithms presented in the current study will need to be validated in a larger dataset and in adolescents or even children. Furthermore, the algorithms should be assessed in comparison to other differential diagnoses to ensure their validity and generalisability.

This study shows the enormous potential of using scalable recording technology to assist differential diagnostics, thereby streamlining the diagnostic process and freeing up resources. The complexity of differential diagnoses, specifically for adults, has resulted in increased waiting times and prolonged suffering. One problem hampering an improvement of the situation is that current screening and diagnostic tools are not recommended for diagnostics in adulthood due to low differential diagnostic specificity ^4^. Thus, developing solutions able to support differential diagnostics that are faster and more objective is urgently needed. Our approach has several important advantages. First, the features capture clinically relevant behaviour, tapping into the context in which symptoms of autism are most pronounced: social interactions. These interactions are captured in live dyadic conversations. Second, we pair user-friendly devices, namely webcams and microphones, with automated feature extraction, making our approach highly scalable as there is no need for manual annotation. Third, we compute features not only within the individual and between the interaction partners, but also between modalities. These cross-modal features are highly relevant in real life, where reactions to an interaction partner is rarely confined by modality. Last, data collection can be integrated into the diagnostic process without risking circularity. Thus, we propose this approach for improving the diagnostic process using TAD to support differential diagnostics, ultimately alleviating suffering caused by long waiting times.

### Method

Throughout the manuscript, we use a threshold of *α* = 0.05 to assess significance. Data was preprocessed in R *4*.*3*.*0* ^47^ using RStudio ^48^ and MATLAB *R2023b* ^49^ with all machine learning analyses computed using Neurominer *V1*.*3 curufin* ^50^. All scripts and preprocessed, anonymised data are available on https://github.com/IreneSophia/NEVIA_ASD-BPD/tree/main/ML_BPD-ASD, while full SVM models are provided on https://osf.io/y8xbp.

### Participants

This project reused data from a previous project which developed machine learning models to classify between ASD-involved (i.e., one autistic and one individual without psychiatric diagnoses) and comparison (COMP, i.e., two individuals without psychiatric diagnoses) dyads ^22,24^. To extend this development to the differential diagnostics considering BPD, we recruited 41 adults with BPD and 75 adults without any psychiatric diagnoses. All participants were between 18 and 60 years of age at the time of testing and gave their written informed consent. The two projects were positively evaluated by the Ethics committee of the Medical Faculty of the LMU Munich (number 19-702 and 22-170) and adhere to the ethical standards of the Helsinki Declaration of 1975, as revised 2008. From the previous project, we included 17 autistic adults and 45 comparison adults without any psychiatric diagnoses (COMP) which were all participants that were included in previous studies ^22,24^. We only included clinical participants with either a BPD or an ASD diagnosis but excluded anyone who was given both diagnoses (for a detailed summary of inclusion and exclusion criteria see S2). ASD participants were recruited through local autism networks as well as from a clinical database. Confirmation by a full diagnostic report was required for ICD-10 diagnoses of ASD (F84.0 or F84.5), ensuring they were given by a qualified clinical psychologist or psychiatrist according to clinical guidelines ^4,51^. BPD participants were recruited at the inpatient ward of the LMU University Hospital and from dedicated self-help groups. After exclusion of the data of some dyads, which is detailed in Figure 5, we included the data of 17 ASD-involved, 21 BPD-involved and 22 COMP dyads, resulting in data from 120 participants. Participants did not differ in age 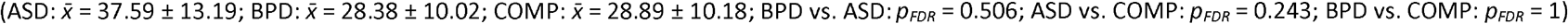, IQ estimate 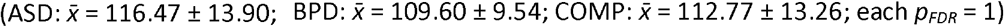 but there was a trend for a difference in gender distribution (ASD: 6 female, 11 male; BPD: 14 female, 7 male; COMP: 52 female, 30 male; *p* = 0.078). However, our exploration of the influence of gender on decision scores in our support vector machines revealed no difference (see S5.4). Testing duration was two to three and a half hours.

**Figure 5.**
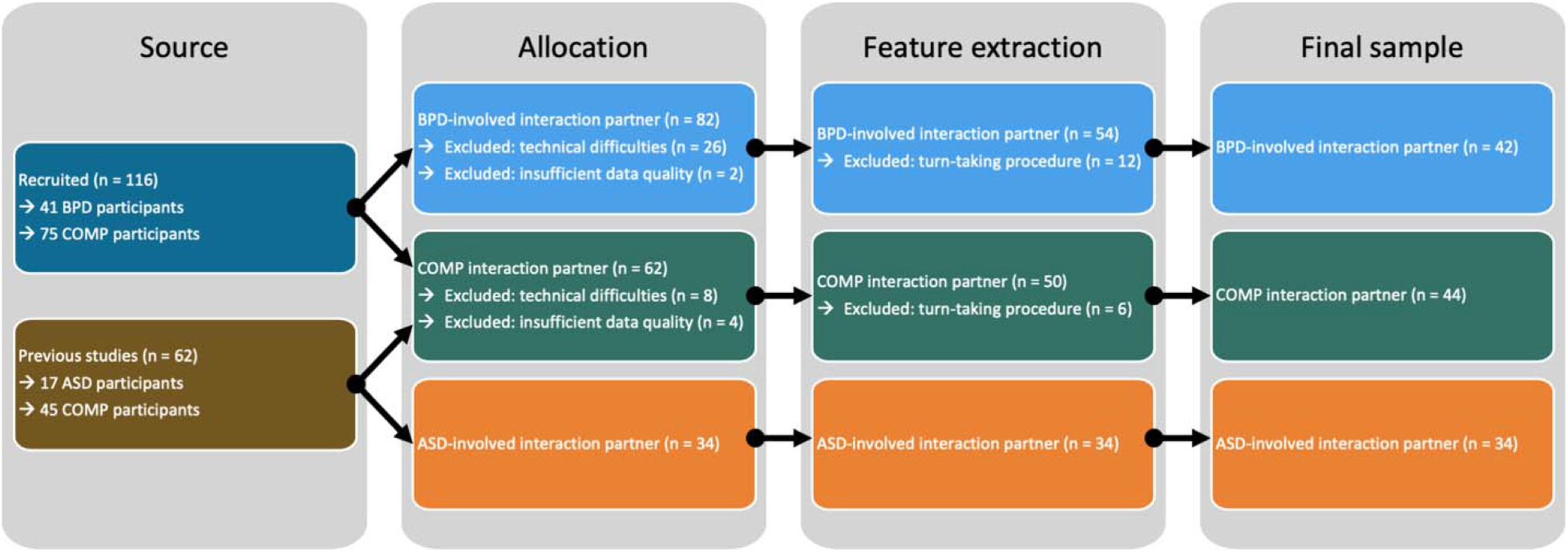
Consort chart detailing the source of the data, exclusion of data and sample sizes for each interaction context. We reused data of ASD and comparison participants without psychiatric diagnoses (COMP) from two previous studies ^22,24^. The data of all participants that were included in the analyses of both of these studies was included. Additionally, we recruited BPD and COMP participants. In a first step, we allocated participants to BPD-involved or COMP dyads based on their availability. Data of dyads where audio or video data of at least one interaction partner was missing was excluded. Due to a technical failure of our audio recording setup which had to be replaced, these numbers are substantial for the BPD-involved interaction contexts. Furthermore, we visually and aurally inspected the data and excluded data from dyads with insufficient data quality, for example, if a microphone picked up both interaction partners. After using the uhm-o-meter ^57,65^ to distinguish between each interaction partner speaking and being silent, we visually inspected the results and excluded data from all dyads where speaking turns were not correctly identified.

### ProcedureSSS

Participants were scheduled based on their availability and were paired in ASD-involved, BPD-involved and COMP dyads with each dyad including at least one comparison person without any psychiatric diagnosis. All participants were unaware of the diagnostic status of their interaction partner, unless they chose to disclose it themselves during testing. They had two eleven-minute conversations in these dyads, one where they were prompted to talk about their hobbies and one where they were asked to plan a meal with foods and drinks they both dislike. The hobbies prompt was chosen to capture monologuing by ASD adults when talking about their interests ^43^, while the mealplanning conversation required increased turn-taking and collaboration from both participants ^26,34^. The order of the conversations was counterbalanced across participants and the experimenter left the room during the conversations. After both conversations were finished, they filled out a custom rapport questionnaire. The conversations were captured with three webcams (Logitech C922, 640×480 pixels, 30Hz; see Figure 1). Webcams were commanded by a PsychoPy ^52^ script to ensure they were time-locked. Participants were fitted with headset microphones (t.bone HeadmiKe, D AKG) connected to the same recorder (Zoom H4N Pro, 44,100 Hz) to capture their speech as separate channels. They also wore Empatica wearables (E4 or EmbracePlus) to collect heart rate and electrodermal activity. These data as well as information from collected blood samples are outside of the scope of this study. All conversations took place in stable artificial light to control for any biases caused by lighting changes ^53^. Since some of the data was collected during the COVID-19 pandemic, we modified the setup to maximise the safety of our participants, for instance by placing a plexiglass between them. Furthermore, participants filled out several questionnaires to capture subjective clinical self-ratings, completed two intelligence assessments which were combined for the IQ estimate and performed the Berlin Emotion Recognition Test (BERT) ^54^. Details and group comparisons are found in the supplementary materials S3.

### Data preprocessing and feature extraction

Audio and video streams were time-locked by aligning them based on the sound and visual of a movie clap denoting the start of the task. Videos and audio tracks were cut to 10min length using DaVinci Resolve. We extracted head position (each three translational and three rotational parameters) and facial expressions as the strength of the activation of action units using OpenFace *2*.*0* ^55^. Furthermore, we used Motion Energy Analysis (MEA) to capture motion quantity for head and body regions from the scene camera ^53^. MEA sums pixel changes from frame to frame in pre-specified regions of interest. Since the lighting is constant, all changes indicate movement, thus creating a time series of motion quantity. Audio data was analysed using a custom processing pipeline in praat ^56^ which incorporated the uhm-o-meter by De Jong and colleagues ^57^. This pipeline extracted individual phonetic and turn-taking features.

We calculated multiple synchrony measures from these extracted features using windowed cross-lagged correlations as implemented in the rMEA package ^58^: interpersonal synchrony between two interaction partners, interpersonal synchronisation focusing on the degree to which one interaction partner was imitating and following their interaction partner as well as intrapersonal synchrony within one interaction partner. For lag, step and window size, we stayed in line with previous literature ^22,24,59–61^. We applied Fisher’s Z-transformation and converted all values to their absolute ^26,62^. Furthermore, we computed turn-based adaptation of pitch and intensity. For details on the feature extraction and transformation, please consult the supplementary materials S4.

### Support vector machines for classification

We used support vector machines to leverage our features to distinguish interaction partners from two interaction pairings at a time, resulting in three comparisons: ASD-involved versus BPD-involved, ASD-involved versus COMP interactions as well as BPD-involved versus COMP interactions. Furthermore, we explored multi-group classification by assigning each interaction partner one label which was determined using the Hamming distance. We conceptually grouped features for the separate base models. *INTRAsync* unites all features capturing intrapersonal synchrony within one interaction partner. *BODYsyn*c, *HEADsync* and *FACEsync* focus on interpersonal synchronisation of the motion quantity of the body, the head and of the strength of activation of facial action units, respectively. *CROSSsync* includes features measuring synchronisation of motion quantity of the body of one interaction partner with the head of the other. *MovEx* contains features describing movement quantity and facial expressiveness of each single interaction partner. Speech includes all features describing speech patterns and turn-taking features extracted from the audio recordings. Last, *CROSSturn* captures cross-modal interactional behaviour, specifically facial expressiveness and motion quantity while the interaction partner is listening or speaking. All models contain features of both conversations, mealplanning and hobbies, and a complete list of features can be found in S4.6. The decision scores of all base models were used in a stacking model to leverage information from all modalities ^63^.

The *Speech* model mirrors the model used for classification between interaction partners in ASD-involved and COMP interactions in Plank *et al*. ^24^. Similarly, five of our eight base models, namely *BODYsync*, *FACEsync, HEADsync, INTRAsync* and *MovEx*, as well as our stacking model mirror the models in Koehler *et al*. ^22^. All models were performed with the same preprocessing and classification settings as in the previous studies unless specified here.

We used a repeated, nested, stratified cross-validation structure where the inner loop allows for hyper-parameter tuning to be separate from the model evaluation in the outer loop. We implemented ten folds with ten permutations in the outer loop and nine folds with one permutation in the inner loop. Due to the dyadic nature of our data, we specifically constructed the cross-validation structure such that both interaction partners of one dyad are always in the same fold. All models were preprocessed and analysed separately. Preprocessing for the base models included scaling the data from −1 to 1 and pruning non-informative features. Here, we slightly deviate from Koehler *et al*. ^22^ who scaled from 0 to 1 to stay in line with Plank *et al*. ^24^ and the NeuroMiner recommendation. Additionally, for all base models with more than 50 features, we performed a principal component analysis, retaining the principal components explaining at least 80% of the variance, before applying pruning again. Base models were trained using LIBLINEAR Support Vector L2-regularised L2-loss classification algorithms with balanced accuracy as the performance criterion and using hyperplane weighing for uneven group sizes. Furthermore, we used ensemble learning to choose the models performing in the top 50%, which deviates from Plank *et al*. ^24^ but is in line with Koehler *et al*. ^22^. The features of the stacking model, which are the decision scores of the base models, were winsorised using a threshold of ±3 standard deviations, and no other preprocessing was performed. Again, we use balanced accuracy as the performance measure and hyperplane weighing, but we switch to a L1-loss LIBSVM algorithm with a Gaussian kernel and use single node learning.

We assessed whether our base models performed above chance with a permutation-based approach where we created 1,000 permutations of the models with shuffled labels. To preserve the dyadic structure of the data, we adjusted the NeuroMiner code, similarly to previous studies ^22,24^. Furthermore, we compared our models based on the best balanced accuracies of each of the ten permutations in the ten folds of the outer loop using a Quade test ^64^.

## Data Availability

All scripts and preprocessed, anonymised data are available online.

https://github.com/IreneSophia/NEVIA_ASD-BPD/tree/main/ML_BPD-ASD

## Acknowledgments

We want to acknowledge all the people helping us in bringing this project to fruition. First, the interns and student assistants of the NEVIA lab who assisted us in preprocessing and inspecting the data: A. Holy, A. Mingione, E. Sangaran, J. Clarke, L. Naumer, P. Radusi and S. Herke. Second, we want to thank F. Cangemi for advising us on the audio recording devices. Third, we want to acknowledge our assistants during the recruitment and data collection, A. Bohn, J. Klieber, K. Krasniqi and L. König. Last, we are grateful to the medical personnel of the LMU University Hospital who helped us recruit our clinical sample, particularly M. Reinhard, as well as to our participants without whom this project would not have been possible. This project was funded by the “Förderprogramm für Forschung und Lehre (FöFoLe) der medizinischen Fakultät der LMU München” which funded JE’s doctoral project (14/2021), by the Stiftung Irene which funded JK’s PhD as well as by the German Research Council (grant numbers 876/3-1 and FA 876/5-1, awarded to CFW).

## Author contributions

Conceptualisation – CFW, ISP, RM

Data curation – JCK, ISP

Formal analysis – ISP

Funding acquisition – CFW, RM

Investigation – AMB, JCK, JE, ISP

Methodology – JCK, ISP, NK

Project administration – AMB, CFW, JCK, ISP

Resources – CFW

Software – JCK, ISP, NK

Supervision – CFW, ISP

Validation – None Visualisation – ISP

Writing – original draft – ISP

Writing review & edition – all authors

