## Supplementary materials for "Highly scalable technology-assisted differential diagnostics of ASD"

I S Plank et al.

2025-10-16

**S7 References** 32

#### S1 Version numbers for R and all packages

```
## [1] "R version 4.5.1 (2025-06-13)"
```

```
## [1] "tidyverse version 2.0.0"
## [1] "knitr version 1.50"
## [1] "ggpubr version 0.6.1"
## [1] "ggrain version 0.0.4"
## [1] "Hmisc version 5.2.3"
## [1] "rstatix version 0.7.2"
## [1] "emmeans version 1.11.2"
## [1] "flextable version 0.9.9"
## [1] "officer version 0.6.10"
## [1] "kableExtra version 1.4.0"
## [1] "english version 1.2.6"
```

### S2 Inclusion and exclusion criteria

Inclusion criteria applying to all participants:

- no neurological diagnoses
- at least 18 and at most 60 years of age
- normal or corrected-to-normal vision
- legally binding, written informed consent from the participant for study participation

Additional inclusion criteria for ASD participants:

- valid F84 diagnosis according to ICD-10
- normal speaking abilities

Additional inclusion criteria for BPD participants:

- valid F60.31 diagnosis according to ICD-10
- no F84 diagnosis according to ICD-10
- IQ estimate above 70

Additional inclusion criteria for comparison participants:

- IQ estimate above 70
- no psychiatric diagnoses
- no current psychotropic medication

Furthermore, we applied the following exclusion criteria:

- underage people or people older than 60 years
- inability to speak
- current or previous neurological disorder
- acute suicidality, self-harming or aggressive behaviour
- lack of legally binding informed consent

Testing was discontinued if participants withdrew their consent, exclusion criteria applied or in the case of technical difficulties.

### S3 Measures

We collected the following self-report questionnaires:

- Autism-like traits: Autism Quotient, AQ[1]
- Empathy: Saarbrücker Persönlichkeitsfragebogen, SPF[2], which is the German version of the interpersonal reactivity index
- Alexithymia: Toronto Alexithymia Scale, TAS20[3]
- Depressiveness: Beck Depression Inventory, BDI[4]
- Self-monitoring: Self-monitoring Scale, SMS[5]
- Movement difficulties: Adult Dyspraxia Checklist, ADC[6]

For all participants recruited for this project, we also collected the short version of the Borderline Symptom List, BSL-23[7].

For 37 of the dyads, we used a plexiglass screen placed between the interaction partners on the table to decrease the chances of spreading an undetected infection. We applied a transparent anti-reflection foil to reduce any mirroring effects. During the conversation, participants took off their face masks. We asked these participants how much the plexiglass influenced them during the interactions (plexi; scale from 0 to 3). All participants were asked how much the cameras influenced their behaviour (video; scale from 0 to 3). Rapport is a sum of the following ratings, all on scales from 0 to 3, thus ranging from 0 to 15:

- How likeable was your interaction partner?
- How friendly was your interaction partner?
- How comfortable did you feel during the conversation?
- How smooth was the communication between you and your conversation partner?
- How well did your conversation partner respond to you?

#### S3.1 Group comparisons based on diagnostic status

```
##        ilabel
## gender ASD BPD COMP
```

Table S1: Group comparisons

| measurement | ASD | BPD | COMP | BPD vs. ASD | COMP vs. ASD | COMP vs. BPD |
|---|---|---|---|---|---|---|
| ADC_total | 49.06 (±17.17), n = 17 | 76.95 (±14.98), n = 21 | 34.99 (±23.86), n = 82 | 0.006* | 0.243 | 0.000* |
| AQ_total | 34.29 (±6.18), n = 17 | 23.67 (±4.72), n = 21 | 14.57 (±5.27), n = 82 | 0.000* | 0.000* | 0.000* |
| BDI_total | 15.94 (±11.57), n = 17 | 24.81 (±10.48), n = 21 | 3.99 (±3.79), n = 82 | 0.715 | 0.000* | 0.000* |
| BERT.acc | 0.80 (±0.07), n = 17 | 0.82 (±0.09), n = 21 | 0.83 (±0.07), n = 82 | 1.000 | 0.919 | 1.000 |
| BERT.rt | 5.79 (±2.78), n = 17 | 3.84 (±1.24), n = 21 | 3.33 (±1.33), n = 82 | 0.453 | 0.000* | 0.637 |
| BSL_total | NaN (±NA), n = 0 | 46.71 (±21.44), n = 21 | 7.19 (±6.84), n = 37 | NA | NA | 0.000* |
| IQ.estimate | 116.47 (±13.90), n = 17 | 109.60 (±9.54), n = 20 | 112.77 (±13.26), n = 82 | 1.000 | 1.000 | 1.000 |
| SMS_total | 5.47 (±2.81), n = 17 | 11.67 (±2.11), n = 21 | 9.80 (±2.62), n = 82 | 0.000* | 0.000* | 0.027* |
| SPF_total | 37.00 (±7.20), n = 17 | 40.95 (±8.36), n = 21 | 44.73 (±5.82), n = 82 | 1.000 | 0.000* | 0.328 |
| TAS_total | 61.76 (±11.03), n = 17 | 55.33 (±11.60), n = 21 | 37.55 (±8.53), n = 82 | 1.000 | 0.000* | 0.000* |
| age | 37.59 (±13.19), n = 17 | 28.38 (±10.02), n = 21 | 28.89 (±10.18), n = 82 | 0.506 | 0.243 | 1.000 |
| plexi | 0.82 (±0.53), n = 17 | 0.85 (±0.90), n = 13 | 0.86 (±0.60), n = 43 | 1.000 | 1.000 | 1.000 |
| rapport | 12.24 (±2.19), n = 17 | 11.95 (±2.64), n = 21 | 12.33 (±2.39), n = 81 | 1.000 | 1.000 | 1.000 |
| video | 0.71 (±0.69), n = 17 | 0.81 (±0.75), n = 21 | 0.63 (±0.68), n = 82 | 1.000 | 1.000 | 1.000 |

```
##    fem   6  14   52
##    mal  11   7   30
```

```
##
##  Pearson's Chi-squared test
##
## data:  tb.gen
## X-squared = 5.1108, df = 2, p-value = 0.07766
```

### S4 Features

#### S4.1 Facial expressions extracted from OpenFace

We only included data of participants with a mean confidence of tracked frames greater than 75% and more than 90% successfully tracked frames. Facial expressions were captured as action units. We did not extract emotional expressions from these facial expressions as coherence between facial expressions and emotions is not a given and might be even less so for autistic people[8].

For the calculation of synchronisation, we included rotational parameters (yaw, roll, pitch) as well as the same action units as in our previous study[9]:

- Mealplanning: 1, 2, 6, 7, 9, 14, 15, 17, 20, 25, 26 and 45
- Hobbies: 1, 2, 6, 7, 9, 15, 17, 20, 23, 25, 26 and 45

We also extracted total facial expressiveness as mean intensity of all action units for each interaction partner to be included in the *MovEx* and the *CROSSturn* models. For the *CROSSturn* model, we also included other action units, as listed below.

These correspond to the following movements:

- AU1: inner brow raiser
- AU2: outer brow raiser
- AU4: brow lowerer (only *CROSSturn*)
- AU5: upper lid raiser (only *CROSSturn*)
- AU6: cheek raiser
- AU7: lid tightener
- AU9: nose wrinkler
- AU10: upper lip raiser (only *CROSSturn*)
- AU12: lip corner puller (only *CROSSturn*)
- AU14: dimpler
- AU15: lip corner depressor
- AU17: chin raiser
- AU20: lip stretcher
- AU23: lip tightener

- AU25: lips part
- AU26: jaw drop
- AU45: blink

Furthermore, we used translational head position parameters to infer head motion using the following formula with $\Delta_t$ referring to the respective frame-to-frame changes:

$$\text{head movement} = \sqrt{\Delta_t x^2 + \Delta_t y^2 + \Delta_t z^2}$$

#### S4.2 Motion quantity extracted using Motion Energy Analysis

This figure shows body (red and purple) and head (yellow and orange) regions of interests of each interaction partner separately:

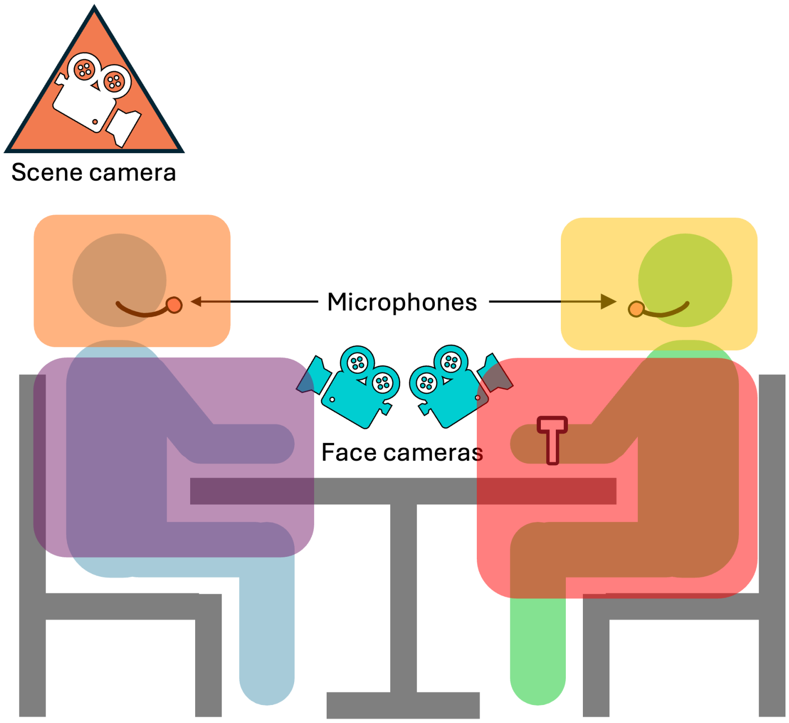

There was always a space between the head and body region. Regions were chosen such that they cover the full range of motion throughout one conversation of one interaction partner. Thus, their sizes differed which is why we scaled all values.

In addition to using the motion quantity to compute synchronisation, we also extracted total movement in each region of interest for each interaction partner to be included in the *MovEx* model.

#### S4.3 Speech and turn-taking features

We extracted pitch using praat's autocorrelation method, a technique widely recognized for its reliability and accuracy[10]. We implemented a two-step pitch extraction method, as outlined by Hirst[11]. First, to capture a broad range of frequencies, we set a low pitch floor of 50 Hz and a high pitch ceiling of 700 Hz, with a time step of 15 ms. All other parameters were set to Praat's default values. Second, using these initial pitch values, we determined the first and third quartiles of pitch for each participant and task. We then used these quartiles to compute individual pitch floors and ceilings with the following algorithm:

$$\text{floor} = \min\left(0.75 \cdot Q_{1,hobbies}, 0.75 \cdot Q_{1,mealplanning}\right)$$

$$\text{ceiling} = \max\left(2.5 \cdot Q_{3,hobbies}, 2.5 \cdot Q_{3,mealplanning}\right)$$

We then used these individual pitch floors (range = 46 to 168Hz, mean = 109.7 ± 31.8) and ceilings (range = 250 to 839Hz, mean = 481.7 ± 147.4) to extract pitch. To ensure an equal number of frames for all participants, we maintained a consistent time step across all analyses. By default, praat calculates this time step using the following formula:

$$\text{timestep} = \frac{0.75}{\text{floor}}$$

Here, we used the same time step of 0.016 as in our previous study[12] which was determined based on the minimum individual pitch floor of that sample. Since the new sample did not include anyone with a lower pitch floor, the time step fits both samples.

Intensity was extracted by convolving the squared sound with a Gaussian analysis window. We used praat's default values of minimum pitch 100Hz and time step of 0.01s.

To estimate synchrony, we extracted continuous pitch and intensity time series for every millisecond of the recording. For pitch extraction, we used consistent parameters across all participants instead of individualized settings. This was necessary because the analysis width depends on the pitch floor. Given the heterogeneity of our sample, we opted for a wide range of considered frequencies, setting the pitch floor at 50Hz and the pitch ceiling at 700Hz. For intensity, we relied on Praat's default values.

In the case of turn-based synchronization, we correlated the median pitch or intensity of each turn with the median pitch or intensity of the preceding turn.

Next, we used the uhm-o-meter[13,14] to differentiate between periods of speaking and silence, identify syllables and extract several prosodic features (total number of syllables, total number of silent phases, duration of speaking as phonation time, speech rate as number of syllables per second, articulation rate as number of syllables per phonation time, average syllable duration and silence-to-turn ratio). The resulting speaking and silent instances were visually and aurally inspected to verify the accuracy of the algorithm.

#### S4.4 Cross-modal features

We captured two types of cross-modal features:

- Interpersonal synchronisation of one person's head with the other person's body movement and vice versa
- AU activation, body and head movement during listening and speaking

#### S4.5 Synchrony computations

We used the following settings for our windowed lagged cross-correlation (WLCC):

Table S2: WLCC settings in seconds

| measure | window | step | lag |
|---|---|---|---|
| Facial action units synchronisation | 7 | 4 | 2 |
| Body MEA synchronisation | 30 | 15 | 5 |
| Head MEA synchronisation | 30 | 15 | 5 |
| Intrapersonal synchrony | 30 | 15 | 5 |
| Pitch synchrony | 16 | 8 | 2 |
| Intensity synchrony | 16 | 8 | 2 |

For each window, the maximum correlation value was chosen out of all relevant lags (peack-picking). We cross-correlated head movements (from OpenFace) with body motion energy time series (from MEA) to estimate intrapersonal synchrony.

#### S4.6 Feature lists

This list shows all features without the information of conversation, i.e., each of these features was added twice to the model, once from the mealplanning and once from the hobbies conversation. The number of features per model is displayed as well. For many of the extracted features, we calculated summary scores some which are indicated by abbreviations (mean, md = median, sd = standard deviation, min = minimum, max = maximum, ske = skewness and kurtosis)

Table S3: List of all mealplanning features of the base models

| model | features |
|---|---|
| **BODYsync (14 features)** | min_M_bodysync, max_M_bodysync, sd_M_bodysync, mean_M_bodysync, md_M_bodysync, skew_M_bodysync, kurtosis_M_bodysync |

| | |
|---|---|
| **CROSSsync (28 features)** | min_M_LOF, min_M_ROF, max_M_LOF, max_M_ROF, md_M_LOF, md_M_ROF, mean_M_LOF, mean_M_ROF, sd_M_LOF, sd_M_ROF, kurtosis_M_LOF, kurtosis_M_ROF, skew_M_LOF, skew_M_ROF |
| **CROSSturn (532 features)** | self_min_M_AU01_r, self_min_M_AU02_r, self_min_M_AU04_r, self_min_M_AU05_r, self_min_M_AU06_r, self_min_M_AU07_r, self_min_M_AU09_r, self_min_M_AU10_r, self_min_M_AU12_r, self_min_M_AU14_r, self_min_M_AU15_r, self_min_M_AU17_r, self_min_M_AU20_r, self_min_M_AU23_r, self_min_M_AU25_r, self_min_M_AU26_r, self_min_M_AU45_r, self_min_M_MEA_body, self_min_M_MEA_head, self_max_M_AU01_r, self_max_M_AU02_r, self_max_M_AU04_r, self_max_M_AU05_r, self_max_M_AU06_r, self_max_M_AU07_r, self_max_M_AU09_r, self_max_M_AU10_r, self_max_M_AU12_r, self_max_M_AU14_r, self_max_M_AU15_r, self_max_M_AU17_r, self_max_M_AU20_r, self_max_M_AU23_r, self_max_M_AU25_r, self_max_M_AU26_r, self_max_M_AU45_r, self_max_M_MEA_body, self_max_M_MEA_head, self_md_M_AU01_r, self_md_M_AU02_r, self_md_M_AU04_r, self_md_M_AU05_r, self_md_M_AU06_r, self_md_M_AU07_r, self_md_M_AU09_r, self_md_M_AU10_r, self_md_M_AU12_r, self_md_M_AU14_r, self_md_M_AU15_r, self_md_M_AU17_r, self_md_M_AU20_r, self_md_M_AU23_r, self_md_M_AU25_r, self_md_M_AU26_r, self_md_M_AU45_r, self_md_M_MEA_body, self_md_M_MEA_head, self_mean_M_AU01_r, self_mean_M_AU02_r, self_mean_M_AU04_r, self_mean_M_AU05_r, self_mean_M_AU06_r, self_mean_M_AU07_r, self_mean_M_AU09_r, self_mean_M_AU10_r, self_mean_M_AU12_r, self_mean_M_AU14_r, self_mean_M_AU15_r, self_mean_M_AU17_r, self_mean_M_AU20_r, self_mean_M_AU23_r, self_mean_M_AU25_r, self_mean_M_AU26_r, self_mean_M_AU45_r, self_mean_M_MEA_body, self_mean_M_MEA_head, self_sd_M_AU01_r, self_sd_M_AU02_r, self_sd_M_AU04_r, self_sd_M_AU05_r, self_sd_M_AU06_r, self_sd_M_AU07_r, self_sd_M_AU09_r, self_sd_M_AU10_r, self_sd_M_AU12_r, self_sd_M_AU14_r, self_sd_M_AU15_r, self_sd_M_AU17_r, self_sd_M_AU20_r, self_sd_M_AU23_r, self_sd_M_AU25_r, self_sd_M_AU26_r, self_sd_M_AU45_r, self_sd_M_MEA_body, self_sd_M_MEA_head, self_kurtosis_M_AU01_r, self_kurtosis_M_AU02_r, self_kurtosis_M_AU04_r, self_kurtosis_M_AU05_r, self_kurtosis_M_AU06_r, self_kurtosis_M_AU07_r, self_kurtosis_M_AU09_r, self_kurtosis_M_AU10_r, self_kurtosis_M_AU12_r, self_kurtosis_M_AU14_r, self_kurtosis_M_AU15_r, self_kurtosis_M_AU17_r, self_kurtosis_M_AU20_r, self_kurtosis_M_AU23_r, self_kurtosis_M_AU25_r, self_kurtosis_M_AU26_r, self_kurtosis_M_AU45_r, self_kurtosis_M_MEA_body, self_kurtosis_M_MEA_head, self_skew_M_AU01_r, self_skew_M_AU02_r, self_skew_M_AU04_r, self_skew_M_AU05_r, self_skew_M_AU06_r, self_skew_M_AU07_r, self_skew_M_AU09_r, self_skew_M_AU10_r, self_skew_M_AU12_r, self_skew_M_AU14_r, self_skew_M_AU15_r, self_skew_M_AU17_r, self_skew_M_AU20_r, self_skew_M_AU23_r, self_skew_M_AU25_r, self_skew_M_AU26_r, self_skew_M_AU45_r, self_skew_M_MEA_body, self_skew_M_MEA_head, other_min_M_AU01_r, other_min_M_AU02_r, other_min_M_AU04_r, other_min_M_AU05_r, other_min_M_AU06_r, other_min_M_AU07_r, other_min_M_AU09_r, other_min_M_AU10_r, other_min_M_AU12_r, other_min_M_AU14_r, other_min_M_AU15_r, other_min_M_AU17_r, other_min_M_AU20_r, other_min_M_AU23_r, other_min_M_AU25_r, other_min_M_AU26_r, other_min_M_AU45_r, other_min_M_MEA_body, other_min_M_MEA_head, other_max_M_AU01_r, other_max_M_AU02_r, other_max_M_AU04_r, other_max_M_AU05_r, other_max_M_AU06_r, other_max_M_AU07_r, other_max_M_AU09_r, other_max_M_AU10_r, other_max_M_AU12_r, other_max_M_AU14_r, other_max_M_AU15_r, other_max_M_AU17_r, other_max_M_AU20_r, other_max_M_AU23_r, other_max_M_AU25_r, other_max_M_AU26_r, other_max_M_AU45_r, other_max_M_MEA_body, other_max_M_MEA_head, other_md_M_AU01_r, other_md_M_AU02_r, other_md_M_AU04_r, other_md_M_AU05_r, other_md_M_AU06_r, other_md_M_AU07_r, other_md_M_AU09_r, other_md_M_AU10_r, other_md_M_AU12_r, other_md_M_AU14_r, other_md_M_AU15_r, other_md_M_AU17_r, other_md_M_AU20_r, other_md_M_AU23_r, other_md_M_AU25_r, other_md_M_AU26_r, other_md_M_AU45_r, other_md_M_MEA_body, other_md_M_MEA_head, other_mean_M_AU01_r, other_mean_M_AU02_r, other_mean_M_AU04_r, other_mean_M_AU05_r, other_mean_M_AU06_r, other_mean_M_AU07_r, other_mean_M_AU09_r, other_mean_M_AU10_r, other_mean_M_AU12_r, other_mean_M_AU14_r, other_mean_M_AU15_r, other_mean_M_AU17_r, other_mean_M_AU20_r, other_mean_M_AU23_r, other_mean_M_AU25_r, other_mean_M_AU26_r, other_mean_M_AU45_r, other_mean_M_MEA_body, other_mean_M_MEA_head, other_sd_M_AU01_r, other_sd_M_AU02_r, other_sd_M_AU04_r, other_sd_M_AU05_r, other_sd_M_AU06_r, other_sd_M_AU07_r, other_sd_M_AU09_r, other_sd_M_AU10_r, other_sd_M_AU12_r, other_sd_M_AU14_r, other_sd_M_AU15_r, other_sd_M_AU17_r, other_sd_M_AU20_r, other_sd_M_AU23_r, other_sd_M_AU25_r, other_sd_M_AU26_r, other_sd_M_AU45_r, other_sd_M_MEA_body, other_sd_M_MEA_head, other_kurtosis_M_AU01_r, other_kurtosis_M_AU02_r, other_kurtosis_M_AU04_r, other_kurtosis_M_AU05_r, other_kurtosis_M_AU06_r, other_kurtosis_M_AU07_r, other_kurtosis_M_AU09_r, other_kurtosis_M_AU10_r, other_kurtosis_M_AU12_r, other_kurtosis_M_AU14_r, other_kurtosis_M_AU15_r, other_kurtosis_M_AU17_r, other_kurtosis_M_AU20_r, other_kurtosis_M_AU23_r, other_kurtosis_M_AU25_r, other_kurtosis_M_AU26_r, other_kurtosis_M_AU45_r, other_kurtosis_M_MEA_body, other_kurtosis_M_MEA_head, other_skew_M_AU01_r, other_skew_M_AU02_r, other_skew_M_AU04_r, other_skew_M_AU05_r, other_skew_M_AU06_r, other_skew_M_AU07_r, other_skew_M_AU09_r, other_skew_M_AU10_r, other_skew_M_AU12_r, other_skew_M_AU14_r, other_skew_M_AU15_r, other_skew_M_AU17_r, other_skew_M_AU20_r, other_skew_M_AU23_r, other_skew_M_AU25_r, other_skew_M_AU26_r, other_skew_M_AU45_r, other_skew_M_MEA_body, other_skew_M_MEA_head |

| | |
|---|---|
| **FACEsync (168 features)** | min_M_AU01_r, max_M_AU01_r, sd_M_AU01_r, mean_M_AU01_r, md_M_AU01_r, skew_M_AU01_r, kurtosis_M_AU01_r, min_M_AU02_r, max_M_AU02_r, sd_M_AU02_r, mean_M_AU02_r, md_M_AU02_r, skew_M_AU02_r, kurtosis_M_AU02_r, min_M_AU06_r, max_M_AU06_r, sd_M_AU06_r, mean_M_AU06_r, md_M_AU06_r, skew_M_AU06_r, kurtosis_M_AU06_r, min_M_AU07_r, max_M_AU07_r, sd_M_AU07_r, mean_M_AU07_r, md_M_AU07_r, skew_M_AU07_r, kurtosis_M_AU07_r, min_M_AU09_r, max_M_AU09_r, sd_M_AU09_r, mean_M_AU09_r, md_M_AU09_r, skew_M_AU09_r, kurtosis_M_AU09_r, min_M_AU14_r, max_M_AU14_r, sd_M_AU14_r, mean_M_AU14_r, md_M_AU14_r, skew_M_AU14_r, kurtosis_M_AU14_r, min_M_AU15_r, max_M_AU15_r, sd_M_AU15_r, mean_M_AU15_r, md_M_AU15_r, skew_M_AU15_r, kurtosis_M_AU15_r, min_M_AU17_r, max_M_AU17_r, sd_M_AU17_r, mean_M_AU17_r, md_M_AU17_r, skew_M_AU17_r, kurtosis_M_AU17_r, min_M_AU20_r, max_M_AU20_r, sd_M_AU20_r, mean_M_AU20_r, md_M_AU20_r, skew_M_AU20_r, kurtosis_M_AU20_r, min_M_AU25_r, max_M_AU25_r, sd_M_AU25_r, mean_M_AU25_r, md_M_AU25_r, skew_M_AU25_r, kurtosis_M_AU25_r, min_M_AU26_r, max_M_AU26_r, sd_M_AU26_r, mean_M_AU26_r, md_M_AU26_r, skew_M_AU26_r, kurtosis_M_AU26_r, min_M_AU45_r, max_M_AU45_r, sd_M_AU45_r, mean_M_AU45_r, md_M_AU45_r, skew_M_AU45_r, kurtosis_M_AU45_r |
| **HEADsync (56 features)** | min_M_headsync, max_M_headsync, sd_M_headsync, mean_M_headsync, md_M_headsync, skew_M_headsync, kurtosis_M_headsync, min_M_pose_Rxsync, max_M_pose_Rxsync, sd_M_pose_Rxsync, mean_M_pose_Rxsync, md_M_pose_Rxsync, skew_M_pose_Rxsync, kurtosis_M_pose_Rxsync, min_M_pose_Rysync, max_M_pose_Rysync, sd_M_pose_Rysync, mean_M_pose_Rysync, md_M_pose_Rysync, skew_M_pose_Rysync, kurtosis_M_pose_Rysync, min_M_pose_Rzsync, max_M_pose_Rzsync, sd_M_pose_Rzsync, mean_M_pose_Rzsync, md_M_pose_Rzsync, skew_M_pose_Rzsync, kurtosis_M_pose_Rzsync |
| **INTRAsync (14 features)** | min_M_intra, max_M_intra, sd_M_intra, mean_M_intra, md_M_intra, skew_M_intra, kurtosis_M_intra |
| **MovEx (6 features)** | M_body_total_movement, M_head_total_movement, mean_intensity_M |
| **Speech (30 features)** | dyad_pit_sync_MEA_M_speech, dyad_int_sync_MEA_M_speech, dyad_spr_M_speech, dyad_str_M_speech, dyad_ttg_M_speech, dyad_no_turns_M_speech, nsyll_M_speech, npause_M_speech, pho_M_speech, art_M_speech, pit_sync_M_speech, int_sync_M_speech, art_sync_M_speech, pit_var_M_speech, int_var_M_speech |

### S5 Model performance

#### S5.1 Distinguishing BPD-involved from COMP interactions

While developing an algorithm for technology-assisted diagnostics of BPD was not the explicit goal of this research project, we explored the application of our features to the classification between BPD-involved and COMP interactions. Despite the features being chosen with symptoms and characteristics of ASD in mind, the *CROSSturn*, *FACEsync*, *HEADsync* and *Speech* models performed above chance in this comparison (*BODYsync*: $p_{FDR} = 1$; *CROSSsync*: $p_{FDR} = 0.282$; *INTRAsync*: $p_{FDR} = 1$; *MovEx*: $p_{FDR} = 0.242$). Specifically, the *HEADsync* model achieved 68.7% balanced accuracy (71.4; 65.9% specificity), the *FACEsync* 64% (64.3; 65.9% specificity), the *Speech* 61.6% (59.5; 63.6% specificity) and the *CROSSturn* 55.7% (52.4; 59.1% specificity). The stacking model performed comparable to the *HEADsync* and the *MovEx* model but outperformed the other base models (see [!T]), reaching 65.3% balanced accuracy (73.8; 56.8% specificity). Thus, the stacking model only misclassified eleven BPD-involved interactions as COMP, but 19 COMP interactions were labelled as BPD-involved.

#### S5.2 One-verus-One comparisons

Table S4: Performance of all the models in One-versus-One comparisons

| comparison | model | sens | spec | BAC | AUC | p.fdr | sig |
|---|---|---|---|---|---|---|---|
| ASD-COMP vs BPD-COMP | BODYsync | 32.353 | 47.619 | 39.986 | 0.361 | 1.000 | |
| ASD-COMP vs BPD-COMP | CROSSsync | 52.941 | 69.048 | 60.994 | 0.678 | 0.136 | |
| ASD-COMP vs BPD-COMP | CROSSturn | 67.647 | 85.714 | 76.681 | 0.749 | 0.000 | * |
| ASD-COMP vs BPD-COMP | FACEsync | 76.471 | 59.524 | 67.997 | 0.719 | 0.000 | * |
| ASD-COMP vs BPD-COMP | HEADsync | 55.882 | 57.143 | 56.513 | 0.609 | 0.936 | |
| ASD-COMP vs BPD-COMP | INTRAsync | 38.235 | 54.762 | 46.499 | 0.455 | 1.000 | |
| ASD-COMP vs BPD-COMP | MovEx | 58.824 | 78.571 | 68.698 | 0.758 | 0.027 | * |
| ASD-COMP vs BPD-COMP | Speech | 64.706 | 76.190 | 70.448 | 0.771 | 0.000 | * |
| ASD-COMP vs BPD-COMP | STACK | 70.588 | 92.857 | 81.723 | 0.805 | NaN | NA |
| ASD-COMP vs COMP-COMP | BODYsync | 58.824 | 61.364 | 60.094 | 0.607 | 0.000 | * |
| ASD-COMP vs COMP-COMP | CROSSsync | 67.647 | 75.000 | 71.324 | 0.763 | 0.000 | * |
| ASD-COMP vs COMP-COMP | CROSSturn | 44.118 | 72.727 | 58.422 | 0.627 | 0.000 | * |

| | | | | | | | |
|---|---|---|---|---|---|---|---|
| ASD-COMP vs COMP-COMP | FACEsync | 79.412 | 70.454 | 74.933 | 0.780 | 0.000 | * |
| ASD-COMP vs COMP-COMP | HEADsync | 58.824 | 68.182 | 63.503 | 0.608 | 1.000 | |
| ASD-COMP vs COMP-COMP | INTRAsync | 26.471 | 40.909 | 33.690 | 0.297 | 1.000 | |
| ASD-COMP vs COMP-COMP | MovEx | 64.706 | 72.727 | 68.717 | 0.777 | 0.000 | * |
| ASD-COMP vs COMP-COMP | Speech | 64.706 | 72.727 | 68.717 | 0.673 | 0.000 | * |
| ASD-COMP vs COMP-COMP | STACK | 79.412 | 81.818 | 80.615 | 0.845 | NaN | NA |
| BPD-COMP vs COMP-COMP | BODYsync | 28.571 | 36.364 | 32.468 | 0.308 | 1.000 | |
| BPD-COMP vs COMP-COMP | CROSSsync | 50.000 | 61.364 | 55.682 | 0.594 | 0.282 | |
| BPD-COMP vs COMP-COMP | CROSSturn | 52.381 | 59.091 | 55.736 | 0.569 | 0.045 | * |
| BPD-COMP vs COMP-COMP | FACEsync | 64.286 | 63.636 | 63.961 | 0.670 | 0.007 | * |
| BPD-COMP vs COMP-COMP | HEADsync | 71.429 | 65.909 | 68.669 | 0.736 | 0.000 | * |
| BPD-COMP vs COMP-COMP | INTRAsync | 57.143 | 54.545 | 55.844 | 0.516 | 1.000 | |
| BPD-COMP vs COMP-COMP | MovEx | 69.048 | 70.454 | 69.751 | 0.686 | 0.242 | |
| BPD-COMP vs COMP-COMP | Speech | 59.524 | 63.636 | 61.580 | 0.693 | 0.007 | * |
| BPD-COMP vs COMP-COMP | STACK | 73.810 | 56.818 | 65.314 | 0.760 | NaN | NA |

#### S5.3 Multi-group comparisons

Table S5: Performance of all the models in Multi-group comparisons

| comparison | model | BAC | p.fdr | sig |
|---|---|---|---|---|
| MultiGroup | BODYsync | 48.6 | 1.000 | |
| MultiGroup | CROSSsync | 58.1 | 0.000 | * |
| MultiGroup | CROSSturn | 57.3 | 0.000 | * |
| MultiGroup | FACEsync | 57.6 | 0.007 | * |
| MultiGroup | HEADsync | 54.6 | 0.000 | * |
| MultiGroup | INTRAsync | 49.5 | 1.000 | |
| MultiGroup | MovEx | 60.8 | 0.000 | * |
| MultiGroup | Speech | 63.9 | 0.000 | * |
| MultiGroup | STACK | 63.2 | NaN | NA |

#### S5.4 Gender comparisons

Did the models perform better for one gender than the other? We perform unpaired Wilcoxon tests for the labels and models separately.

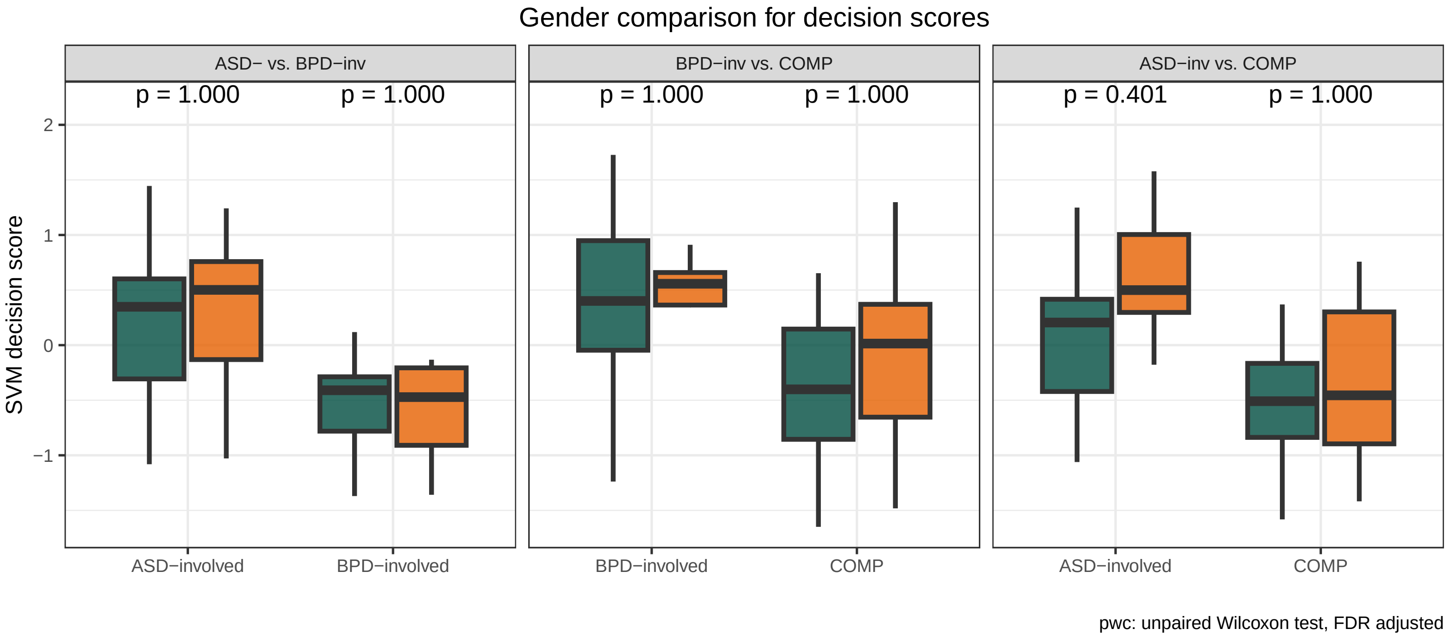

### S6 Model visualisations

The following figures were created inspired by NeuroMiner[15] visualisations and show the sign-based consistency[16] as well as the feature weights of the models distinguishing between interaction partners from ASD- and BPD-involved interactions. For models with a large number of features, the top 16 features are plotted.

##### S6.1 ASD-involved versus BPD-involved classifiers

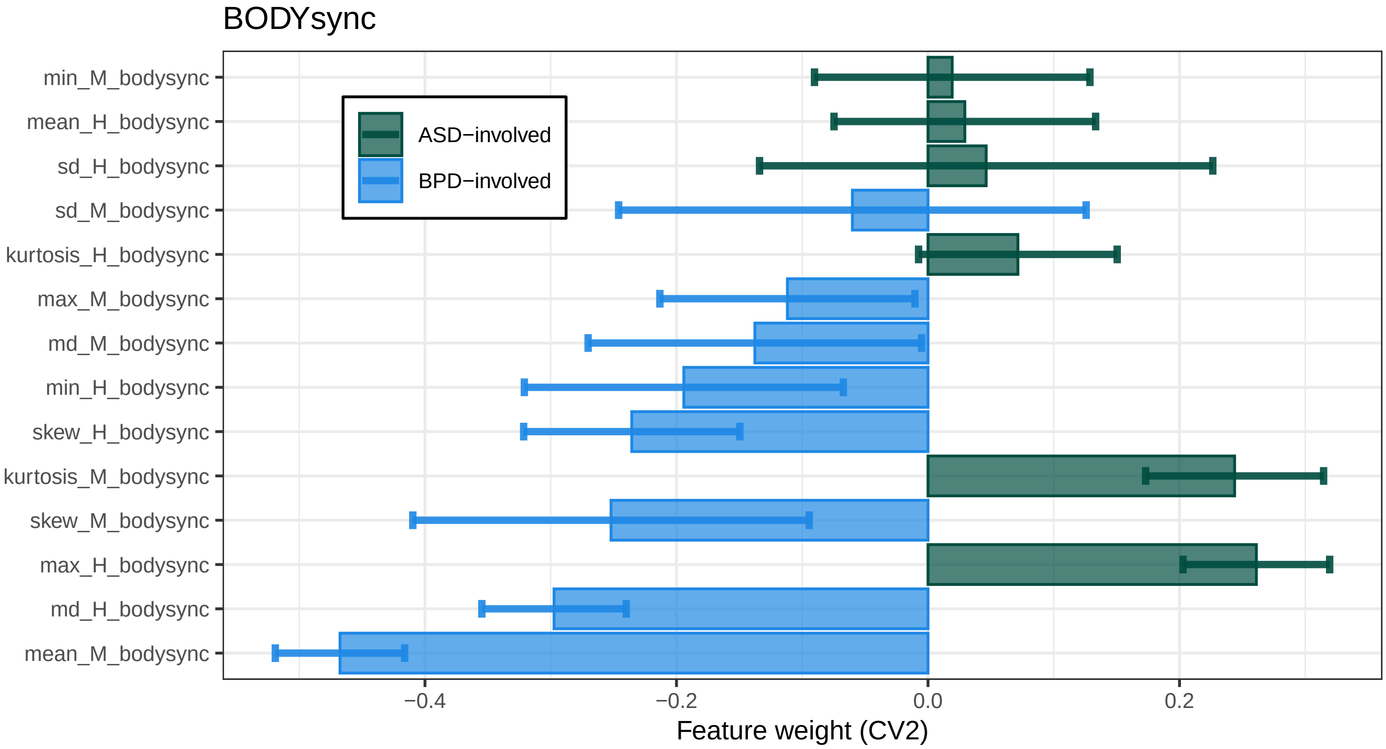

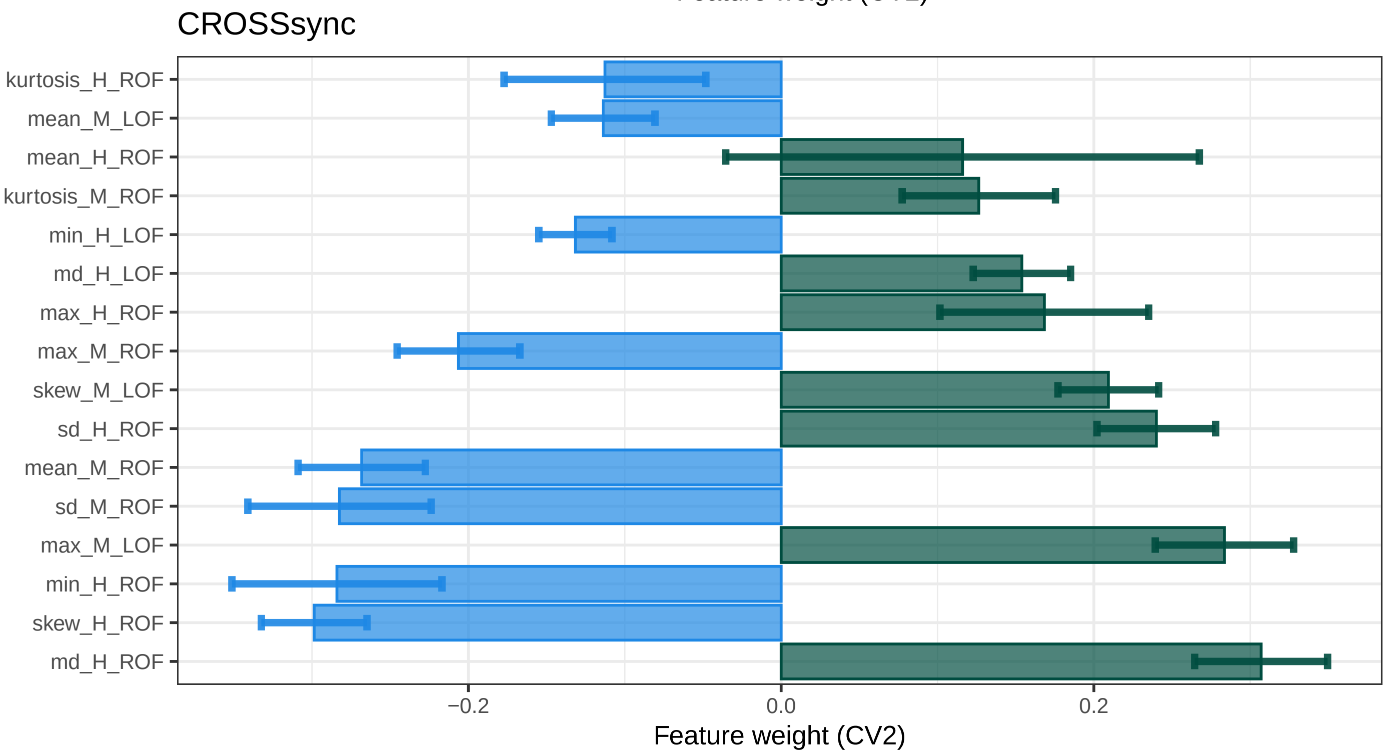

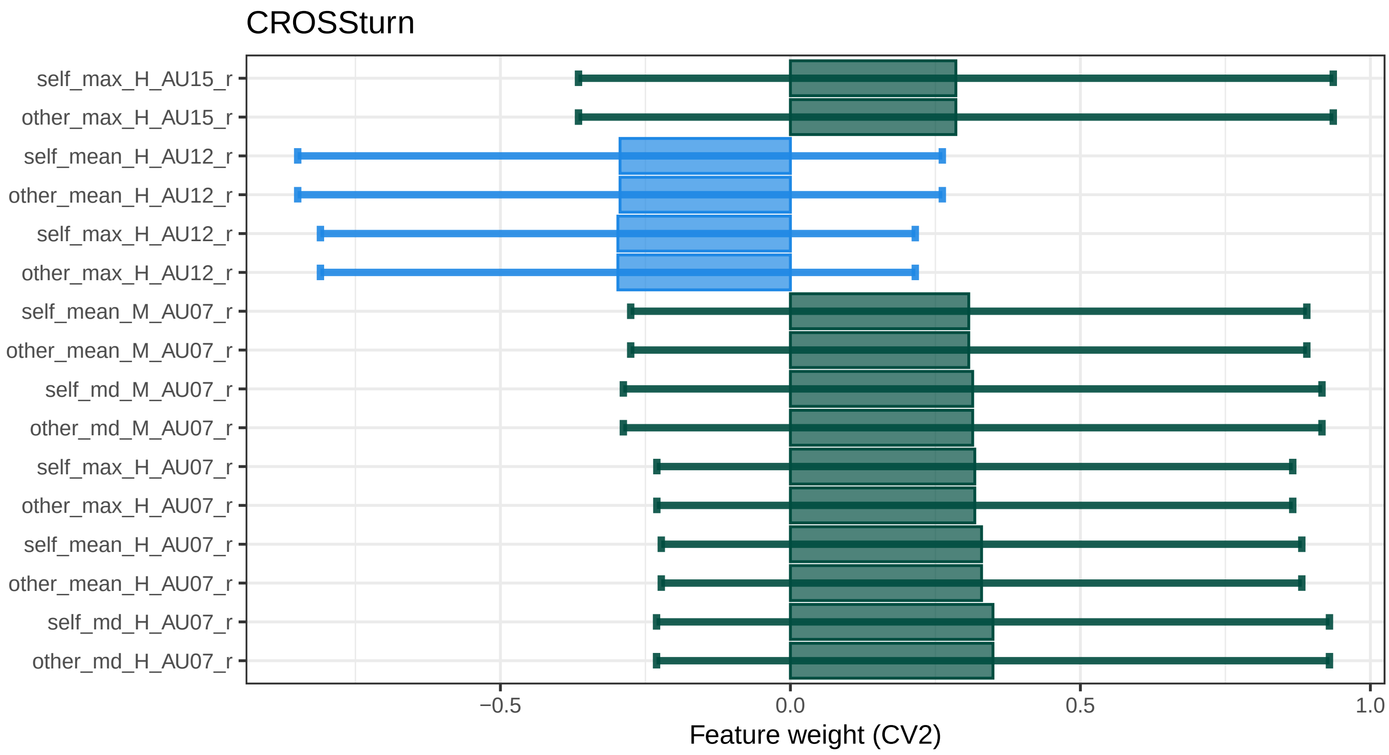
CROSSturn
self_max_H_AU15_r
other_max_H_AU15_r
self_mean_H_AU12_r
other_mean_H_AU12_r
self_max_H_AU12_r
other_max_H_AU12_r
self_mean_M_AU07_r
other_mean_M_AU07_r
self_md_M_AU07_r
other_md_M_AU07_r
self_max_H_AU07_r
other_max_H_AU07_r
self_mean_H_AU07_r
other_mean_H_AU07_r
self_md_H_AU07_r
other_md_H_AU07_r
−0.5
0.0
0.5
1.0
Feature weight (CV2)

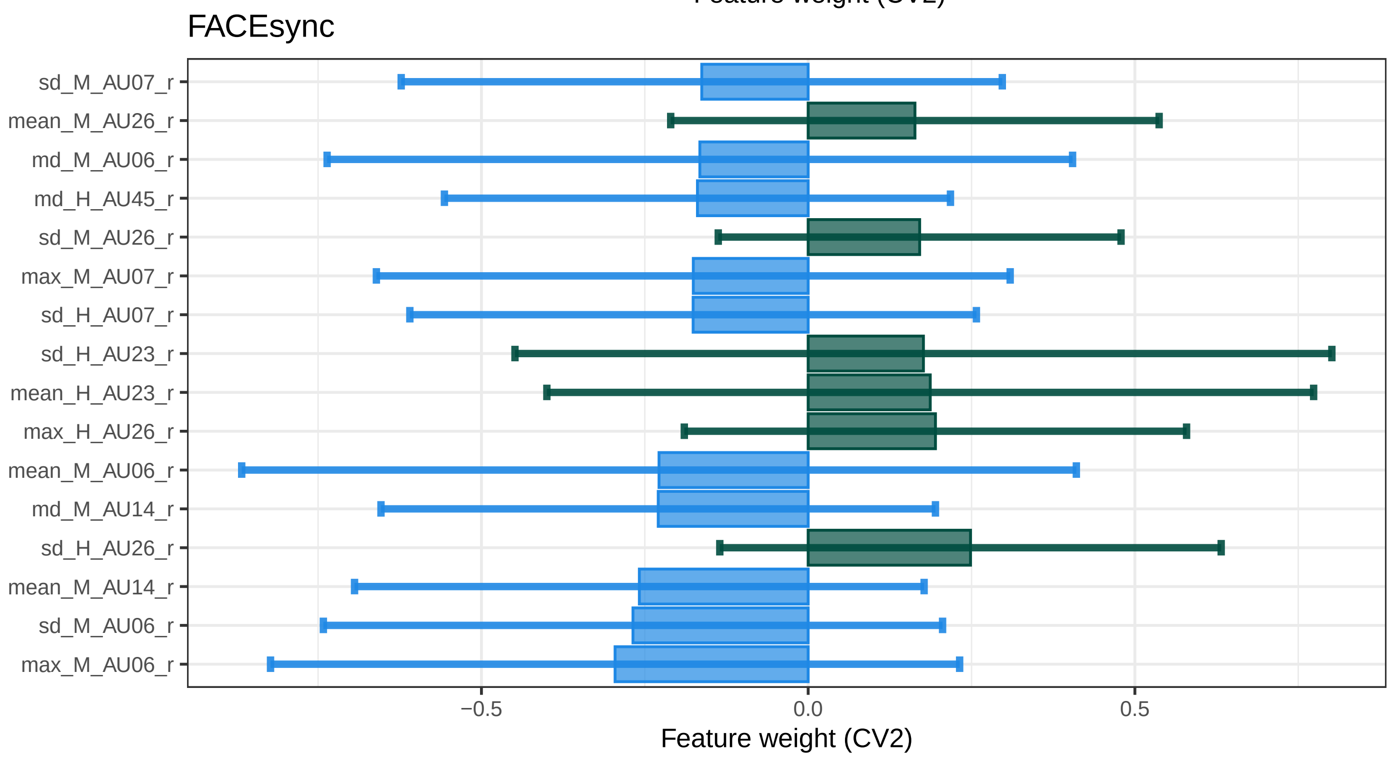
FACEsync
sd_M_AU07_r
mean_M_AU26_r
md_M_AU06_r
md_H_AU45_r
sd_M_AU26_r
max_M_AU07_r
sd_H_AU07_r
sd_H_AU23_r
mean_H_AU23_r
max_H_AU26_r
mean_M_AU06_r
md_M_AU14_r
sd_H_AU26_r
mean_M_AU14_r
sd_M_AU06_r
max_M_AU06_r
−0.5
0.0
0.5
Feature weight (CV2)

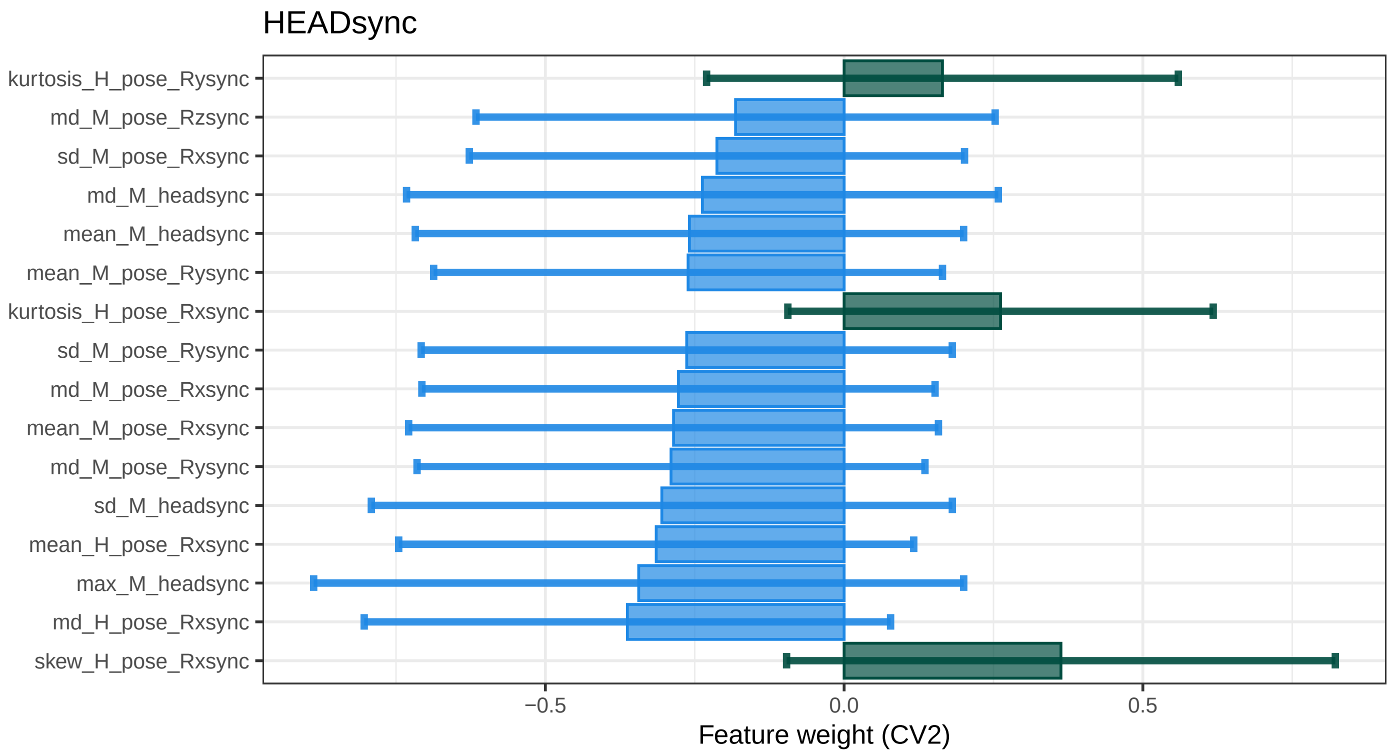
HEADsync
kurtosis_H_pose_Rysync
md_M_pose_Rzsync
sd_M_pose_Rxsync
md_M_headsync
mean_M_headsync
mean_M_pose_Rysync
kurtosis_H_pose_Rxsync
sd_M_pose_Rysync
md_M_pose_Rxsync
mean_M_pose_Rxsync
md_M_pose_Rysync
sd_M_headsync
mean_H_pose_Rxsync
max_M_headsync
md_H_pose_Rxsync
skew_H_pose_Rxsync
−0.5
0.0
0.5
Feature weight (CV2)

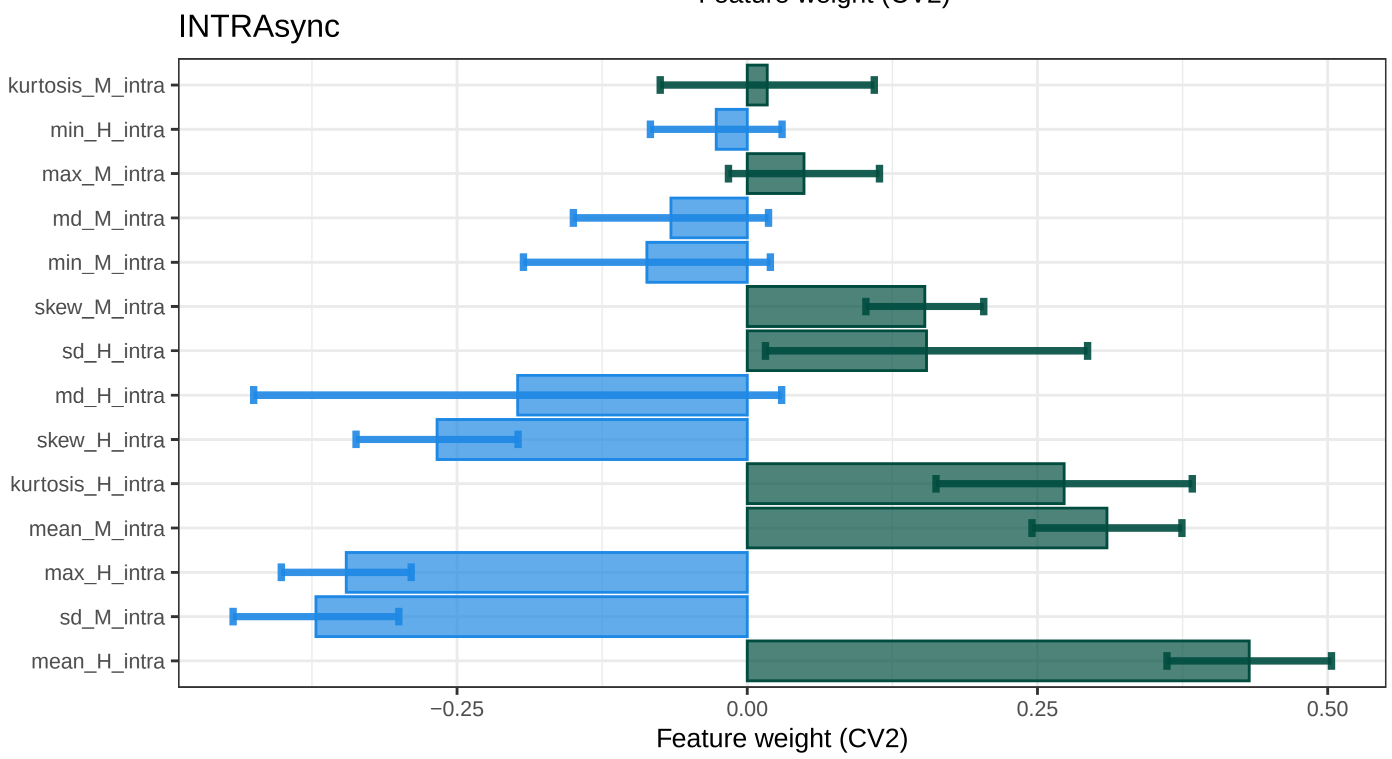
INTRAsync
kurtosis_M_intra
min_H_intra
max_M_intra
md_M_intra
min_M_intra
skew_M_intra
sd_H_intra
md_H_intra
skew_H_intra
kurtosis_H_intra
mean_M_intra
max_H_intra
sd_M_intra
mean_H_intra
−0.25
0.00
0.25
0.50
Feature weight (CV2)

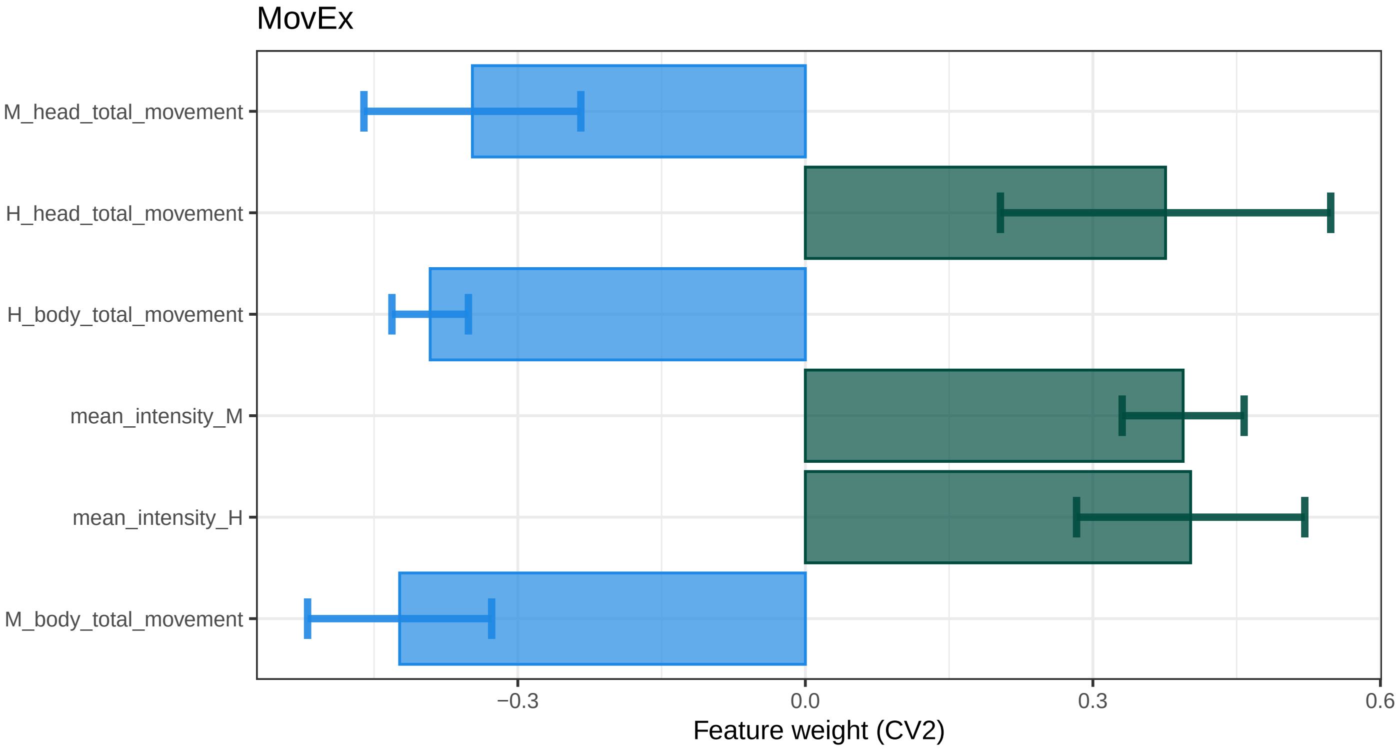
MovEx
M_head_total_movement
H_head_total_movement
H_body_total_movement
mean_intensity_M
mean_intensity_H
M_body_total_movement
−0.3
0.0
0.3
0.6
Feature weight (CV2)

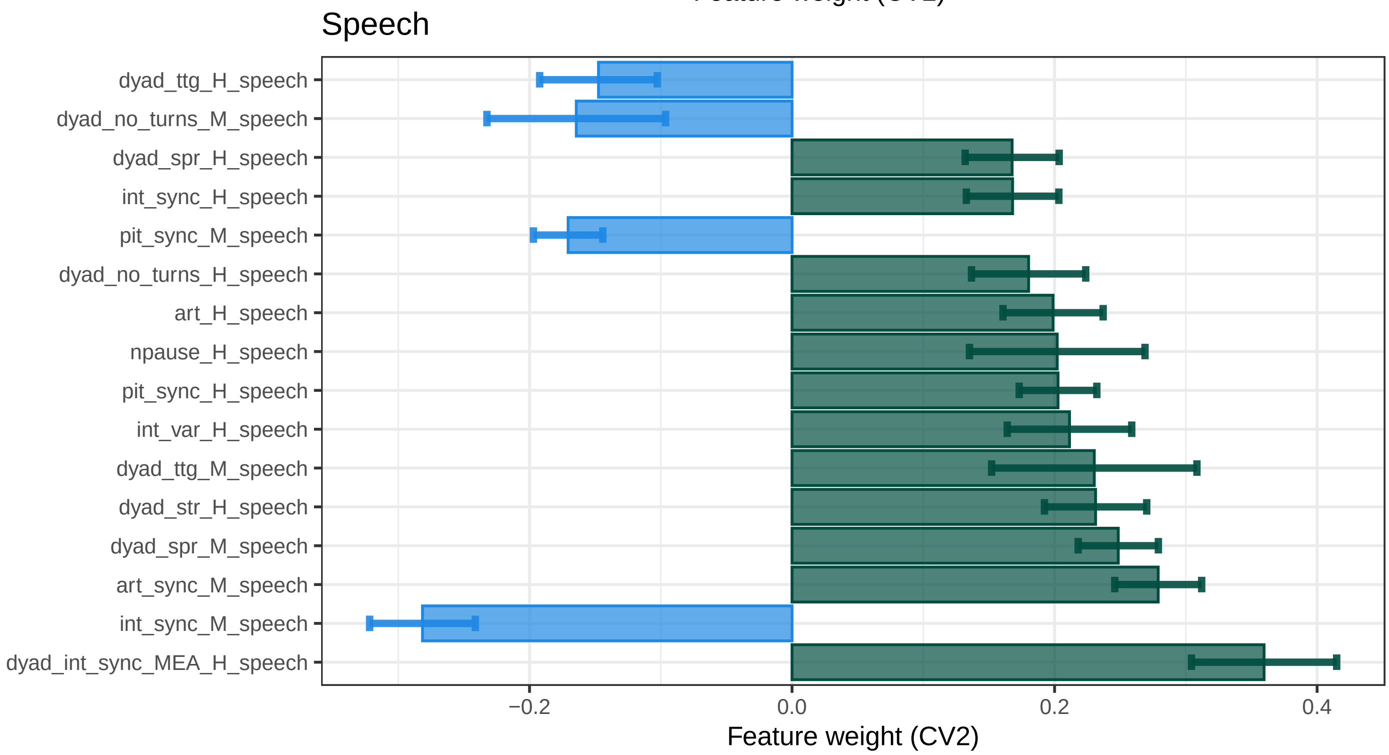
Speech
dyad_ttg_H_speech
dyad_no_turns_M_speech
dyad_spr_H_speech
int_sync_H_speech
pit_sync_M_speech
dyad_no_turns_H_speech
art_H_speech
npause_H_speech
pit_sync_H_speech
int_var_H_speech
dyad_ttg_M_speech
dyad_str_H_speech
dyad_spr_M_speech
art_sync_M_speech
int_sync_M_speech
dyad_int_sync_MEA_H_speech
−0.2
0.0
0.2
0.4
Feature weight (CV2)

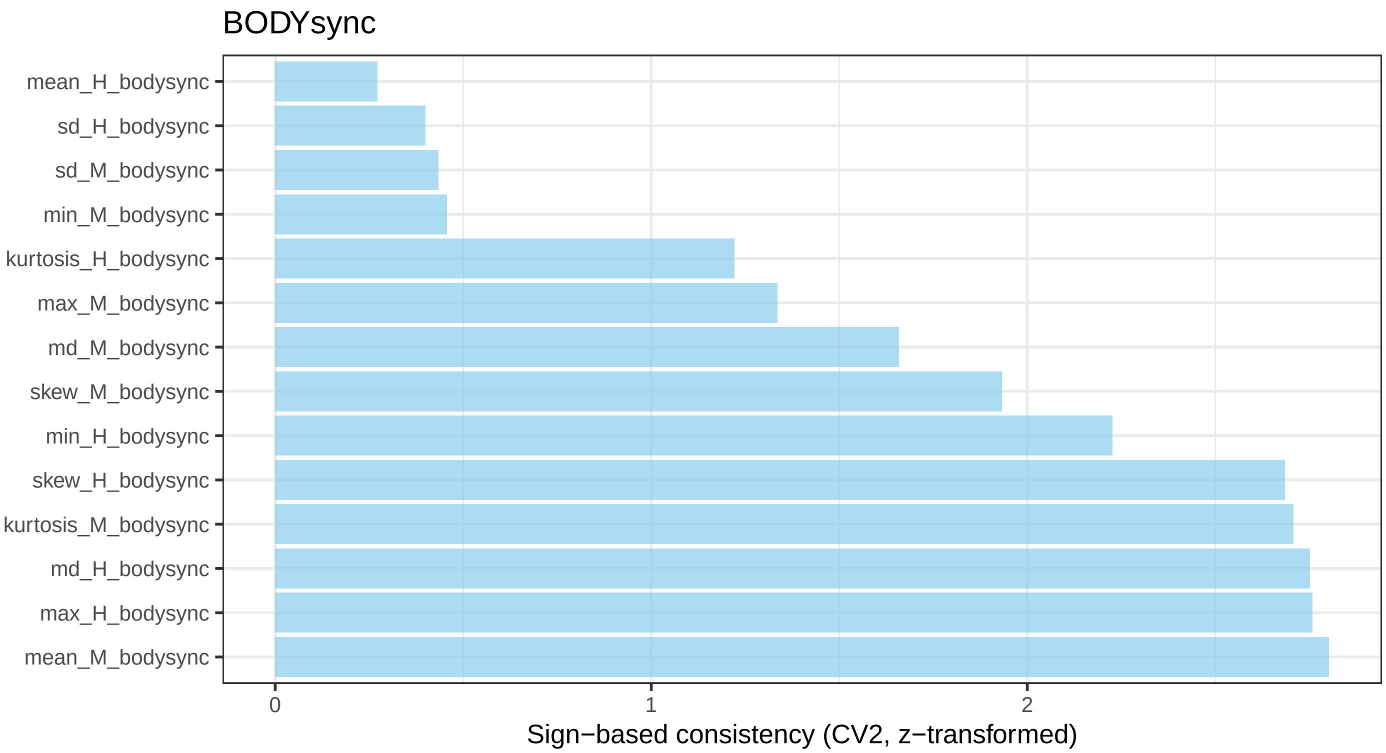
BODYsync
mean_H_bodysync
sd_H_bodysync
sd_M_bodysync
min_M_bodysync
kurtosis_H_bodysync
max_M_bodysync
md_M_bodysync
skew_M_bodysync
min_H_bodysync
skew_H_bodysync
kurtosis_M_bodysync
md_H_bodysync
max_H_bodysync
mean_M_bodysync
0
1
2
Sign−based consistency (CV2, z−transformed)

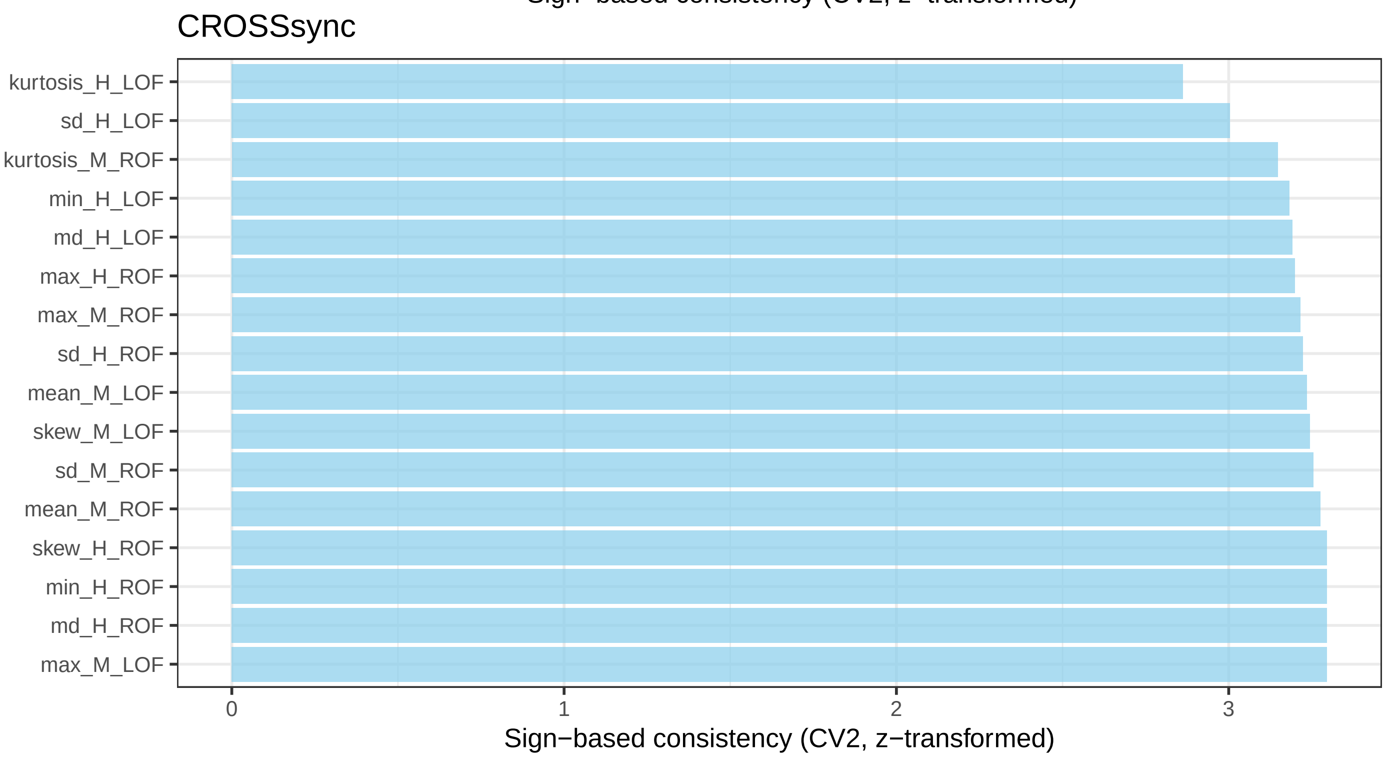
CROSSsync
kurtosis_H_LOF
sd_H_LOF
kurtosis_M_ROF
min_H_LOF
md_H_LOF
max_H_ROF
max_M_ROF
sd_H_ROF
mean_M_LOF
skew_M_LOF
sd_M_ROF
mean_M_ROF
skew_H_ROF
min_H_ROF
md_H_ROF
max_M_LOF
0
1
2
3
Sign−based consistency (CV2, z−transformed)

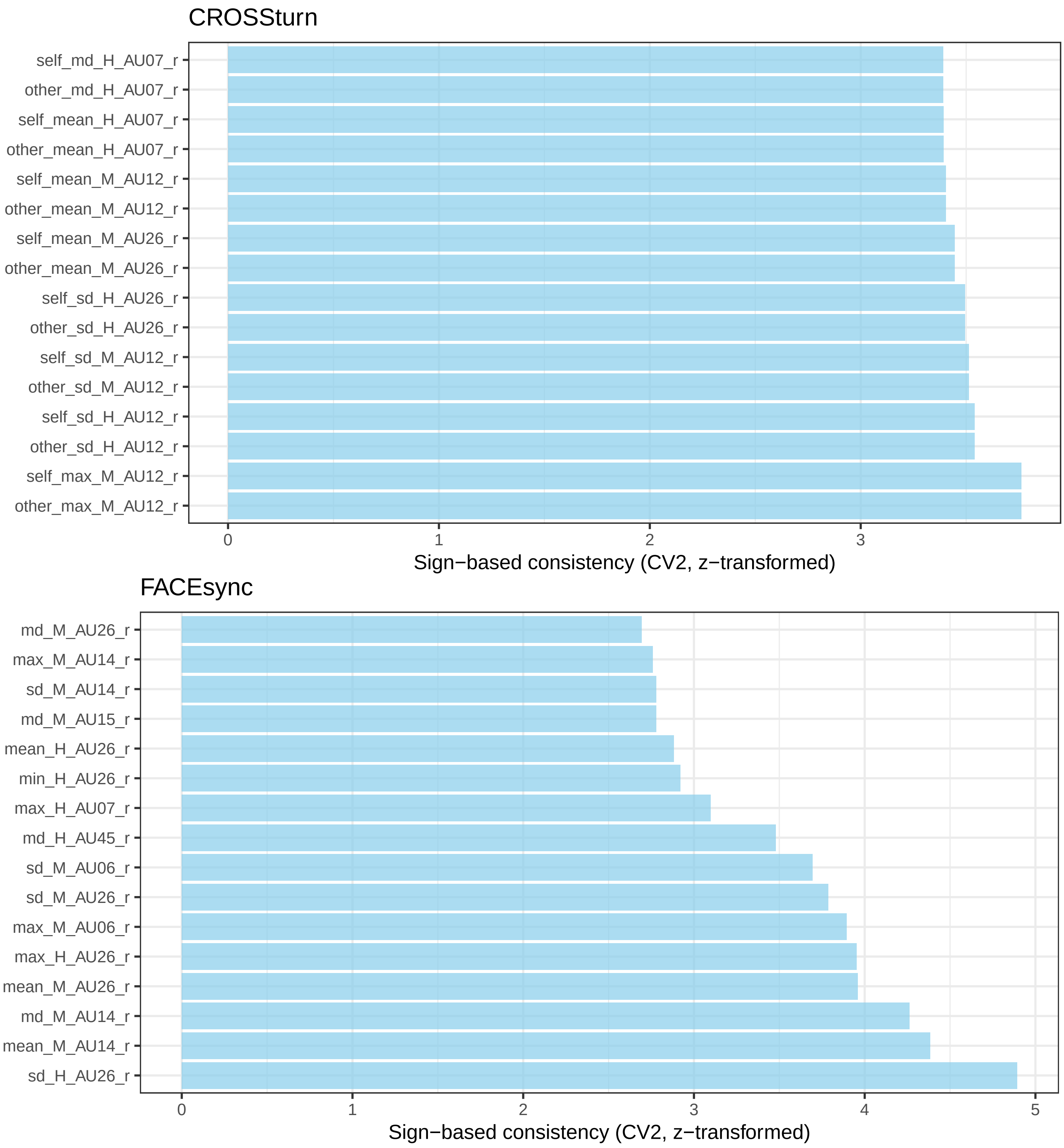
CROSSturn
self_md_H_AU07_r
other_md_H_AU07_r
self_mean_H_AU07_r
other_mean_H_AU07_r
self_mean_M_AU12_r
other_mean_M_AU12_r
self_mean_M_AU26_r
other_mean_M_AU26_r
self_sd_H_AU26_r
other_sd_H_AU26_r
self_sd_M_AU12_r
other_sd_M_AU12_r
self_sd_H_AU12_r
other_sd_H_AU12_r
self_max_M_AU12_r
other_max_M_AU12_r
0
1
2
3
Sign−based consistency (CV2, z−transformed)
FACEsync
md_M_AU26_r
max_M_AU14_r
sd_M_AU14_r
md_M_AU15_r
mean_H_AU26_r
min_H_AU26_r
max_H_AU07_r
md_H_AU45_r
sd_M_AU06_r
sd_M_AU26_r
max_M_AU06_r
max_H_AU26_r
mean_M_AU26_r
md_M_AU14_r
mean_M_AU14_r
sd_H_AU26_r
0
1
2
3
4
5
Sign−based consistency (CV2, z−transformed)

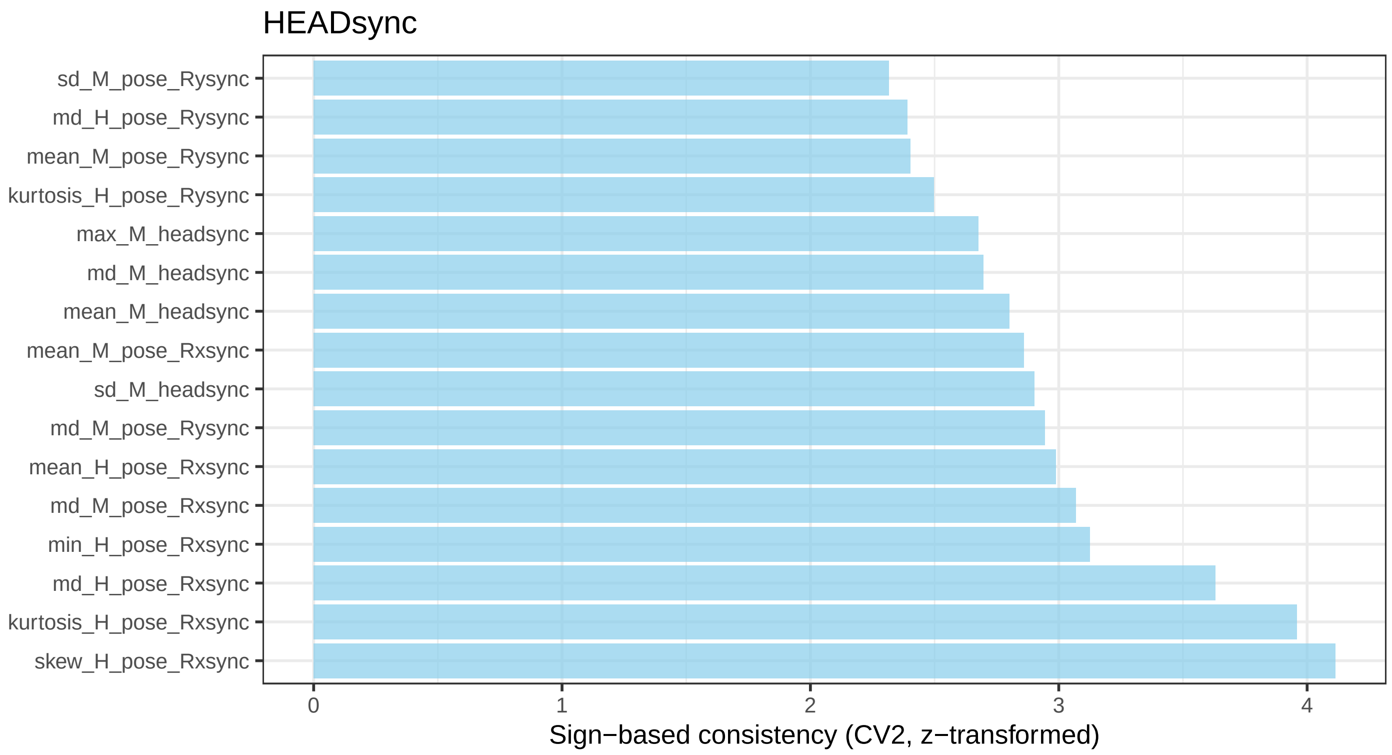
HEADsync
sd_M_pose_Rysync
md_H_pose_Rysync
mean_M_pose_Rysync
kurtosis_H_pose_Rysync
max_M_headsync
md_M_headsync
mean_M_headsync
mean_M_pose_Rxsync
sd_M_headsync
md_M_pose_Rysync
mean_H_pose_Rxsync
md_M_pose_Rxsync
min_H_pose_Rxsync
md_H_pose_Rxsync
kurtosis_H_pose_Rxsync
skew_H_pose_Rxsync
0
1
2
3
4
Sign−based consistency (CV2, z−transformed)

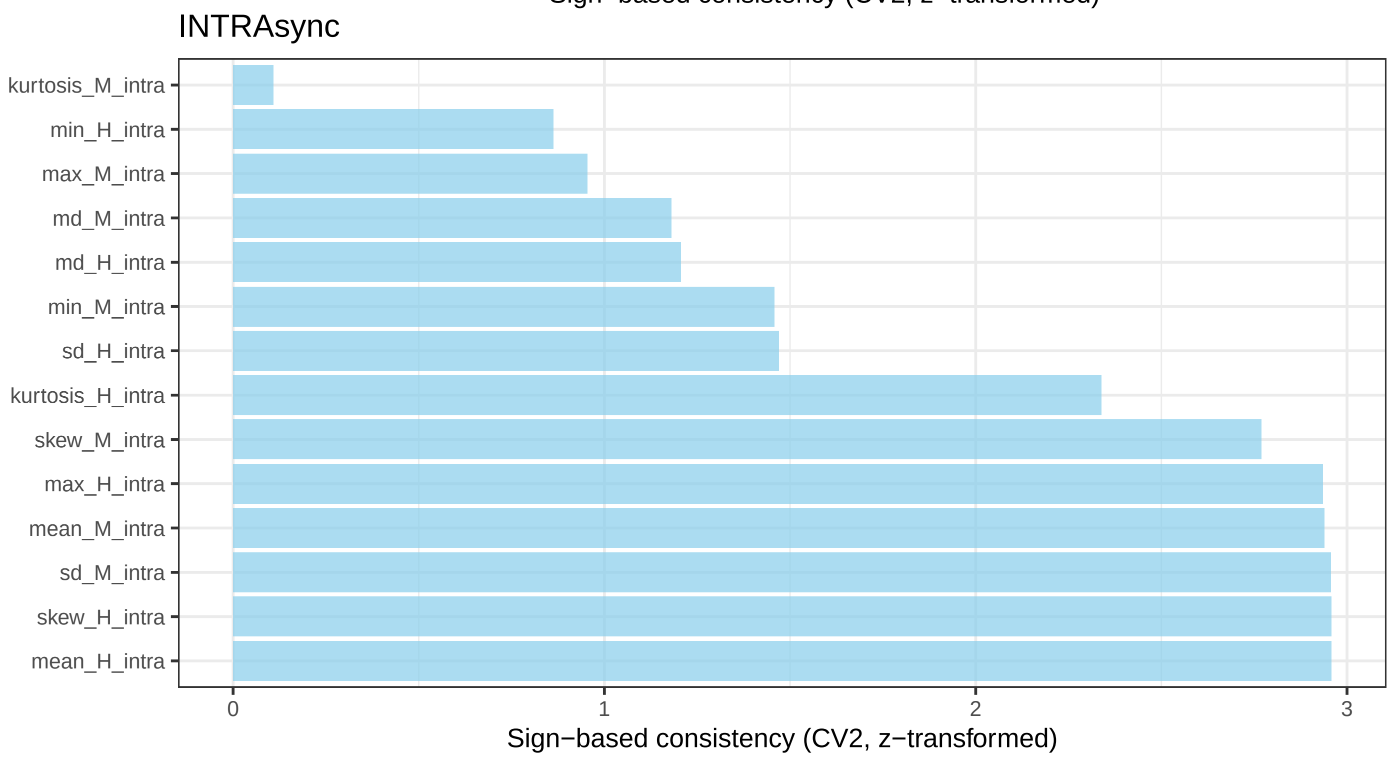
INTRAsync
kurtosis_M_intra
min_H_intra
max_M_intra
md_M_intra
md_H_intra
min_M_intra
sd_H_intra
kurtosis_H_intra
skew_M_intra
max_H_intra
mean_M_intra
sd_M_intra
skew_H_intra
mean_H_intra
0
1
2
3
Sign−based consistency (CV2, z−transformed)

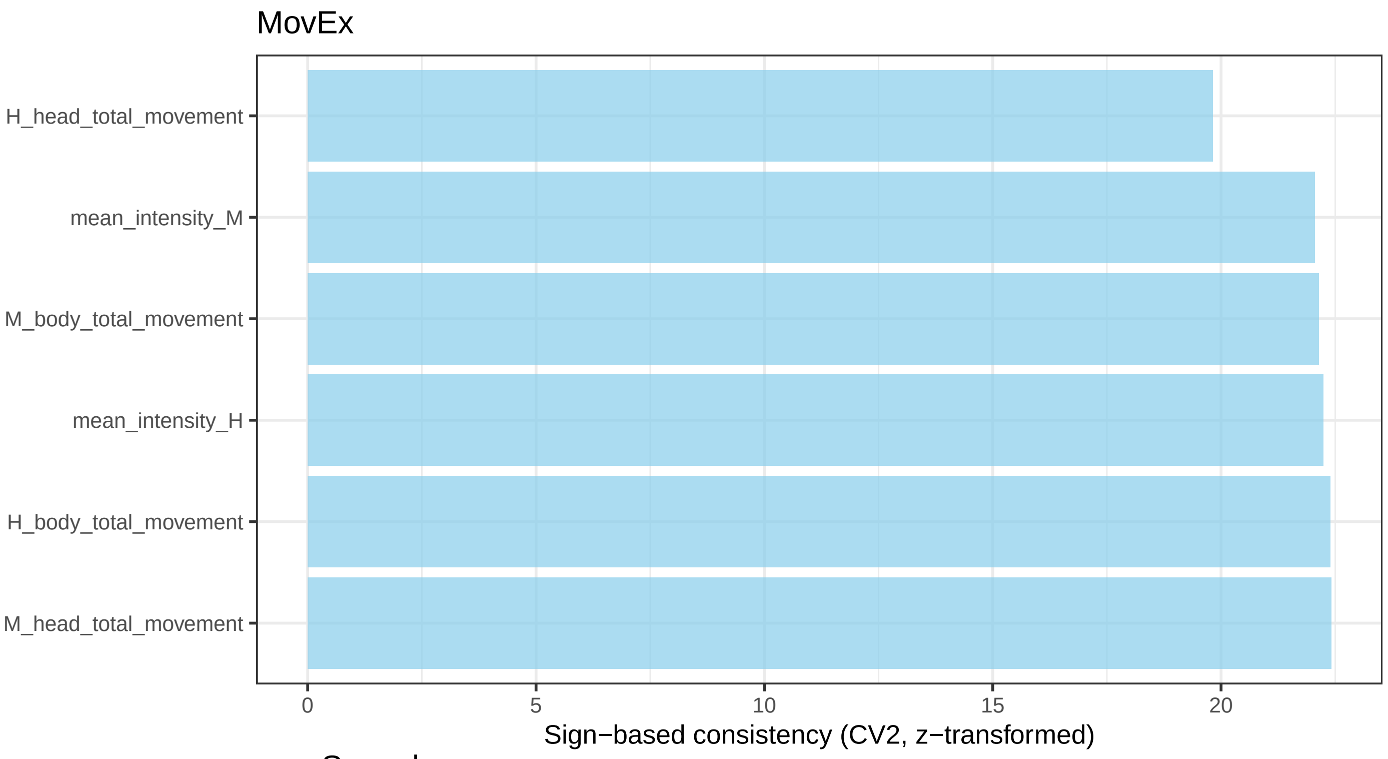
MovEx
H_head_total_movement
mean_intensity_M
M_body_total_movement
mean_intensity_H
H_body_total_movement
M_head_total_movement
0
5
10
15
20
Sign−based consistency (CV2, z−transformed)

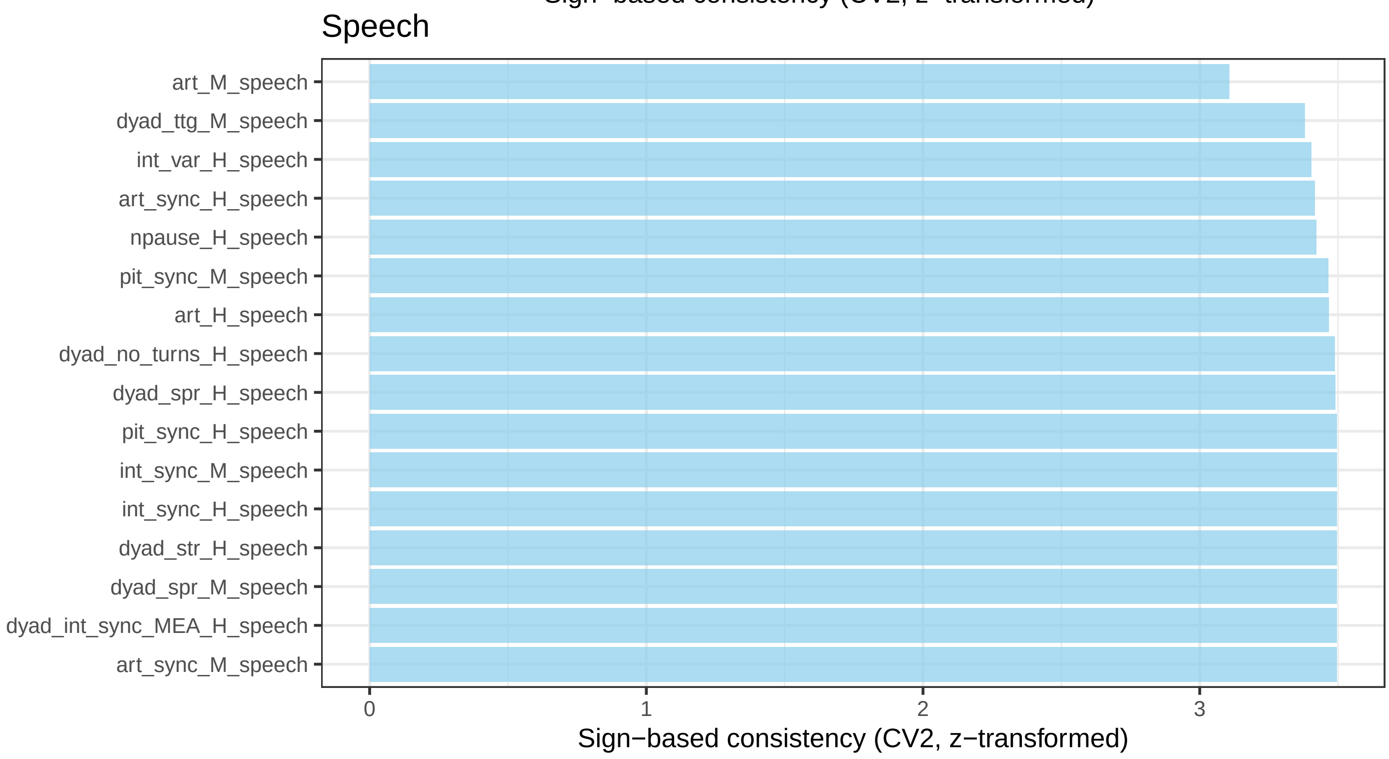
Speech
art_M_speech
dyad_ttg_M_speech
int_var_H_speech
art_sync_H_speech
npause_H_speech
pit_sync_M_speech
art_H_speech
dyad_no_turns_H_speech
dyad_spr_H_speech
pit_sync_H_speech
int_sync_M_speech
int_sync_H_speech
dyad_str_H_speech
dyad_spr_M_speech
dyad_int_sync_MEA_H_speech
art_sync_M_speech
0
1
2
3
Sign−based consistency (CV2, z−transformed)

##### S6.2 ASD-involved versus COMP classifiers

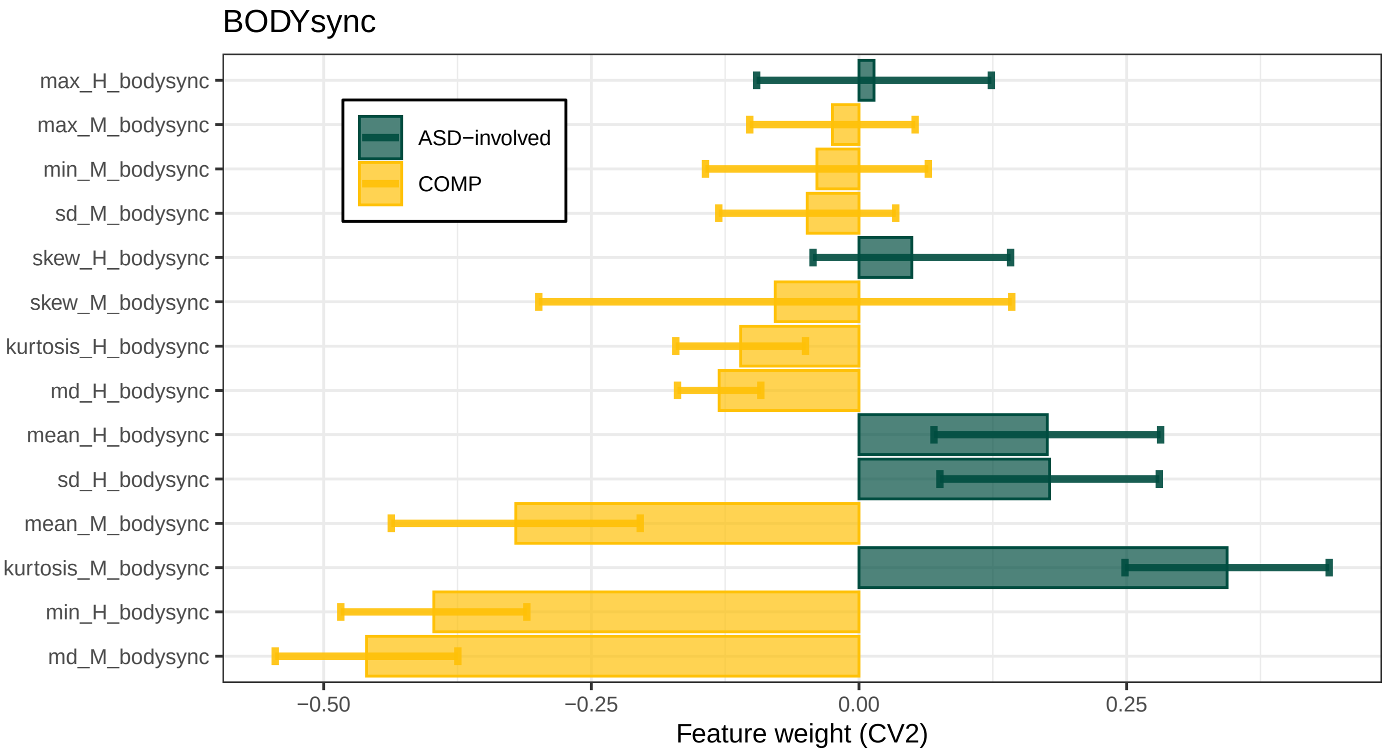

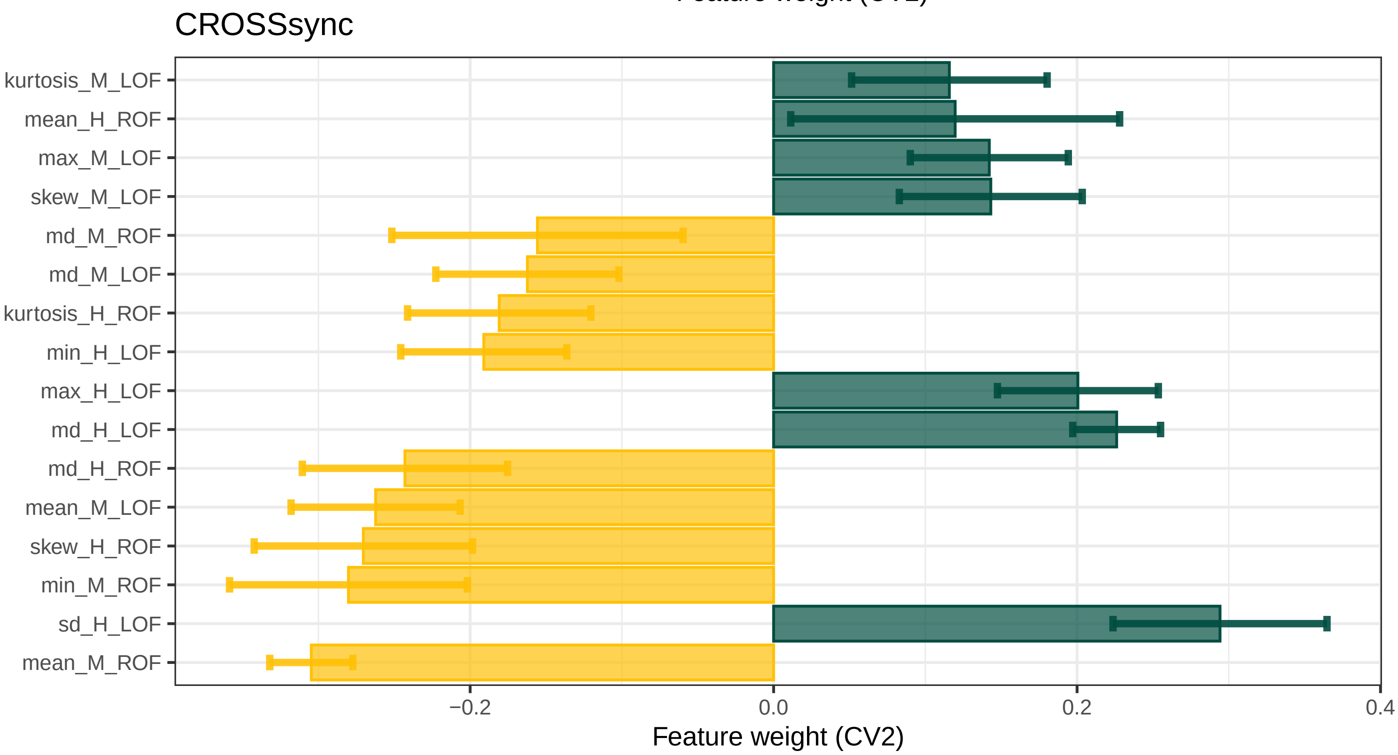

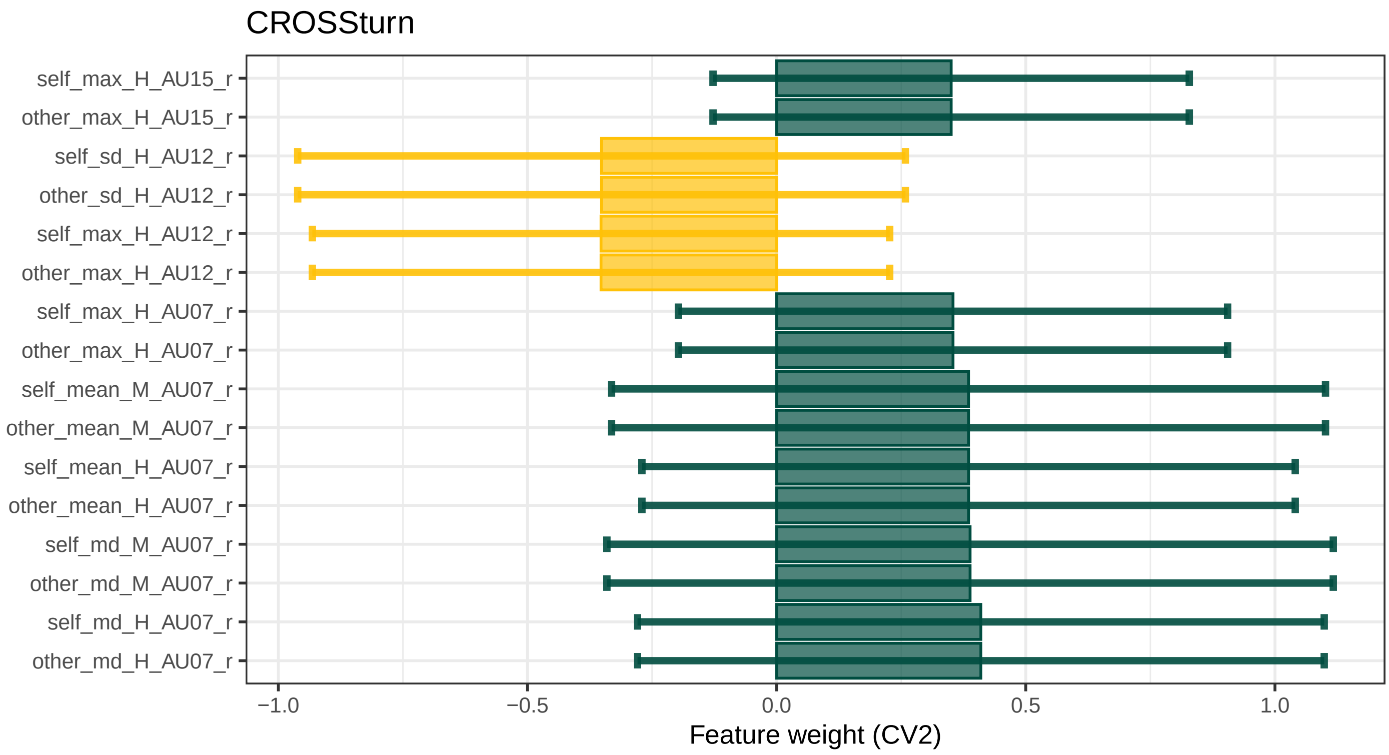
CROSSturn
self_max_H_AU15_r
other_max_H_AU15_r
self_sd_H_AU12_r
other_sd_H_AU12_r
self_max_H_AU12_r
other_max_H_AU12_r
self_max_H_AU07_r
other_max_H_AU07_r
self_mean_M_AU07_r
other_mean_M_AU07_r
self_mean_H_AU07_r
other_mean_H_AU07_r
self_md_M_AU07_r
other_md_M_AU07_r
self_md_H_AU07_r
other_md_H_AU07_r
−1.0
−0.5
0.0
0.5
1.0
Feature weight (CV2)

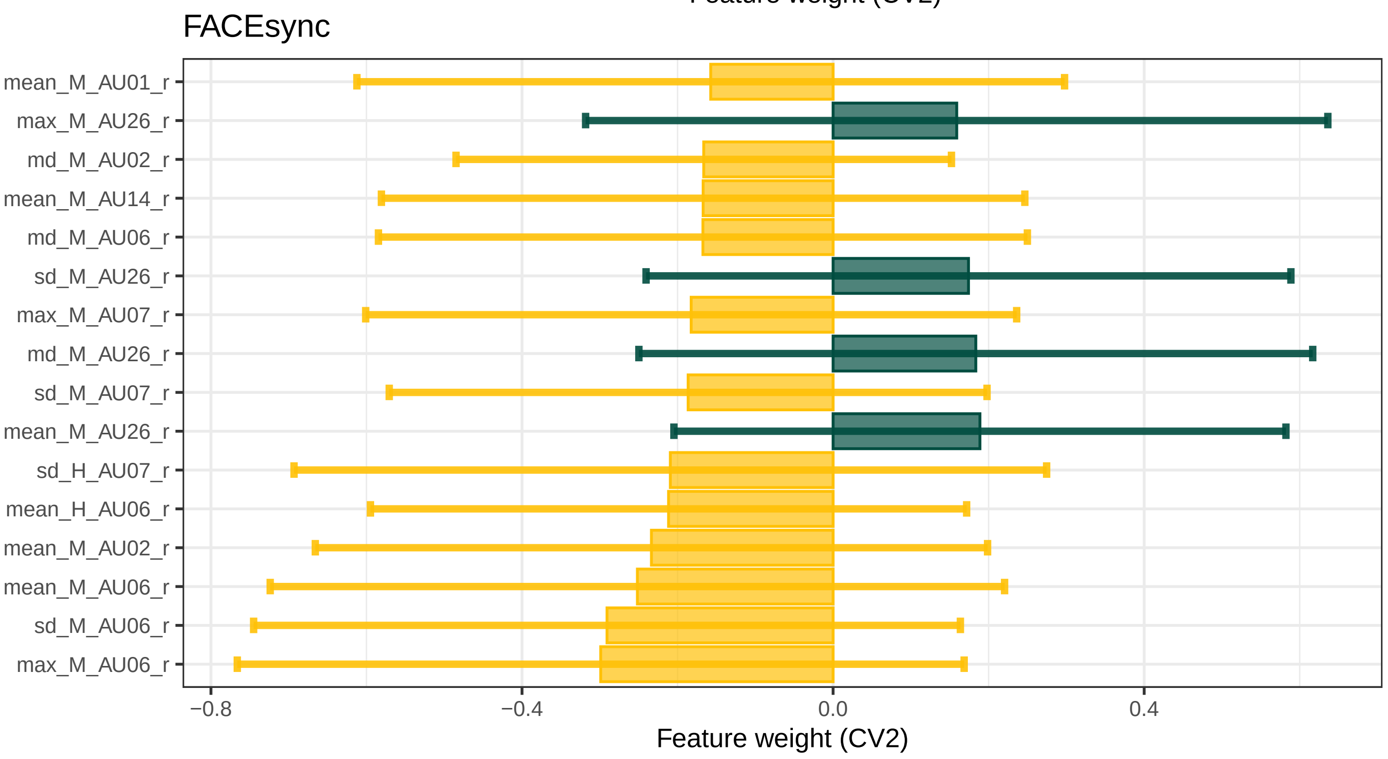
FACEsync
mean_M_AU01_r
max_M_AU26_r
md_M_AU02_r
mean_M_AU14_r
md_M_AU06_r
sd_M_AU26_r
max_M_AU07_r
md_M_AU26_r
sd_M_AU07_r
mean_M_AU26_r
sd_H_AU07_r
mean_H_AU06_r
mean_M_AU02_r
mean_M_AU06_r
sd_M_AU06_r
max_M_AU06_r
−0.8
−0.4
0.0
0.4
Feature weight (CV2)

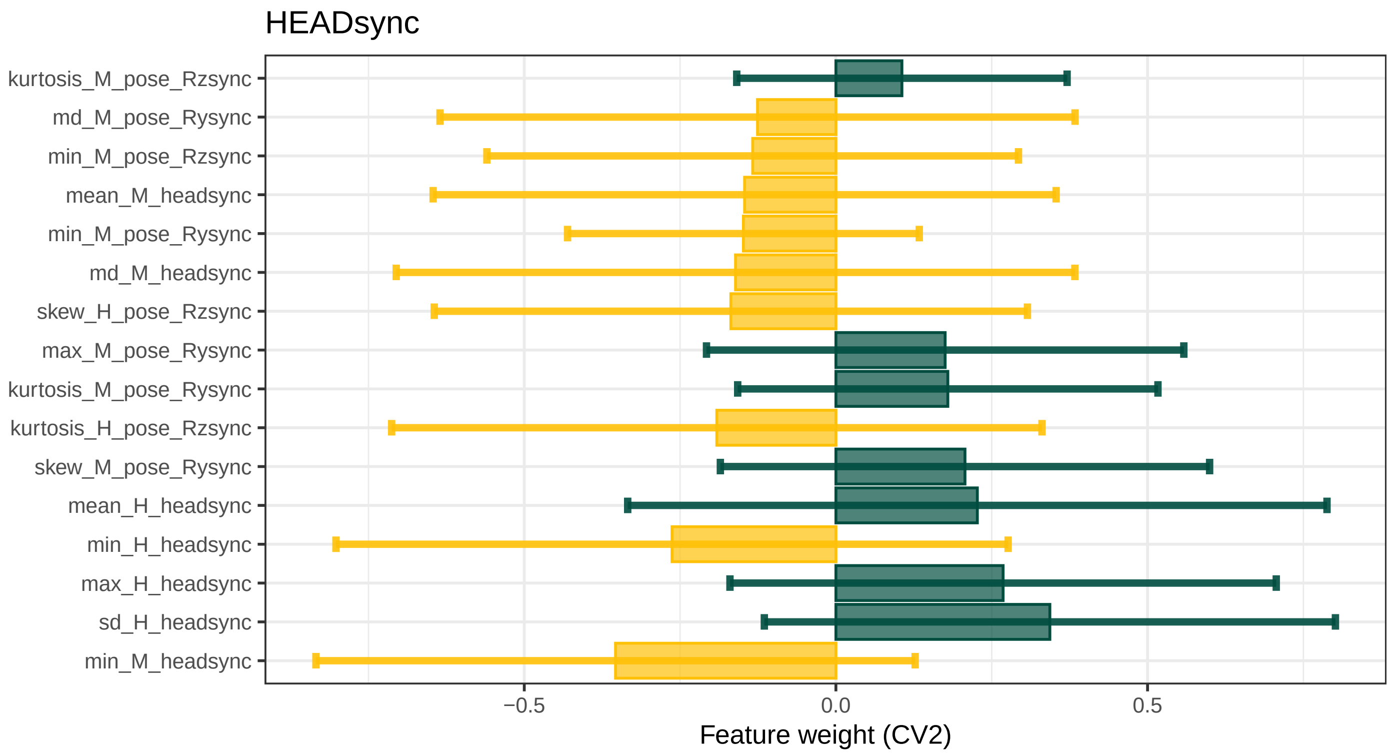
HEADsync
kurtosis_M_pose_Rzsync
md_M_pose_Rysync
min_M_pose_Rzsync
mean_M_headsync
min_M_pose_Rysync
md_M_headsync
skew_H_pose_Rzsync
max_M_pose_Rysync
kurtosis_M_pose_Rysync
kurtosis_H_pose_Rzsync
skew_M_pose_Rysync
mean_H_headsync
min_H_headsync
max_H_headsync
sd_H_headsync
min_M_headsync
−0.5
0.0
0.5
Feature weight (CV2)

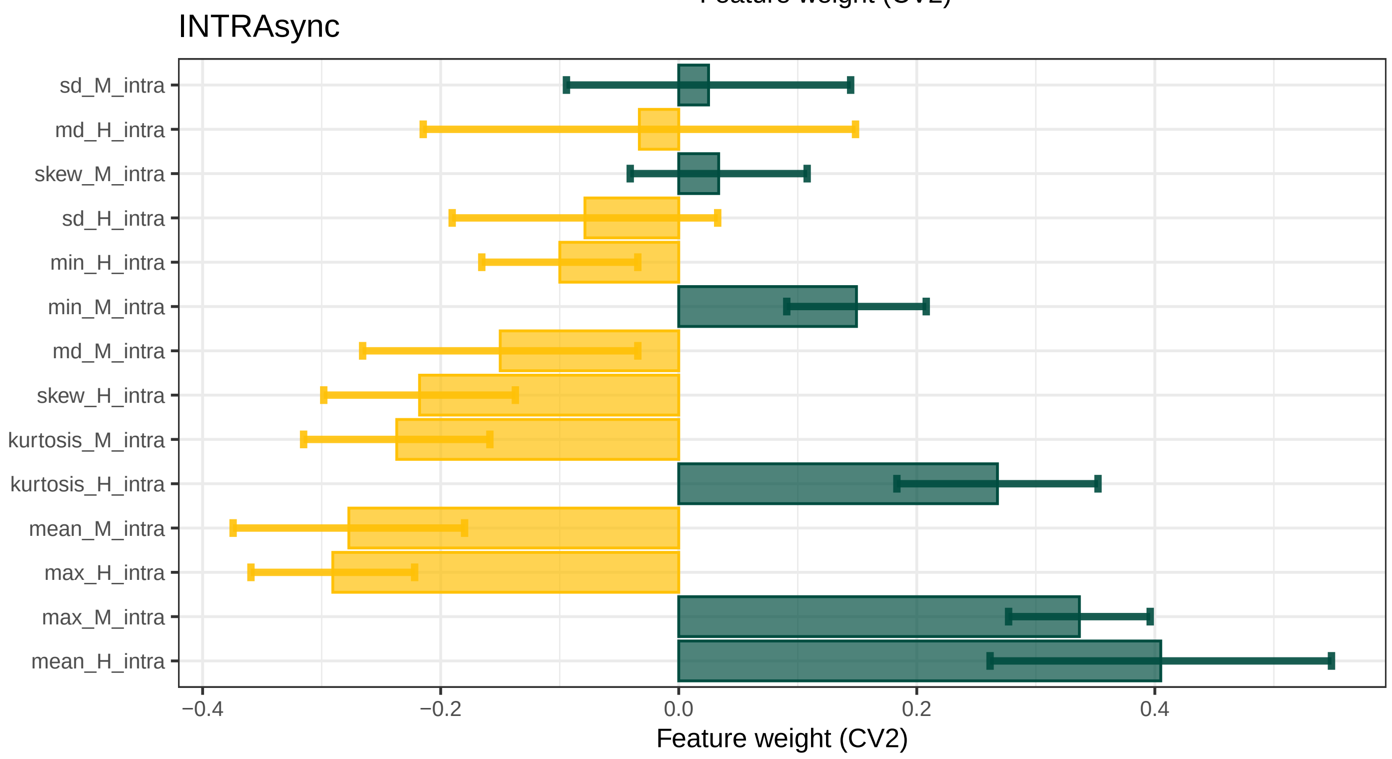
INTRAsync
sd_M_intra
md_H_intra
skew_M_intra
sd_H_intra
min_H_intra
min_M_intra
md_M_intra
skew_H_intra
kurtosis_M_intra
kurtosis_H_intra
mean_M_intra
max_H_intra
max_M_intra
mean_H_intra
−0.4
−0.2
0.0
0.2
0.4
Feature weight (CV2)

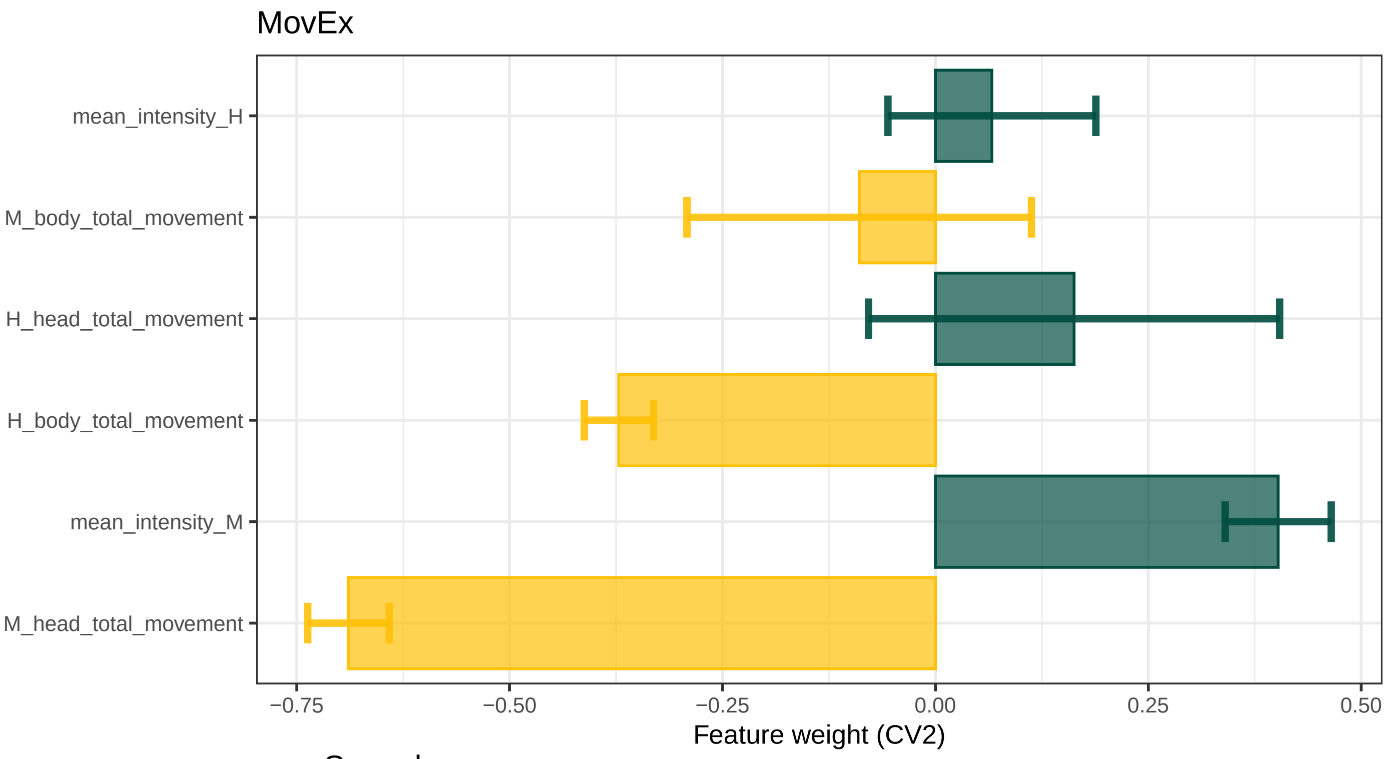
MovEx
mean_intensity_H
M_body_total_movement
H_head_total_movement
H_body_total_movement
mean_intensity_M
M_head_total_movement
−0.75
−0.50
−0.25
0.00
0.25
0.50
Feature weight (CV2)

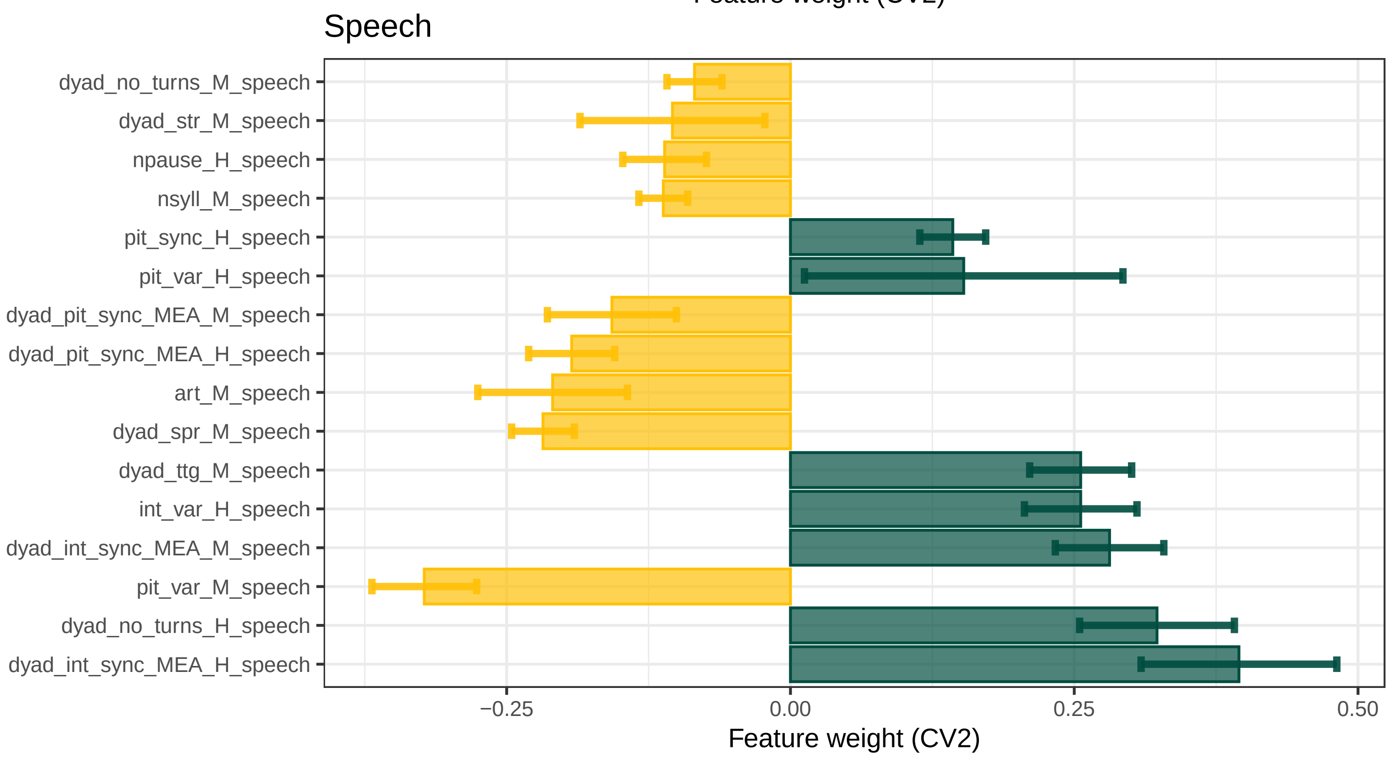
Speech
dyad_no_turns_M_speech
dyad_str_M_speech
npause_H_speech
nsyll_M_speech
pit_sync_H_speech
pit_var_H_speech
dyad_pit_sync_MEA_M_speech
dyad_pit_sync_MEA_H_speech
art_M_speech
dyad_spr_M_speech
dyad_ttg_M_speech
int_var_H_speech
dyad_int_sync_MEA_M_speech
pit_var_M_speech
dyad_no_turns_H_speech
dyad_int_sync_MEA_H_speech
−0.25
0.00
0.25
0.50
Feature weight (CV2)

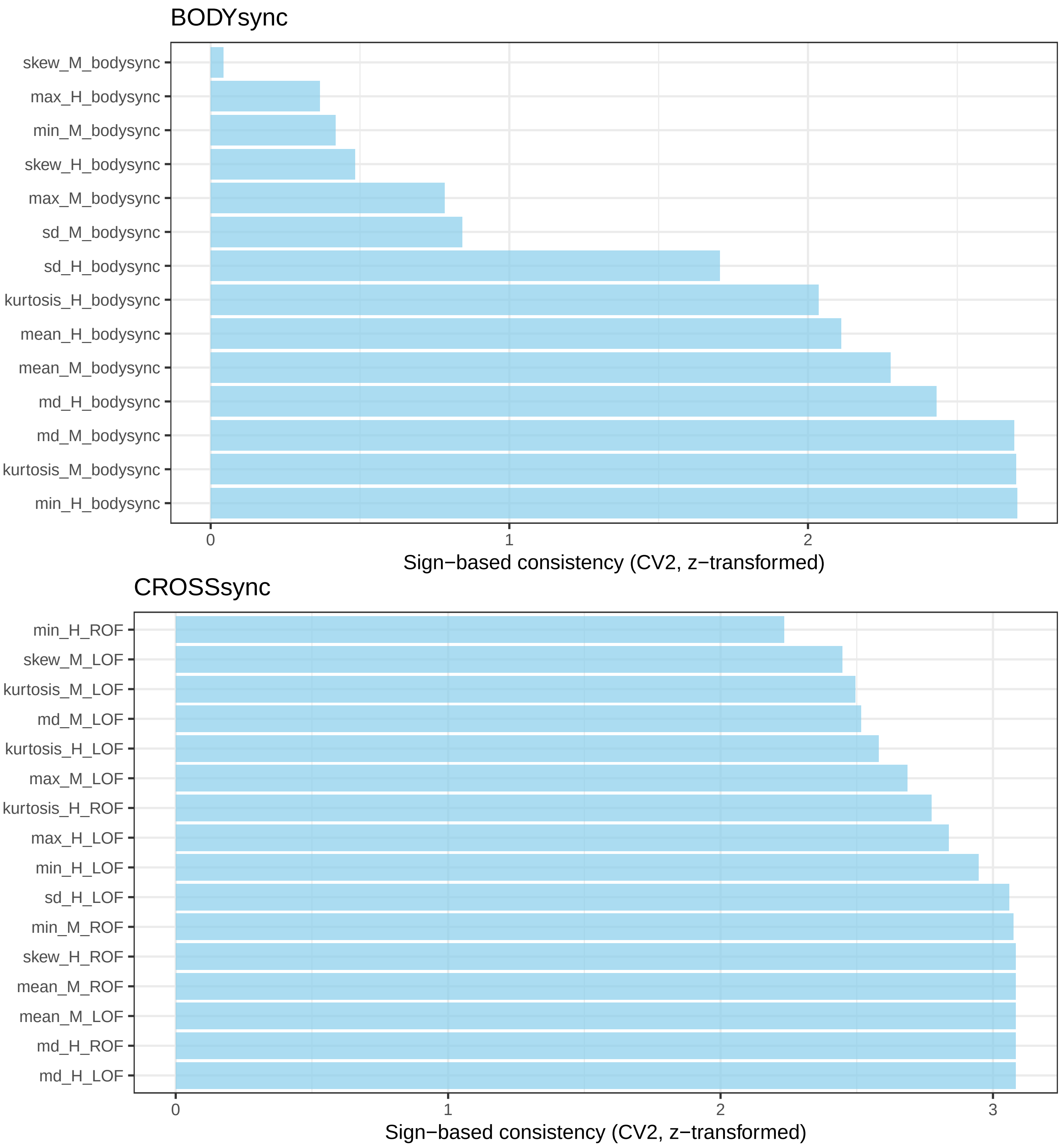
BODYsync
skew_M_bodysync
max_H_bodysync
min_M_bodysync
skew_H_bodysync
max_M_bodysync
sd_M_bodysync
sd_H_bodysync
kurtosis_H_bodysync
mean_H_bodysync
mean_M_bodysync
md_H_bodysync
md_M_bodysync
kurtosis_M_bodysync
min_H_bodysync
0
1
2
Sign−based consistency (CV2, z−transformed)
CROSSsync
min_H_ROF
skew_M_LOF
kurtosis_M_LOF
md_M_LOF
kurtosis_H_LOF
max_M_LOF
kurtosis_H_ROF
max_H_LOF
min_H_LOF
sd_H_LOF
min_M_ROF
skew_H_ROF
mean_M_ROF
mean_M_LOF
md_H_ROF
md_H_LOF
0
1
2
3
Sign−based consistency (CV2, z−transformed)

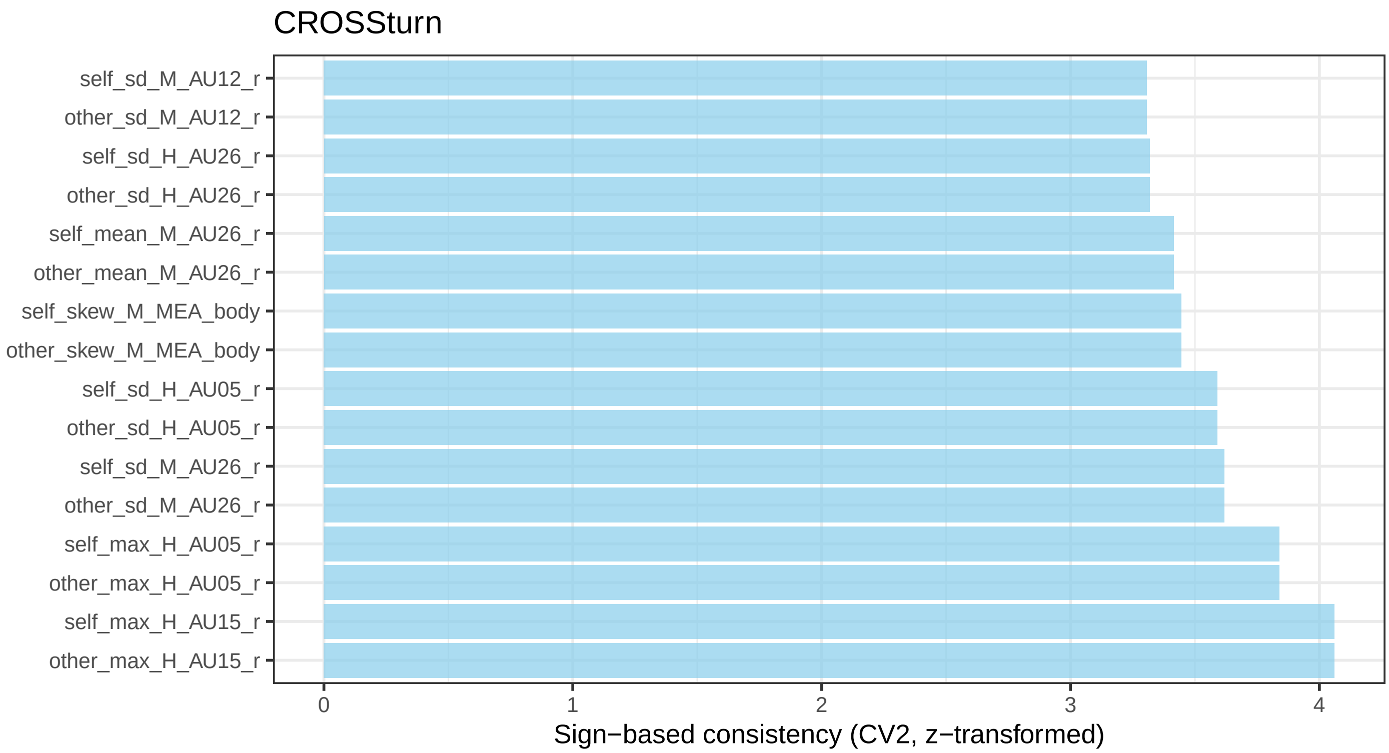
CROSSturn
self_sd_M_AU12_r
other_sd_M_AU12_r
self_sd_H_AU26_r
other_sd_H_AU26_r
self_mean_M_AU26_r
other_mean_M_AU26_r
self_skew_M_MEA_body
other_skew_M_MEA_body
self_sd_H_AU05_r
other_sd_H_AU05_r
self_sd_M_AU26_r
other_sd_M_AU26_r
self_max_H_AU05_r
other_max_H_AU05_r
self_max_H_AU15_r
other_max_H_AU15_r
0
1
2
3
4
Sign−based consistency (CV2, z−transformed)

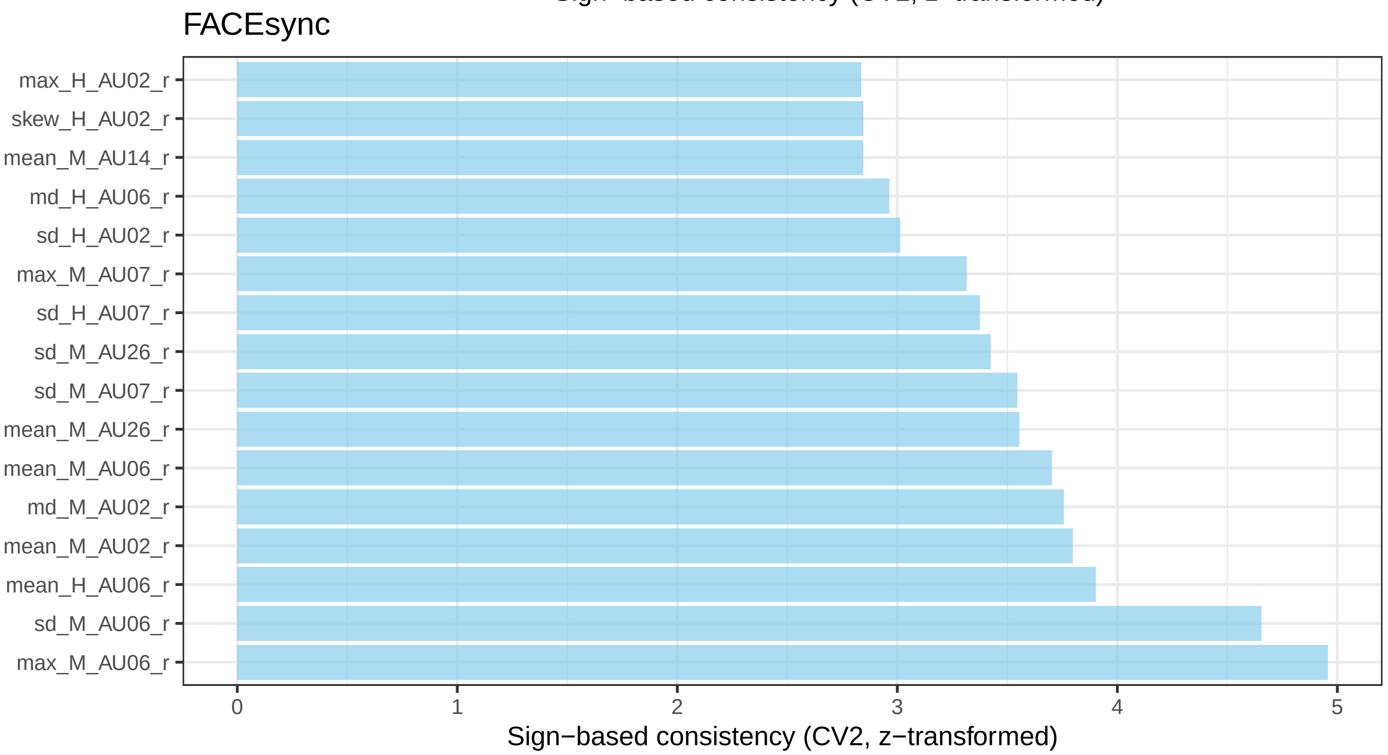
FACEsync
max_H_AU02_r
skew_H_AU02_r
mean_M_AU14_r
md_H_AU06_r
sd_H_AU02_r
max_M_AU07_r
sd_H_AU07_r
sd_M_AU26_r
sd_M_AU07_r
mean_M_AU26_r
mean_M_AU06_r
md_M_AU02_r
mean_M_AU02_r
mean_H_AU06_r
sd_M_AU06_r
max_M_AU06_r
0
1
2
3
4
5
Sign−based consistency (CV2, z−transformed)

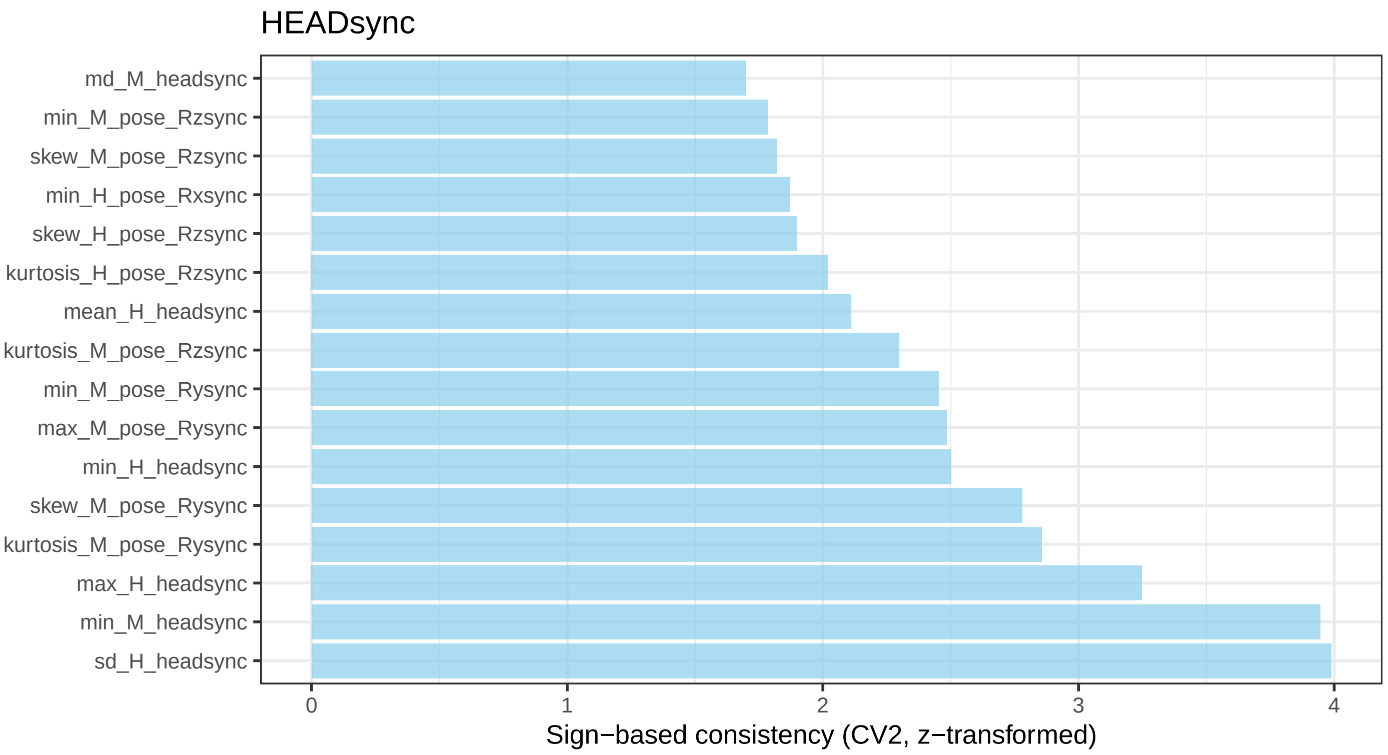
HEADsync
md_M_headsync
min_M_pose_Rzsync
skew_M_pose_Rzsync
min_H_pose_Rxsync
skew_H_pose_Rzsync
kurtosis_H_pose_Rzsync
mean_H_headsync
kurtosis_M_pose_Rzsync
min_M_pose_Rysync
max_M_pose_Rysync
min_H_headsync
skew_M_pose_Rysync
kurtosis_M_pose_Rysync
max_H_headsync
min_M_headsync
sd_H_headsync
0
1
2
3
4
Sign−based consistency (CV2, z−transformed)

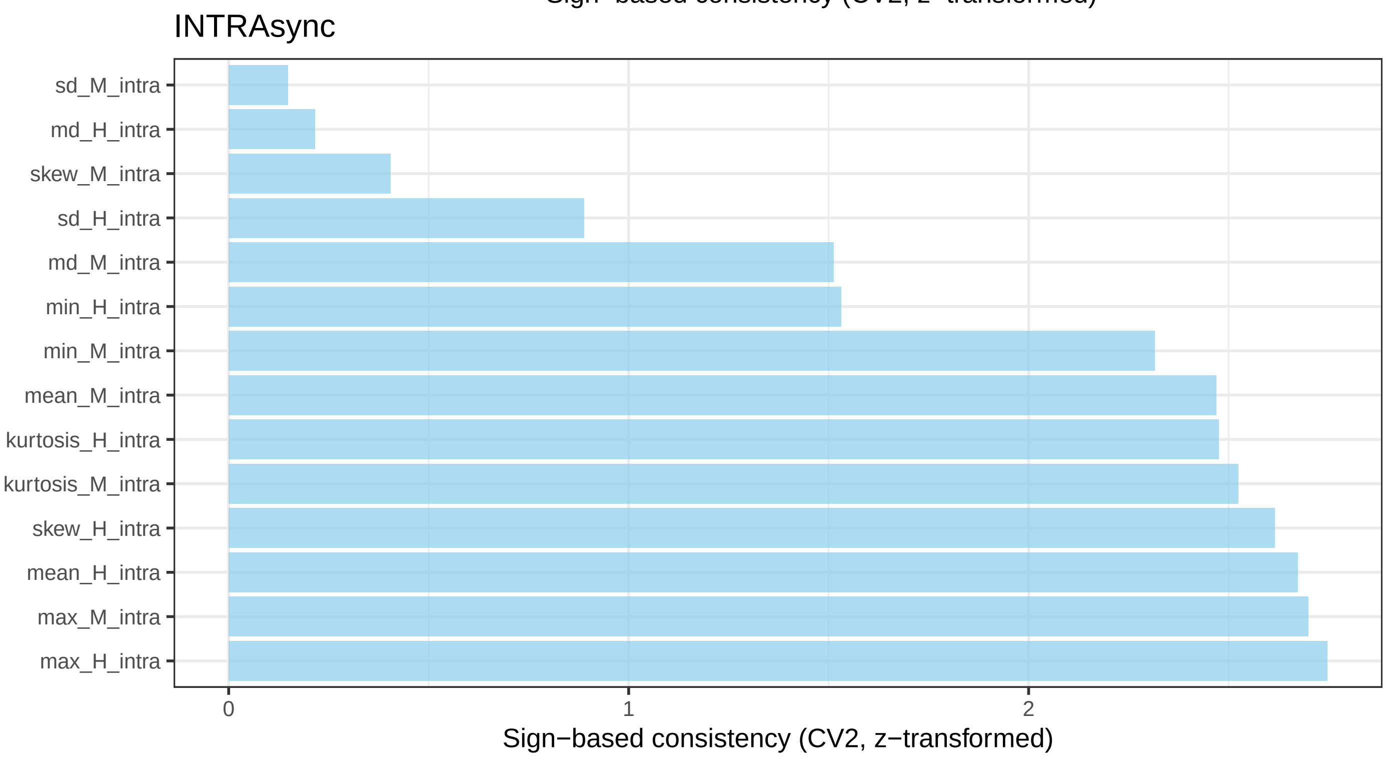
INTRAsync
sd_M_intra
md_H_intra
skew_M_intra
sd_H_intra
md_M_intra
min_H_intra
min_M_intra
mean_M_intra
kurtosis_H_intra
kurtosis_M_intra
skew_H_intra
mean_H_intra
max_M_intra
max_H_intra
0
1
2
Sign−based consistency (CV2, z−transformed)

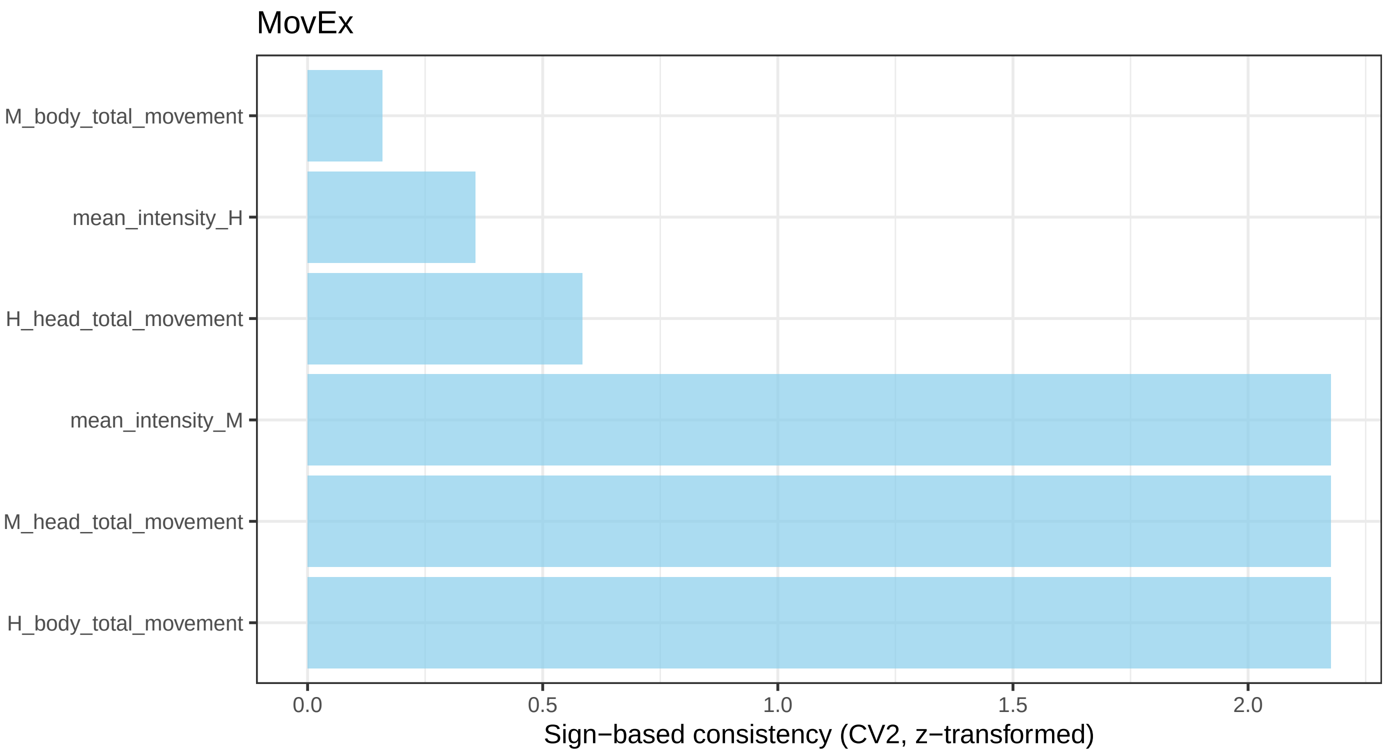

MovEx
M_body_total_movement
mean_intensity_H
H_head_total_movement
mean_intensity_M
M_head_total_movement
H_body_total_movement
0.0
0.5
1.0
1.5
2.0
Sign−based consistency (CV2, z−transformed)

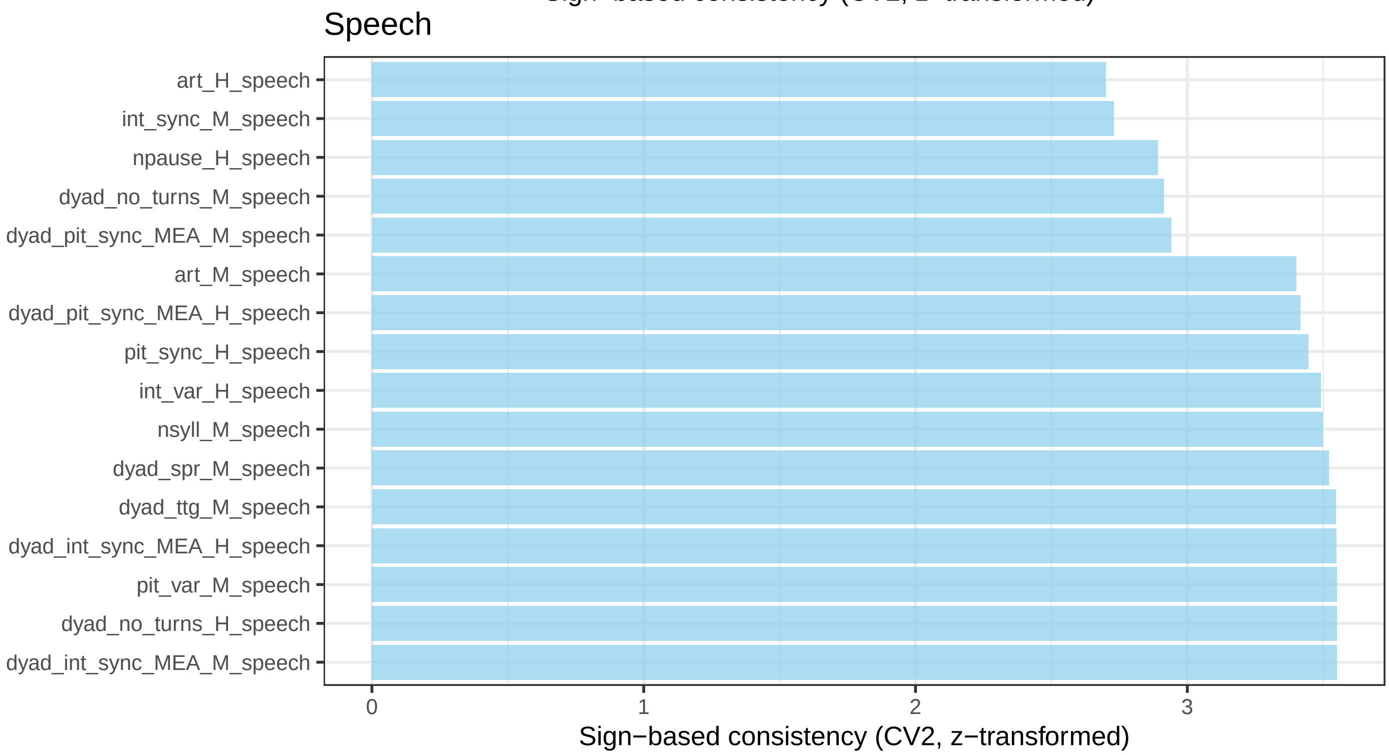

Speech
art_H_speech
int_sync_M_speech
npause_H_speech
dyad_no_turns_M_speech
dyad_pit_sync_MEA_M_speech
art_M_speech
dyad_pit_sync_MEA_H_speech
pit_sync_H_speech
int_var_H_speech
nsyll_M_speech
dyad_spr_M_speech
dyad_ttg_M_speech
dyad_int_sync_MEA_H_speech
pit_var_M_speech
dyad_no_turns_H_speech
dyad_int_sync_MEA_M_speech
0
1
2
3
Sign−based consistency (CV2, z−transformed)

##### S6.3 ASD-involved versus COMP classifiers

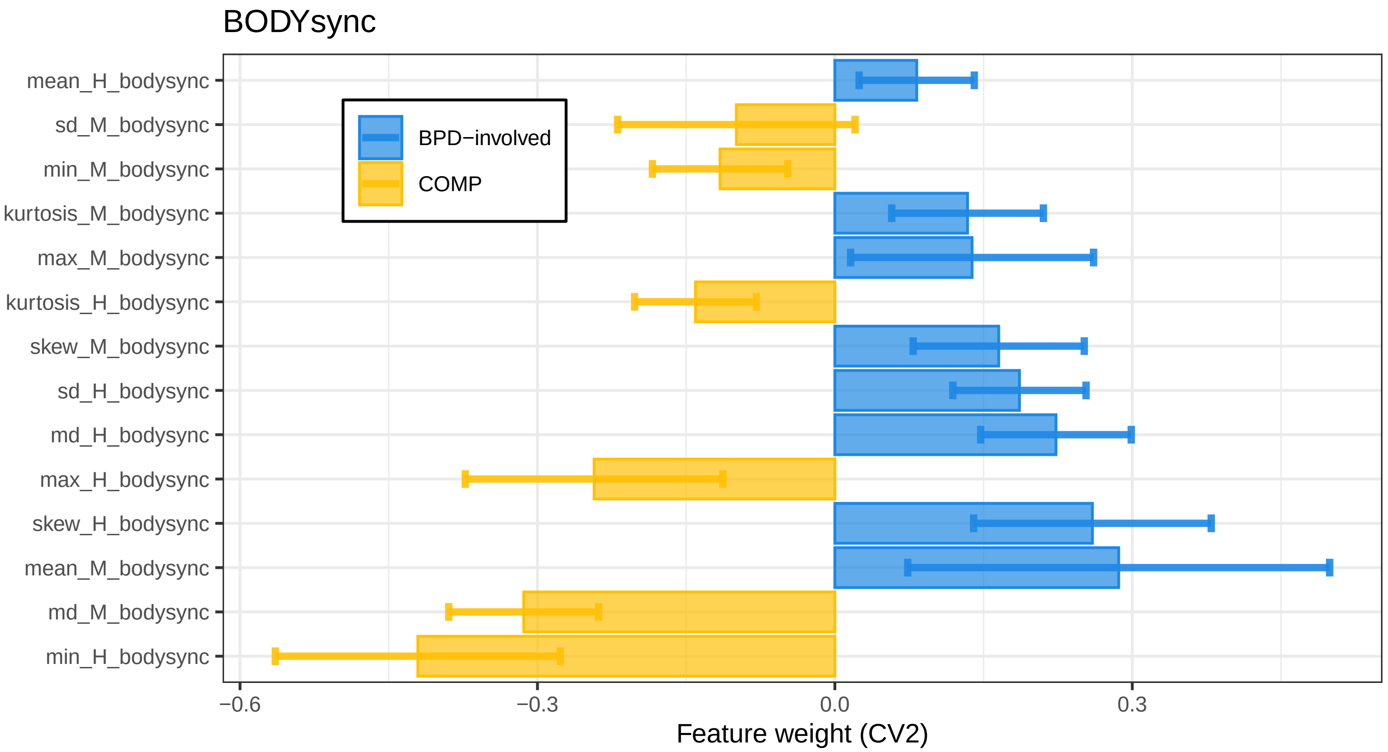

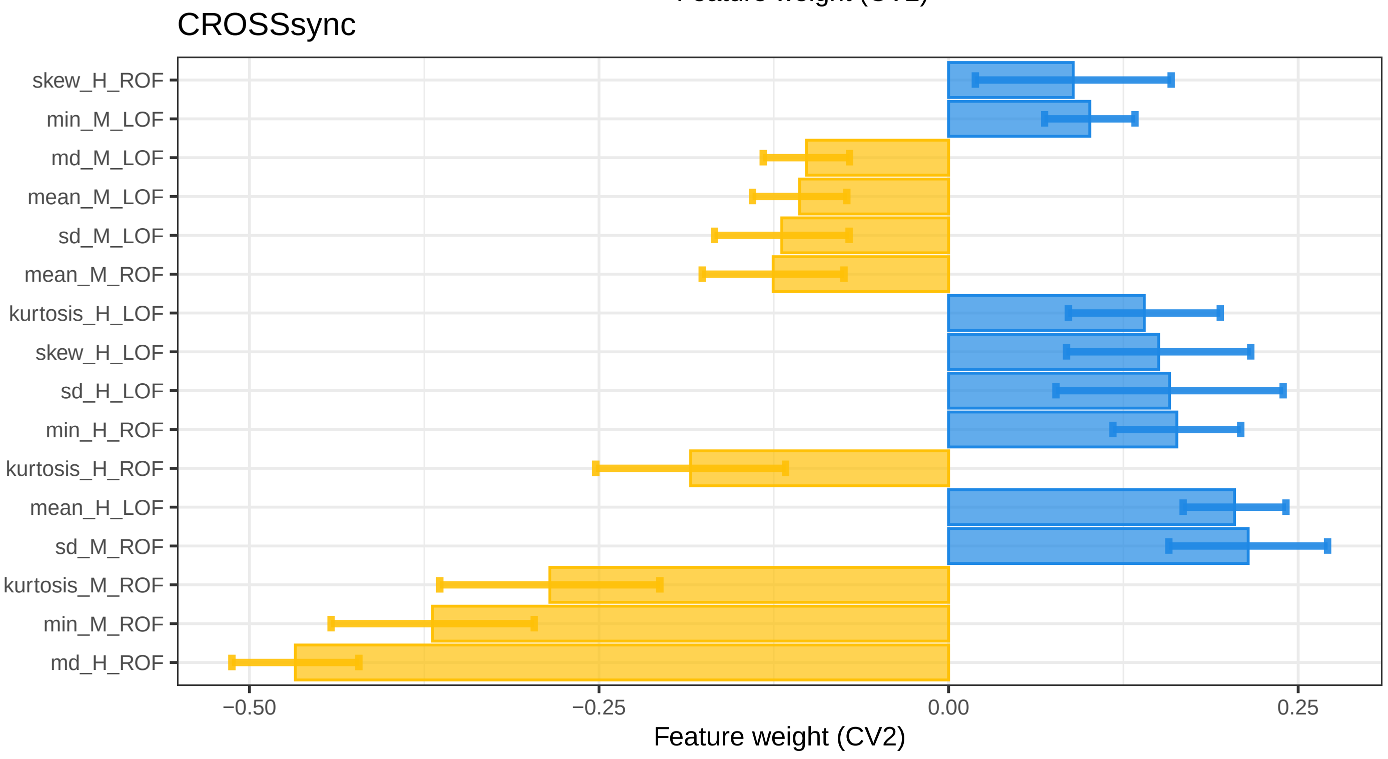

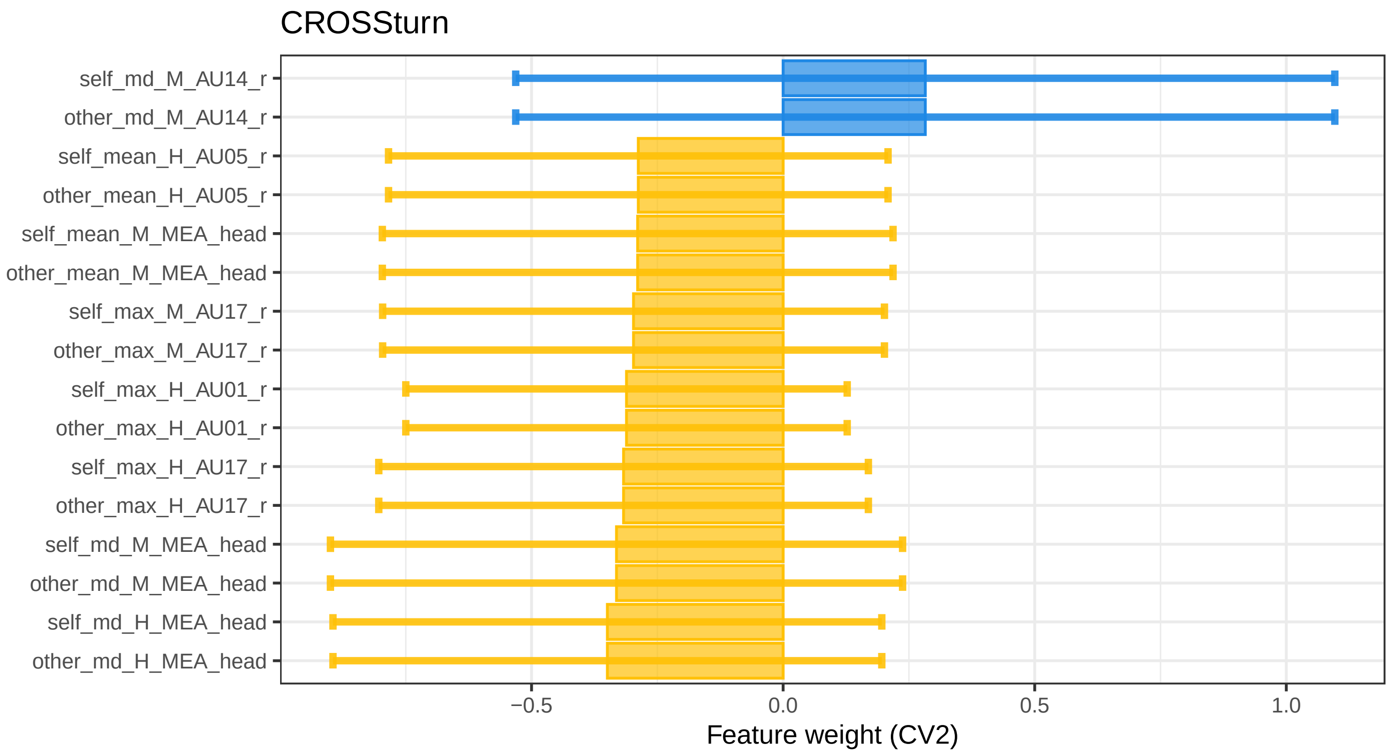
CROSSturn
self_md_M_AU14_r
other_md_M_AU14_r
self_mean_H_AU05_r
other_mean_H_AU05_r
self_mean_M_MEA_head
other_mean_M_MEA_head
self_max_M_AU17_r
other_max_M_AU17_r
self_max_H_AU01_r
other_max_H_AU01_r
self_max_H_AU17_r
other_max_H_AU17_r
self_md_M_MEA_head
other_md_M_MEA_head
self_md_H_MEA_head
other_md_H_MEA_head
−0.5
0.0
0.5
1.0
Feature weight (CV2)

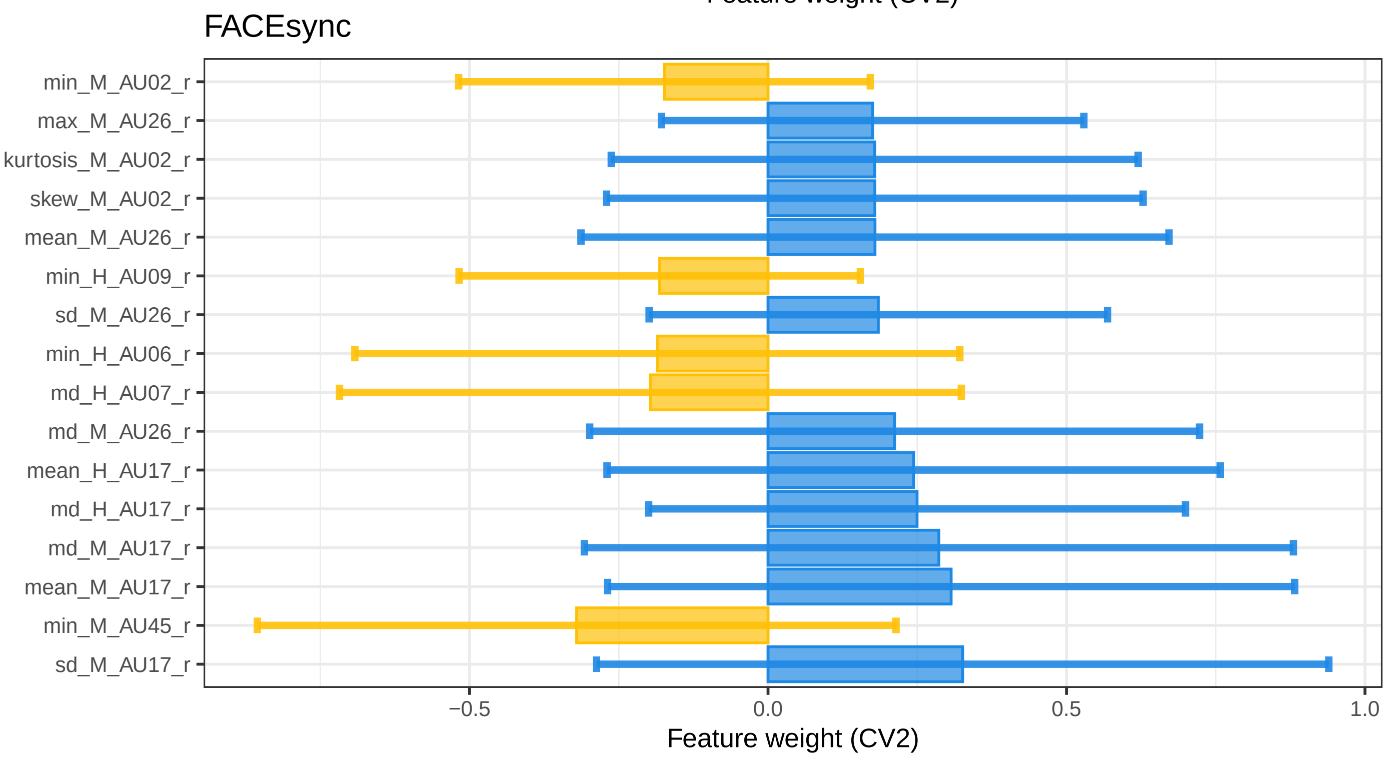
FACEsync
min_M_AU02_r
max_M_AU26_r
kurtosis_M_AU02_r
skew_M_AU02_r
mean_M_AU26_r
min_H_AU09_r
sd_M_AU26_r
min_H_AU06_r
md_H_AU07_r
md_M_AU26_r
mean_H_AU17_r
md_H_AU17_r
md_M_AU17_r
mean_M_AU17_r
min_M_AU45_r
sd_M_AU17_r
−0.5
0.0
0.5
1.0
Feature weight (CV2)

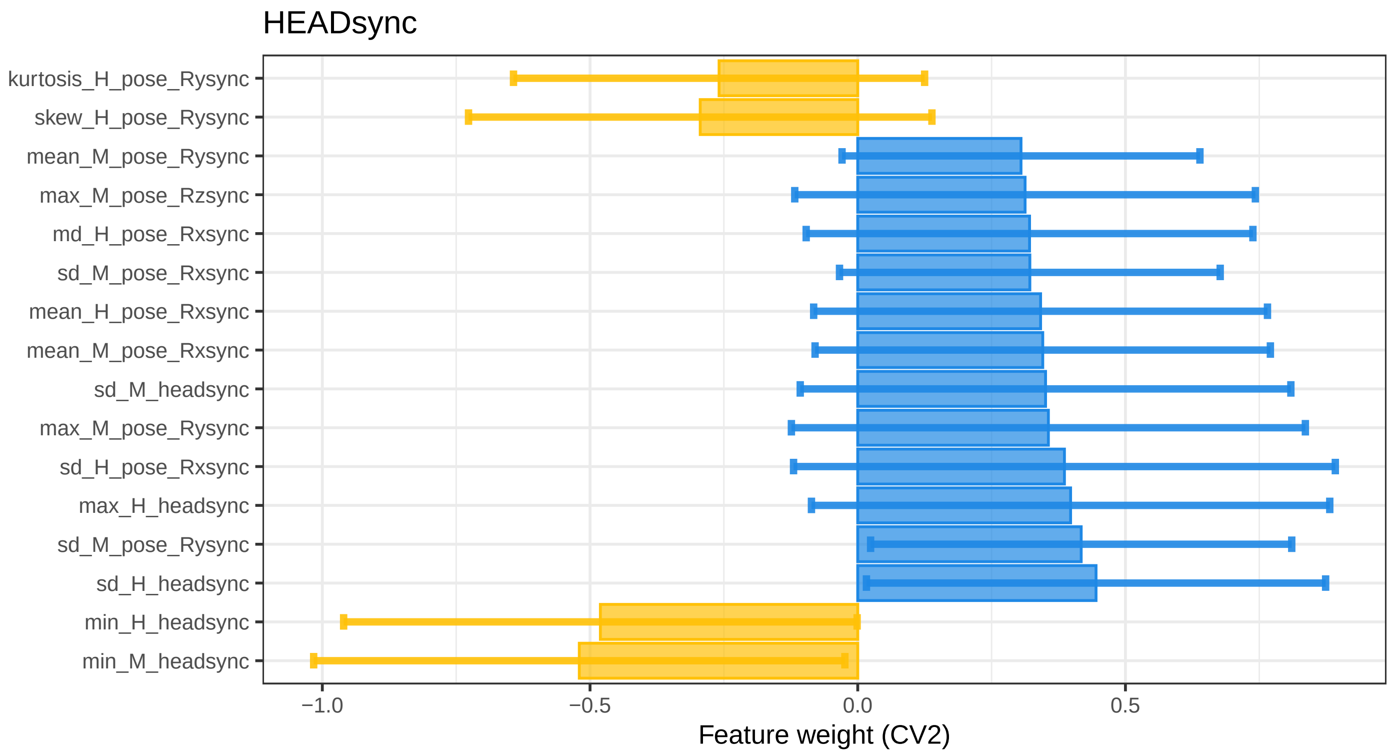
HEADsync
kurtosis_H_pose_Rysync
skew_H_pose_Rysync
mean_M_pose_Rysync
max_M_pose_Rzsync
md_H_pose_Rxsync
sd_M_pose_Rxsync
mean_H_pose_Rxsync
mean_M_pose_Rxsync
sd_M_headsync
max_M_pose_Rysync
sd_H_pose_Rxsync
max_H_headsync
sd_M_pose_Rysync
sd_H_headsync
min_H_headsync
min_M_headsync
−1.0
−0.5
0.0
0.5
Feature weight (CV2)

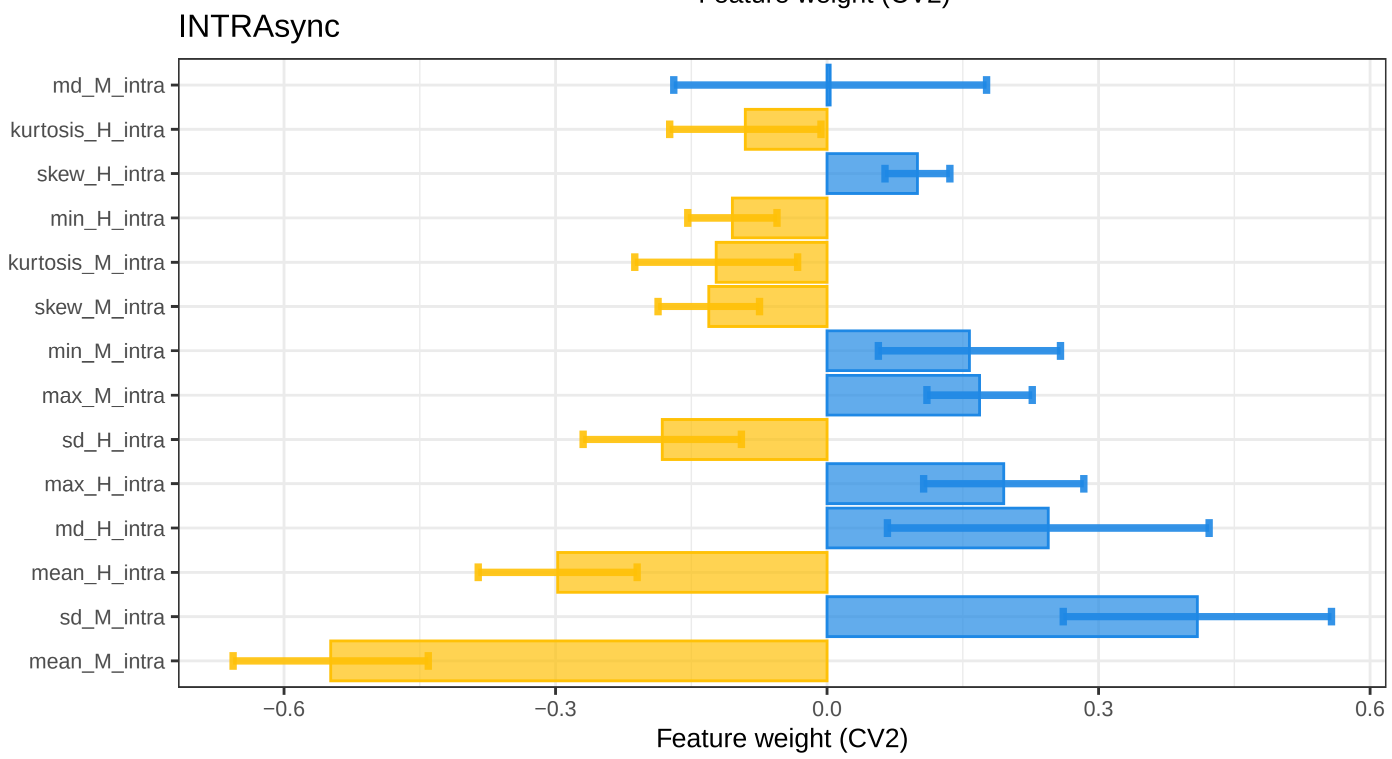
INTRAsync
md_M_intra
kurtosis_H_intra
skew_H_intra
min_H_intra
kurtosis_M_intra
skew_M_intra
min_M_intra
max_M_intra
sd_H_intra
max_H_intra
md_H_intra
mean_H_intra
sd_M_intra
mean_M_intra
−0.6
−0.3
0.0
0.3
0.6
Feature weight (CV2)

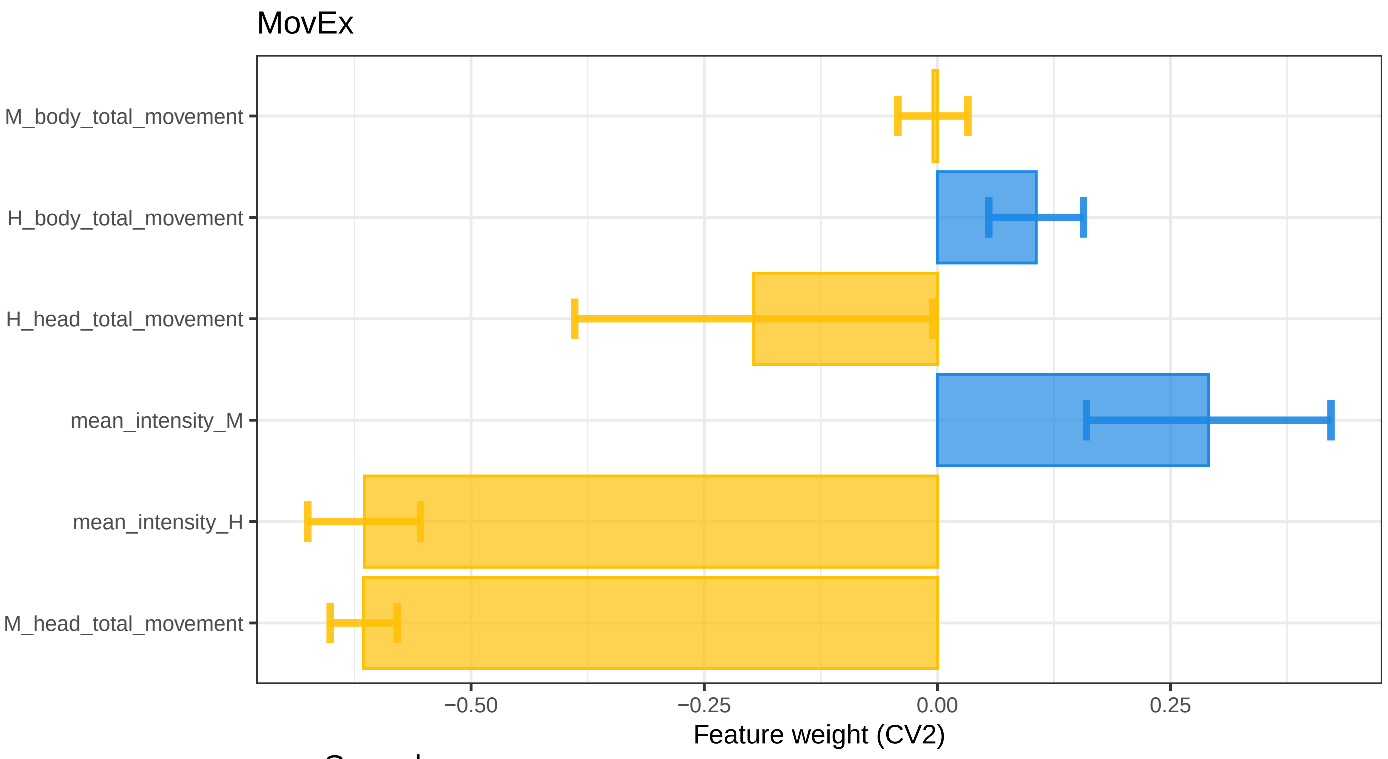
MovEx
M_body_total_movement
H_body_total_movement
H_head_total_movement
mean_intensity_M
mean_intensity_H
M_head_total_movement
−0.50
−0.25
0.00
0.25
Feature weight (CV2)

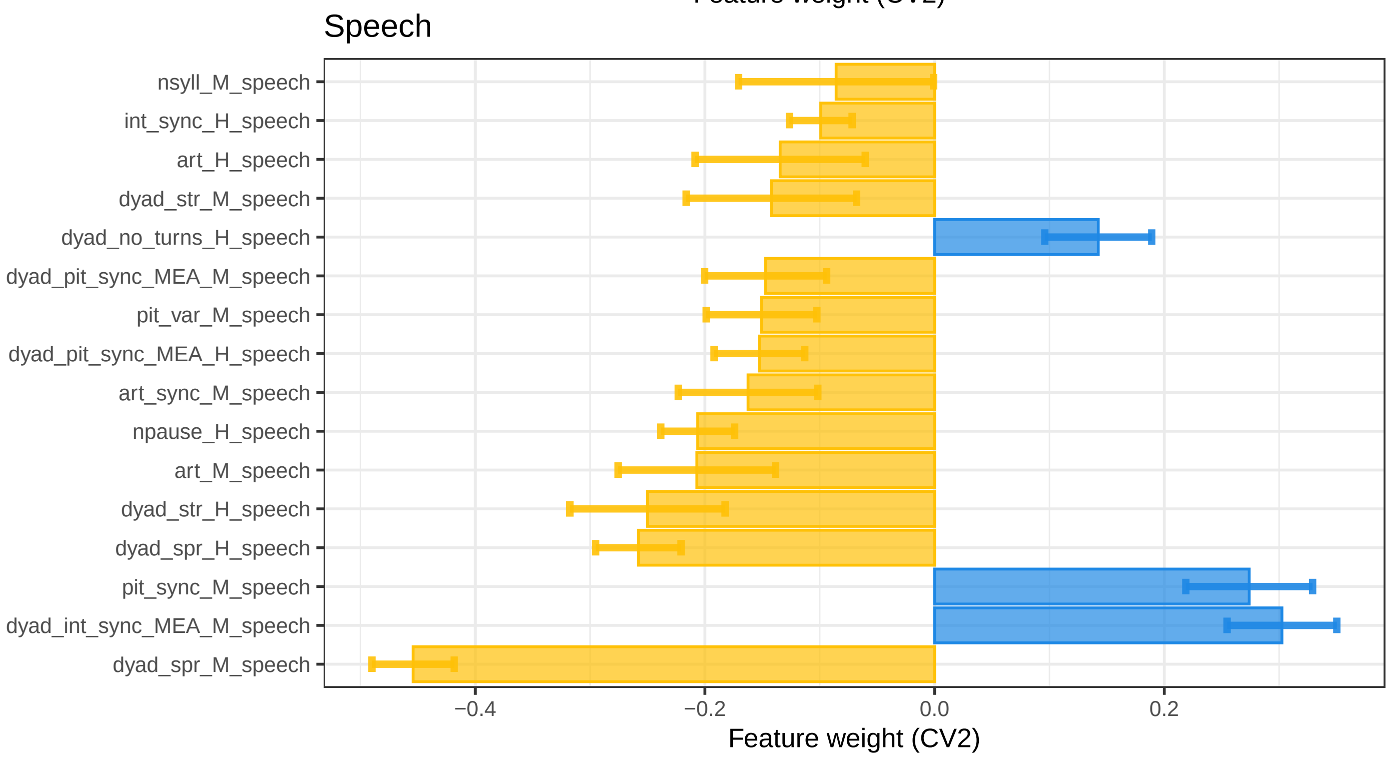
Speech
nsyll_M_speech
int_sync_H_speech
art_H_speech
dyad_str_M_speech
dyad_no_turns_H_speech
dyad_pit_sync_MEA_M_speech
pit_var_M_speech
dyad_pit_sync_MEA_H_speech
art_sync_M_speech
npause_H_speech
art_M_speech
dyad_str_H_speech
dyad_spr_H_speech
pit_sync_M_speech
dyad_int_sync_MEA_M_speech
dyad_spr_M_speech
−0.4
−0.2
0.0
0.2
Feature weight (CV2)

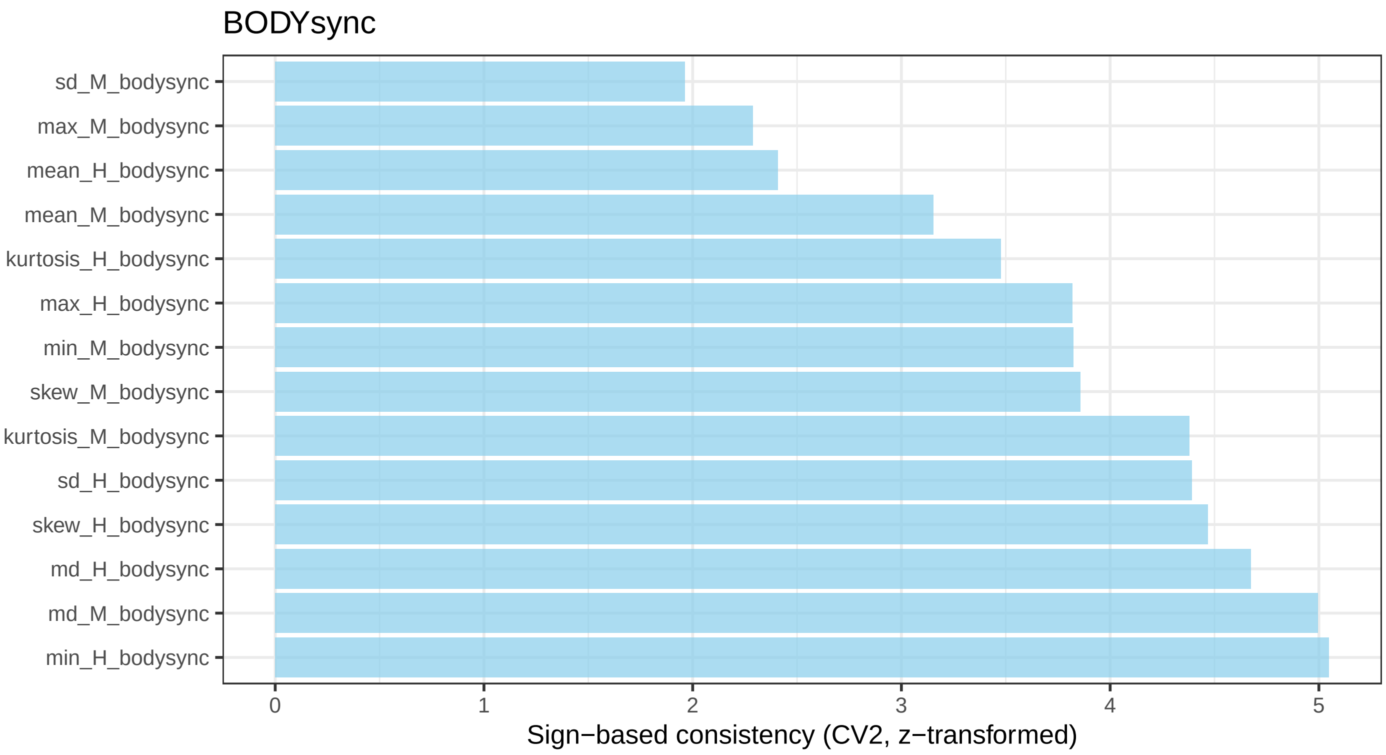
BODYsync
sd_M_bodysync
max_M_bodysync
mean_H_bodysync
mean_M_bodysync
kurtosis_H_bodysync
max_H_bodysync
min_M_bodysync
skew_M_bodysync
kurtosis_M_bodysync
sd_H_bodysync
skew_H_bodysync
md_H_bodysync
md_M_bodysync
min_H_bodysync
0
1
2
3
4
5
Sign−based consistency (CV2, z−transformed)

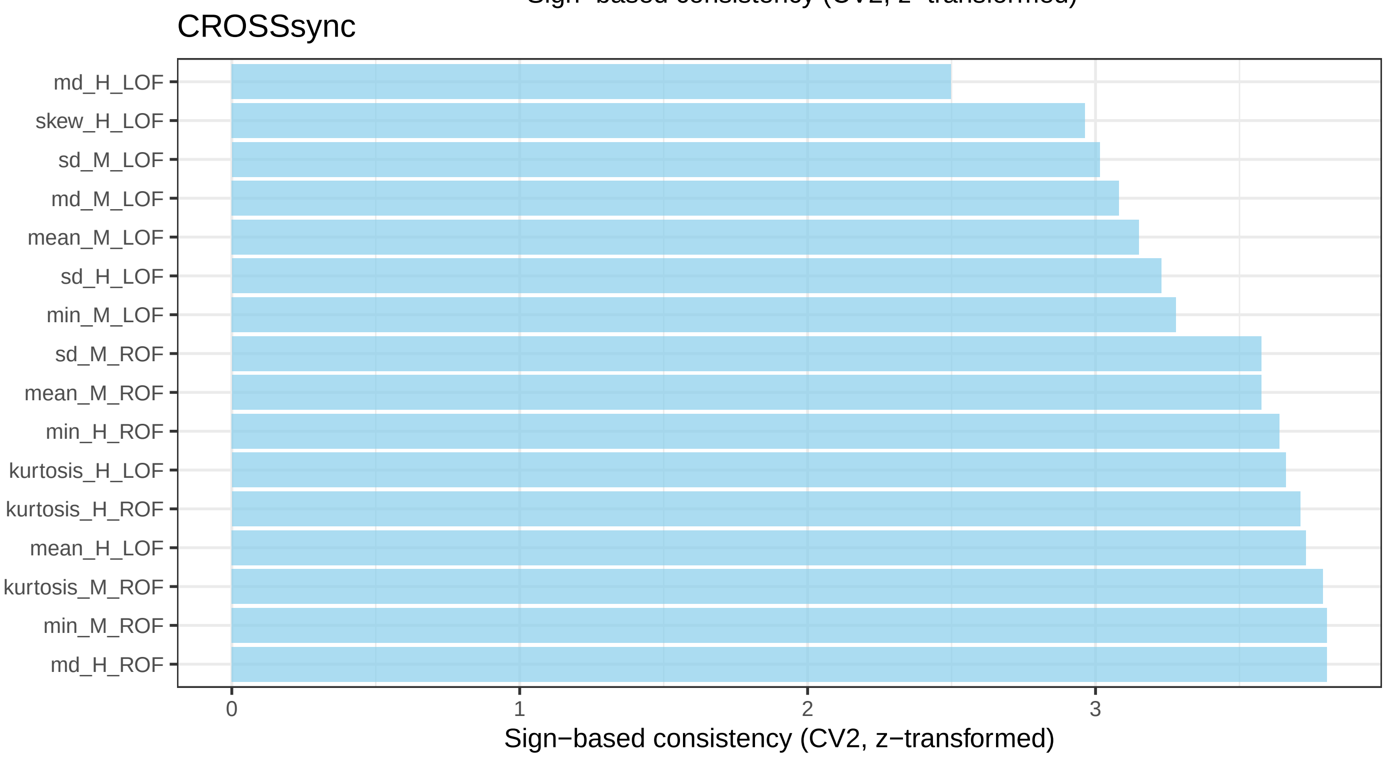
CROSSsync
md_H_LOF
skew_H_LOF
sd_M_LOF
md_M_LOF
mean_M_LOF
sd_H_LOF
min_M_LOF
sd_M_ROF
mean_M_ROF
min_H_ROF
kurtosis_H_LOF
kurtosis_H_ROF
mean_H_LOF
kurtosis_M_ROF
min_M_ROF
md_H_ROF
0
1
2
3
Sign−based consistency (CV2, z−transformed)

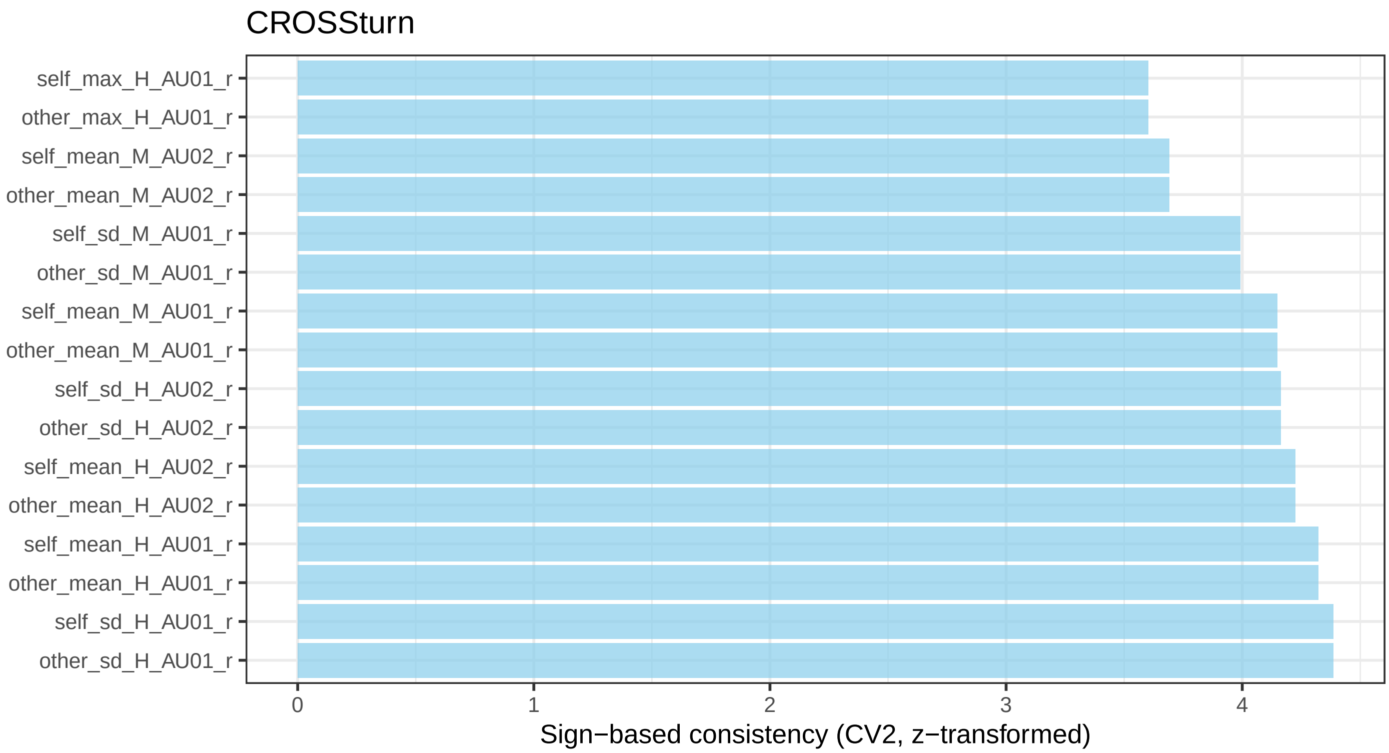
CROSSturn
self_max_H_AU01_r
other_max_H_AU01_r
self_mean_M_AU02_r
other_mean_M_AU02_r
self_sd_M_AU01_r
other_sd_M_AU01_r
self_mean_M_AU01_r
other_mean_M_AU01_r
self_sd_H_AU02_r
other_sd_H_AU02_r
self_mean_H_AU02_r
other_mean_H_AU02_r
self_mean_H_AU01_r
other_mean_H_AU01_r
self_sd_H_AU01_r
other_sd_H_AU01_r
0
1
2
3
4
Sign−based consistency (CV2, z−transformed)

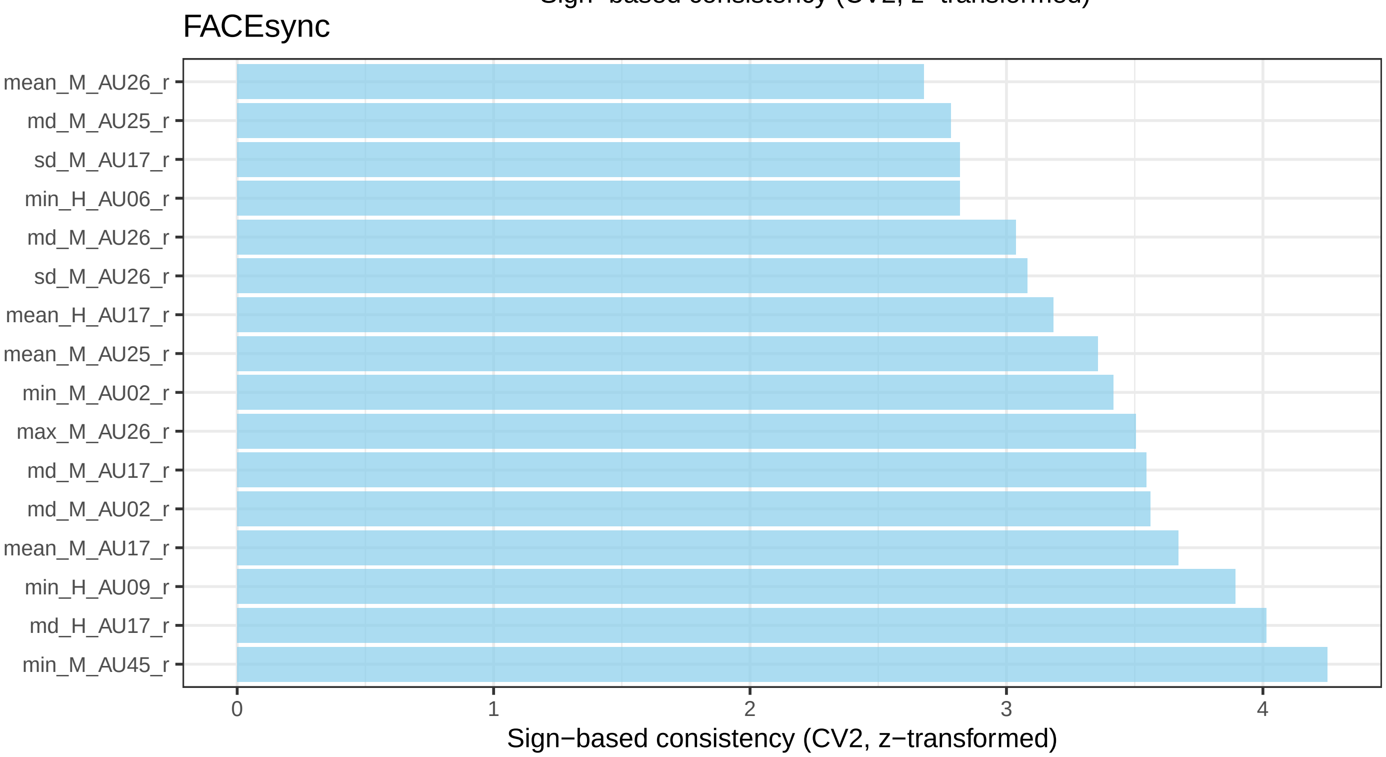
FACEsync
mean_M_AU26_r
md_M_AU25_r
sd_M_AU17_r
min_H_AU06_r
md_M_AU26_r
sd_M_AU26_r
mean_H_AU17_r
mean_M_AU25_r
min_M_AU02_r
max_M_AU26_r
md_M_AU17_r
md_M_AU02_r
mean_M_AU17_r
min_H_AU09_r
md_H_AU17_r
min_M_AU45_r
0
1
2
3
4
Sign−based consistency (CV2, z−transformed)

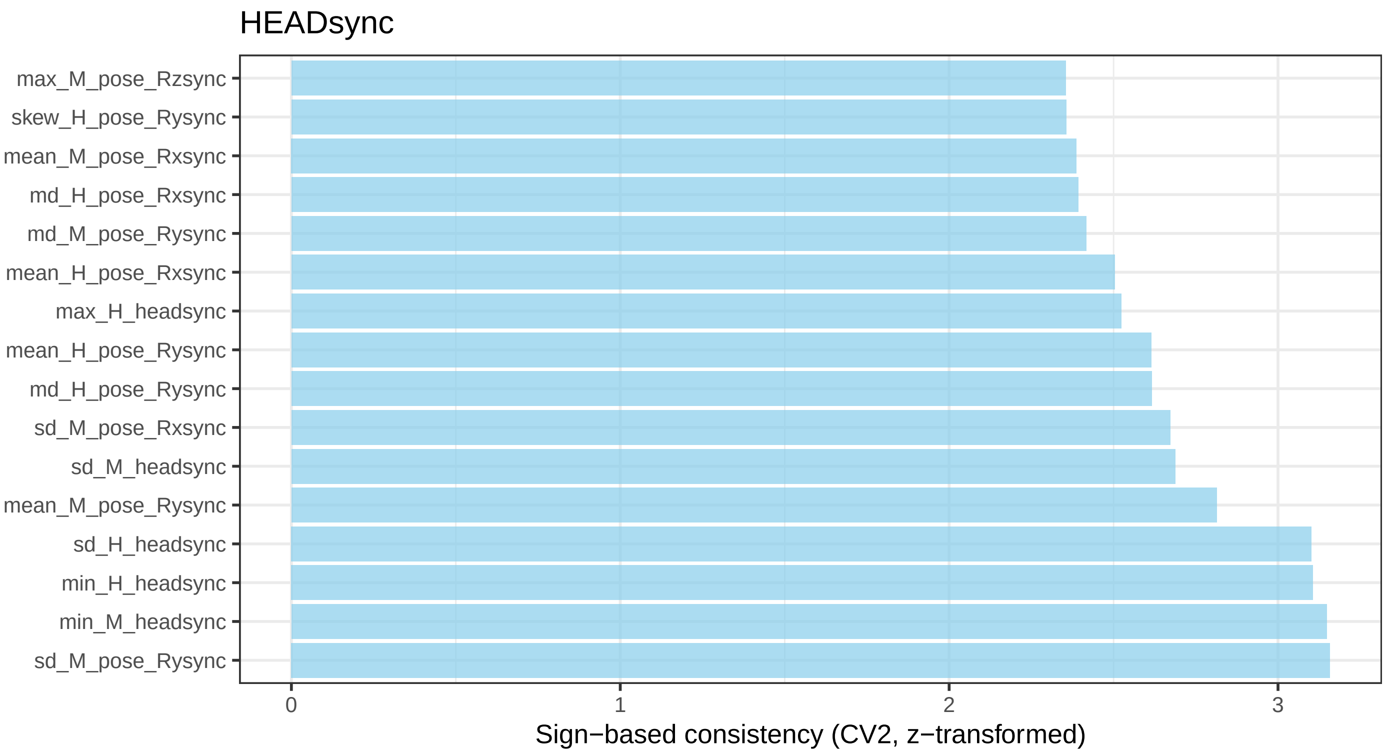
HEADsync
max_M_pose_Rzsync
skew_H_pose_Rysync
mean_M_pose_Rxsync
md_H_pose_Rxsync
md_M_pose_Rysync
mean_H_pose_Rxsync
max_H_headsync
mean_H_pose_Rysync
md_H_pose_Rysync
sd_M_pose_Rxsync
sd_M_headsync
mean_M_pose_Rysync
sd_H_headsync
min_H_headsync
min_M_headsync
sd_M_pose_Rysync
0
1
2
3
Sign−based consistency (CV2, z−transformed)

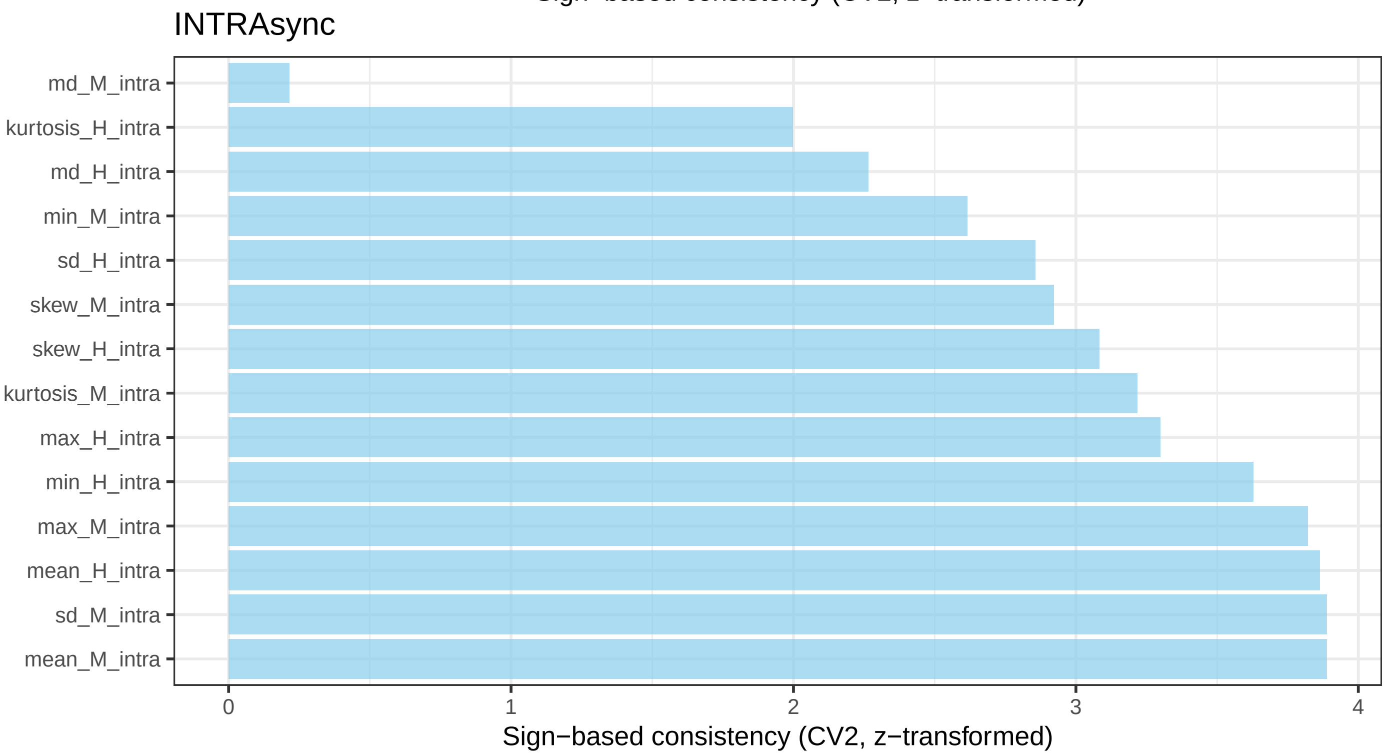
INTRAsync
md_M_intra
kurtosis_H_intra
md_H_intra
min_M_intra
sd_H_intra
skew_M_intra
skew_H_intra
kurtosis_M_intra
max_H_intra
min_H_intra
max_M_intra
mean_H_intra
sd_M_intra
mean_M_intra
0
1
2
3
4
Sign−based consistency (CV2, z−transformed)

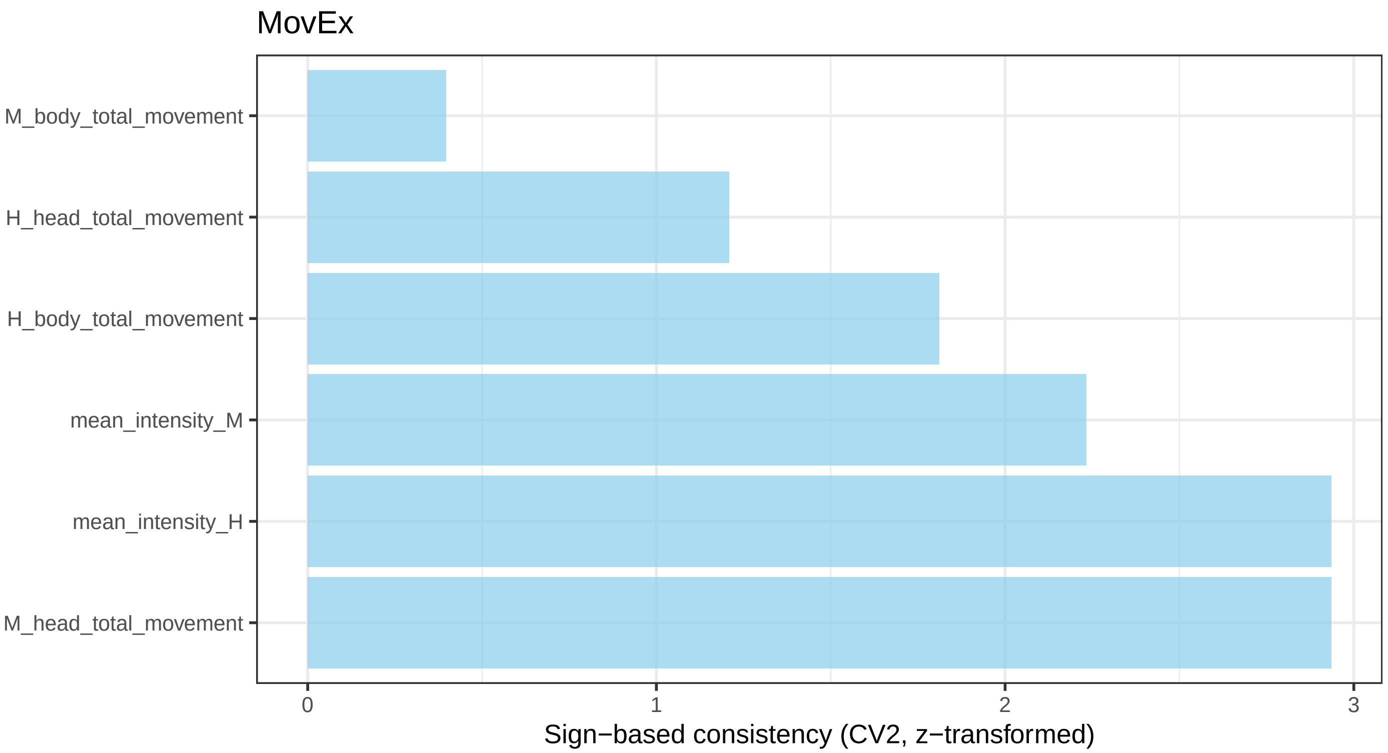

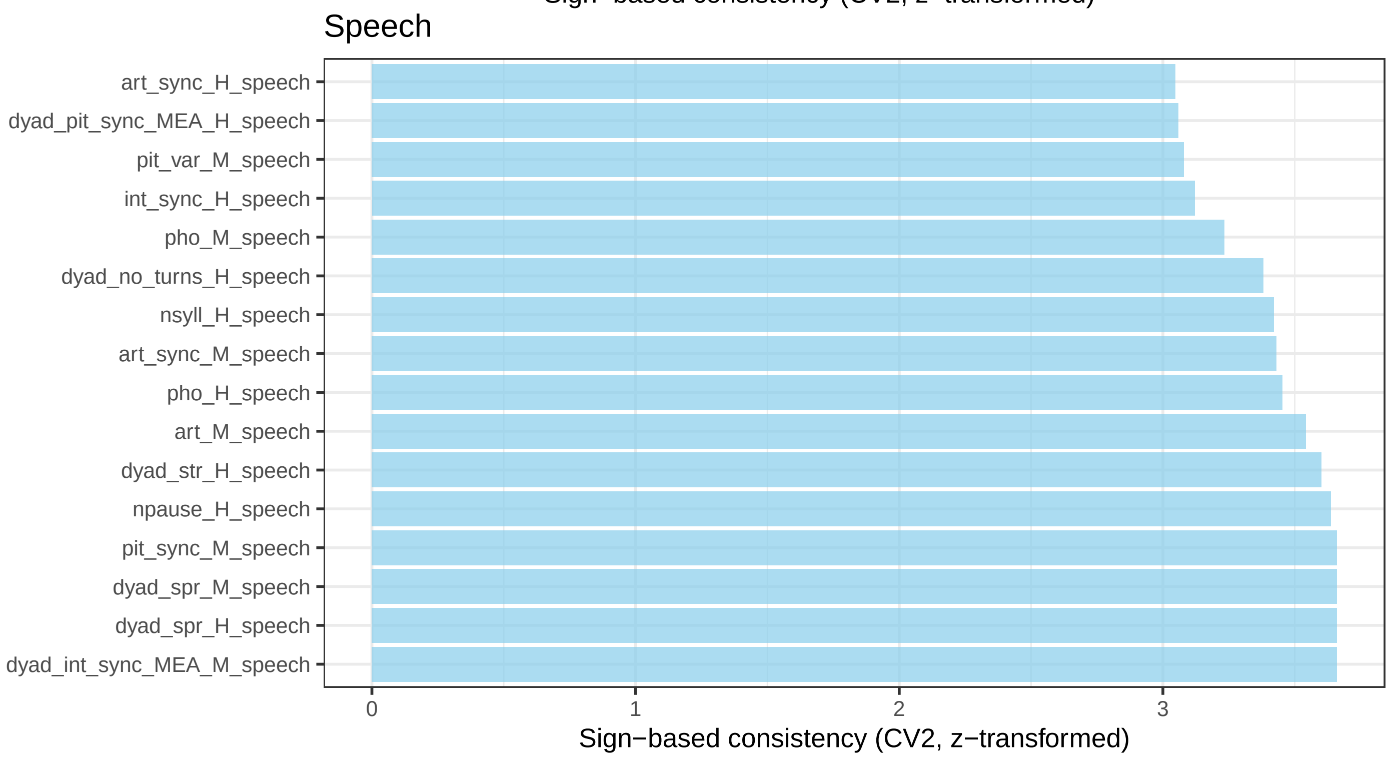
